# Bioactive glycans in a microbiome-directed food for malnourished children

**DOI:** 10.1101/2023.08.14.23293998

**Authors:** Matthew C. Hibberd, Daniel M. Webber, Dmitry A. Rodionov, Suzanne Henrissat, Robert Y. Chen, Cyrus Zhou, Hannah M. Lynn, Yi Wang, Hao-Wei Chang, Evan M. Lee, Janaki Lelwala-Guruge, Marat D. Kazanov, Aleksandr A. Arzamasov, Semen A. Leyn, Vincent Lombard, Nicolas Terrapon, Bernard Henrissat, Juan J. Castillo, Garret Couture, Nikita P. Bacalzo, Ye Chen, Carlito B. Lebrilla, Ishita Mostafa, Subhasish Das, Mustafa Mahfuz, Michael J. Barratt, Andrei L. Osterman, Tahmeed Ahmed, Jeffrey I. Gordon

## Abstract

Evidence is accumulating that perturbed postnatal development of the gut microbiome contributes to childhood malnutrition^1–4^. Designing effective microbiome-directed therapeutic foods to repair these perturbations requires knowledge about how food components interact with the microbiome to alter its expressed functions. Here we use biospecimens from a randomized, controlled trial of a microbiome-directed complementary food prototype (MDCF-2) that produced superior rates of weight gain compared to a conventional ready-to-use supplementary food (RUSF) in 12-18-month-old Bangladeshi children with moderate acute malnutrition (MAM)^4^. We reconstructed 1000 bacterial genomes (metagenome-assembled genomes, MAGs) present in their fecal microbiomes, identified 75 whose abundances were positively associated with weight gain (change in weight-for-length Z score, WLZ), characterized gene expression changes in these MAGs as a function of treatment type and WLZ response, and used mass spectrometry to quantify carbohydrate structures in MDCF-2 and feces. The results reveal treatment-induced changes in expression of carbohydrate metabolic pathways in WLZ-associated MAGs. Comparing participants consuming MDCF-2 versus RUSF, and MDCF-2-treated children in the upper versus lower quartiles of WLZ responses revealed that two *Prevotella copri* MAGs positively associated with WLZ were principal contributors to MDCF-2-induced expression of metabolic pathways involved in utilization of its component glycans. Moreover, the predicted specificities of carbohydrate active enzymes expressed by polysaccharide utilization loci (PULs) in these two MAGs correlate with the (i) *in vitro* growth of Bangladeshi *P. copri* strains, possessing differing degrees of PUL and overall genomic content similarity to these MAGs, cultured in defined medium containing different purified glycans representative of those in MDCF-2, and (ii) levels of carbohydrate structures identified in feces from clinical trial participants. In the accompanying paper^5^, we use a gnotobiotic mouse model colonized with age- and WLZ-associated bacterial taxa cultured from this study population, and fed diets resembling those consumed by study participants, to directly test the relationship between *P. copri*, MDCF-2 glycan metabolism, host ponderal growth responses, and intestinal gene expression and metabolism. The ability to identify bioactive glycan structures in MDCFs that are metabolized by growth-associated bacterial taxa will help guide recommendations about use of this MDCF for children with acute malnutrition representing different geographic locales and ages, as well as enable development of bioequivalent, or more efficacious, formulations composed of culturally acceptable and affordable ingredients.

---

The global health challenge of childhood undernutrition is great; in 2020, an estimated 149 million children under five years of age were stunted (low height-for-age) while 45 million exhibited wasting (low weight-for-length, WLZ)^6^. Undernutrition and its long-term sequelae are the leading causes of morbidity and mortality in this age range. Sequelae include persistent impairments in linear growth, immune and metabolic functions, and neurodevelopment - all of which have proven to be largely resistant to current interventions^7^. Although food insecurity is not the sole driver of undernutrition^8^, the profound disruption of economies and food systems by the COVID-19 pandemic has greatly exacerbated this global health challenge^9^.

Studies of healthy members of birth cohorts living in several countries have identified shared features of gut microbial community assembly – a process that is largely completed by the end of the second postnatal year^10,11^. Children with moderate or severe acute malnutrition (MAM or SAM) have impaired ponderal growth (wasting). Their microbial community development is perturbed, resulting in microbiota configurations that resemble those of chronologically younger children^10^. The metabolic maturation of malnourished children is also compromised compared to their healthy peers^12^. Colonization of gnotobiotic mice with fecal microbiota samples collected from healthy children or from chronologically age-matched children with acute malnutrition disclosed that microbial communities from the latter transmitted impaired weight gain and altered bone growth phenotypes, plus produced immune and metabolic abnormalities^1,2,13^.

We have used gnotobiotic mouse and piglet models to design MDCF formulations for repairing the microbial communities of children with MAM. MAM is defined as having a WLZ score that is 2-3 standard deviations below the median of a multi-national cohort of age-matched healthy children. In a 3-month randomized controlled feeding study of 12-18-month-old Bangladeshi children with MAM, we demonstrated that a lead formulation (MDCF-2) produced significant improvement in the rate of weight gain (β-WLZ) compared to a conventional RUSF that was not designed to alter the gut microbiota^4^. The superior effect of MDCF-2 on β-WLZ occurred even though its caloric density is 15% lower than RUSF. Plasma proteomic analyses revealed 70 proteins whose levels had statistically significant positive correlations with the change in WLZ, including mediators of musculoskeletal growth and neurodevelopment. These proteins were increased to a significantly greater degree in MDCF-2-treated children compared to those receiving RUSF. Levels of several proteins involved in immunoinflammatory processes were negatively correlated with WLZ and significantly reduced by MDCF-2 treatment^4^. Sequencing PCR amplicons generated from bacterial 16S rRNA genes present in fecal biospecimens disclosed 23 bacterial taxa that were significantly associated with WLZ; 21 were positively associated while two were negatively associated. The abundances of the positively associated taxa increased to a significantly greater degree with MDCF-2 compared to RUSF treatment^4^.

In this study, we reconstruct the genomes of bacteria present in the gut communities of participants in the completed trial, identify metabolic pathways that are differentially expressed in response to MDCF-2 in MAGs that are positively associated with WLZ, and determine how their differential expression relates to the processing of components of MDCF-2 and ponderal growth responses. The results highlight the remarkable strain specificity of microbiome responses and point to two *Prevotella copri* strains as key mediators of MDCF-2 glycan metabolism and host ponderal responses. In the accompanying study, we directly test these relationships using a gnotobiotic mouse model.

## Results

### Reconstructing bacterial genomes associated with ponderal growth responses

**Fig. 1a** summarizes the design of the randomized, controlled feeding study of children with MAM, aged 15.4±2.0 months (mean±SD) at enrollment. These children lived in an impoverished urban area (Mirpur) located in Dhaka, Bangladesh. The 3-month intervention involved twice-daily dietary supplementation with either MDCF-2 or RUSF (2 x 25g servings, providing ∼220-250 kcal/day)^4^. A total of 59 children in each treatment group completed the intervention and a one month follow-up; fecal samples were collected every 10 days during the first month and every four weeks thereafter. There were no statistically significant differences in the amount of nutritional supplement consumed between children receiving MDCF-2 versus RUSF, no differences in the proportion of children who satisfied World Health Organization requirements for minimum meal frequency or minimum acceptable diet, and no differences in the amount of breast milk consumed between the two treatment groups^4^.

**Fig. 1.**
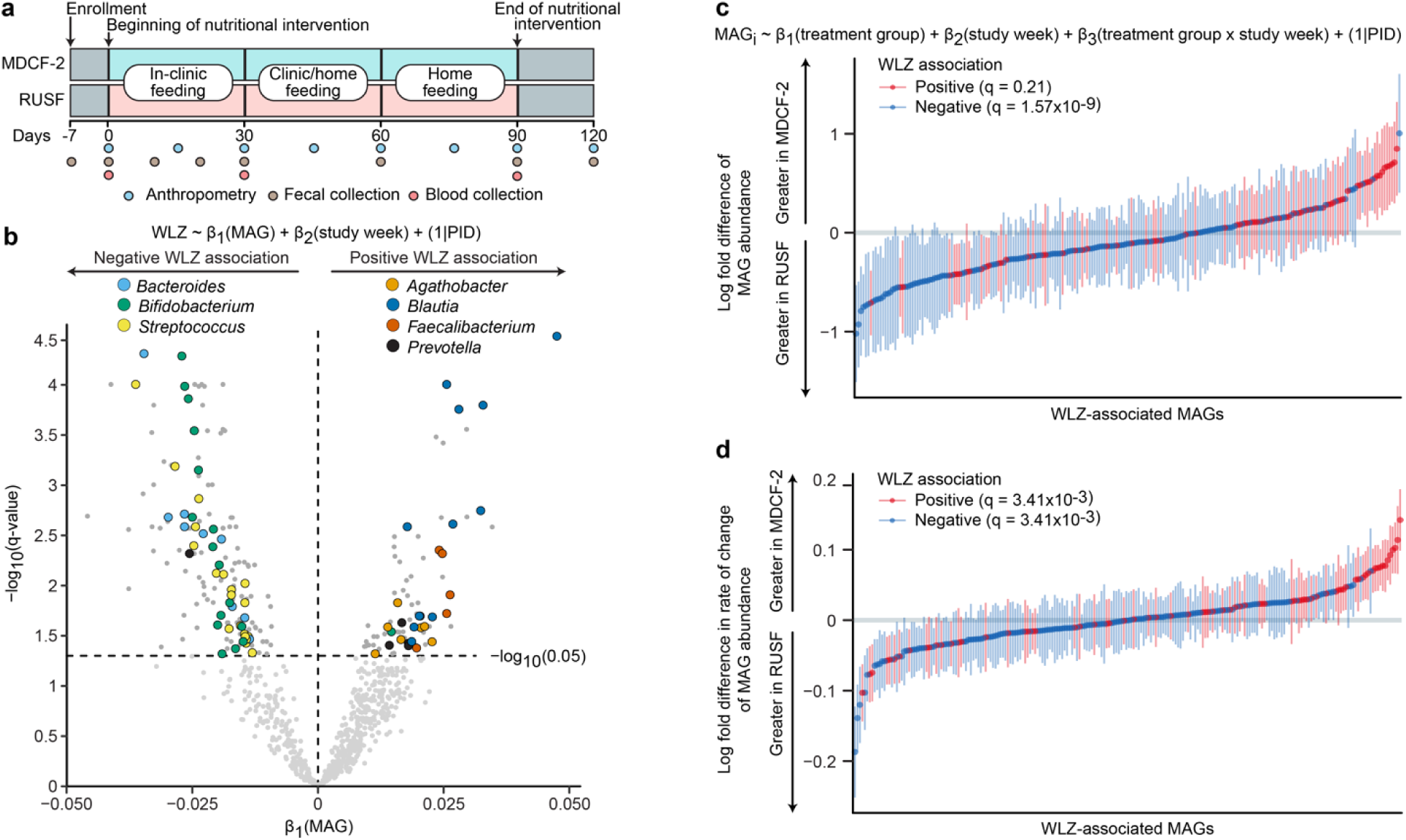
Identification of ponderal growth (WLZ)-associated MAGs. (**a**) Study design. (**b**) Volcano plot indicating the results of linear mixed effects modeling of the relationship between MAG abundance and WLZ scores for all trial participants, irrespective of treatment. Bacterial genera that are prevalent in the list of MAGs significantly associated with WLZ are colored by their taxonomic classification. Abbreviation: PID, participant identifier. (**c,d**) Results of gene set enrichment analysis (GSEA) of WLZ-associated MAGs ranked by the magnitude of their difference in abundance (panel c) or by their change in ‘abundance over time’ (panel d) in response to MDCF-2 versus RUSF treatment. Plotted values indicate the mean log_2_-fold difference (±SEM) in each model coefficient between the two treatment groups. The statistical significance of enrichment (q-value, GSEA) of MAGs that are positively or negatively associated with WLZ is shown.

To reconstruct the genomes of bacterial taxa present in the gut microbiomes of study participants, we isolated DNA from all fecal samples (n=942; 7-8 samples/participant) and performed short-read shotgun sequencing. DNA recovered from fecal biospecimens collected at t=0 and 3 months from the subset of participants comprising the upper quartile of the ponderal growth response to MDCF-2 (n=15)^4^ were subjected to additional long-read sequencing. We assembled pooled shotgun sequencing data from each participant’s fecal samples (short-read only, or short-plus long-reads when available) and aggregated contigs into metagenome-assembled genomes (MAGs) (**Extended Data Fig. 1a**; *Supplementary Methods*; *Supplementary Discussion*). The resulting set of 1,000 high-quality MAGs (defined as ≥90% complete and ≤5% contaminated based on marker gene analysis; **Supplementary Table 1a**) represented 65.6±8.0% and 66.2±7.9% of all quality controlled, paired-end shotgun reads generated from all 942 fecal DNA samples analyzed in the MDCF-2 and RUSF treatment groups, respectively [2.3±1.4×10^7^ 150 nt paired-end reads/sample (mean±SD); **Supplementary Table 2a**]. Taxonomy was assigned to MAGs using a consensus approach that included marker gene and kmer-based classification together with the Genome Taxonomy Database (GTDB; **Supplementary Table 1a**)^14^. Abundances were calculated for each MAG in the 707 fecal samples that spanned the beginning of treatment through the 1-month post-intervention time point and for which matching anthropometric measurements from children had been collected. A total of 837 MAGs satisfied our abundance and prevalence thresholds (see *Methods* and **Supplementary Table 2b**). We then employed linear mixed effects models to identify 222 MAGs whose abundances were significantly associated with WLZ [β_1_(MAG), q<0.05, **Fig. 1b**] over the 90-day course of the intervention and 30-day follow-up (see **Supplementary Table 3** for the 75 positively associated and 147 negatively associated MAGs). MAGs that were significantly positively associated with WLZ were predominantly members of the genera *Agathobacter*, *Blautia*, *Faecalibacterium* and *Prevotella*, while members of *Bacteroides, Bifidobacterium* and *Streptococcus* were prevalent among MAGs negatively associated with WLZ (**Fig. 1b**, **Extended Data Fig. 2**, **Supplementary Table 3**).

Changes in MAG abundances were subsequently modeled as a function of treatment group, study week, and the interaction between treatment group and study week, controlling for repeated measurements taken from the same individual (see equation in **Fig. 1c** and *Methods*). The ‘treatment group’ coefficient describes the mean difference in abundance of a given MAG between the MDCF-2 and RUSF groups over the course of the intervention (**Fig. 1c**), while the interaction coefficient in the equation describes the difference in the rate of change in abundance of a given MAG (**Fig. 1d**). Restricting this analysis to the time of initiation of treatment did not reveal any statistically significant differences in MAG abundances between the two groups (q>0.05, one linear model per MAG; **Supplementary Table 2b**). Expanding the analysis to include all time points from initiation to end of treatment disclosed that although no individual MAG abundances were significantly associated with MDCF-2 or RUSF consumption, MAGs whose abundances increased faster in the MDCF-2 compared to the RUSF group were significantly enriched for those positively associated with WLZ [q=3.41×10^-3^, gene set enrichment analysis (GSEA); **Fig. 1d**]. In contrast, MAGs with a higher mean abundance as well as those that increased more rapidly in RUSF-treated children were significantly enriched for those negatively associated with WLZ (q=1.57×10^-9^ and q=3.41×10^-3^, respectively; GSEA) (**Fig. 1c,d; Supplementary Table 4**).

We utilized a ‘subsystems’ approach adapted from the SEED genome annotation platform^15,16^ to identify genes that comprise metabolic pathways represented in WLZ-associated MAGs. To do so, genes were aligned to a reference collection of 2,856 human gut bacterial genomes that had been subjected to *in silico* reconstructions of metabolic pathways reflecting major nutrient biosynthetic and degradative capabilities in mcSEED, a microbial community-centered implementation of SEED^17^. We employed this reference collection and procedures described in **Extended Data Fig. 3** and *Supplementary Methods* to assign putative functions to a subset of 199,334 proteins in the 1,000 MAGs (**Supplementary Table 5**); these proteins, which represented 1,308 non-redundant functions, formed the basis for predicting which of 106 metabolic pathways were present or absent in each MAG. This effort generated a set of inferred metabolic phenotypes for each MAG (**Supplementary Tables 6** and **7**). GSEA disclosed multiple metabolic pathways involved in carbohydrate utilization that were significantly enriched in WLZ-associated MAGs (q<0.05), and in MAGs ranked by their changes in abundance in response to MDCF-2 compared to RUSF treatment. While other non-carbohydrate pathways were also identified using this approach (*e.g*., those involved in aspects of amino acid and bile acid metabolism), pathways involved in carbohydrate utilization predominated (*P* = 0.006, Fisher’s test; **Extended Data Fig. 4**; **Supplementary Table 8**).

### Carbohydrate composition of MDCF-2 and RUSF

Prior to analyzing the transcriptional responses of MAGs to each nutritional intervention, we characterized the carbohydrates present in MDCF-2 and RUSF, as well as their constituent ingredients [chickpea flour, soybean flour, peanut paste and mashed green banana pulp in the case of MDCF-2; rice, lentil and milk powder in the case of RUSF (**Supplementary Table 9a**)]. Ultrahigh-performance liquid chromatography-triple quadrupole mass spectrometry (UHPLC-QqQ-MS) was used to quantify 14 monosaccharides and 49 unique glycosidic linkages. Polysaccharide content was defined using a procedure in which polysaccharides were chemically cleaved into oligosaccharides, after which the structures of these liberated oligosaccharides were then used to characterize and quantify their ‘parent’ polysaccharide^18^.

The results revealed that L-arabinose, D-xylose, L-fucose, D-mannose, and D-galacturonic acid (GalA) are significantly more abundant in MDCF-2 (*P*<0.05; t-test), as are 14 linkages, eight of which contain these monosaccharides (**Extended Data Fig. 5a,b; Supplementary Table 9b,c,e,f)**. Integrating the quantitative polysaccharide and glycoside linkage data allowed us to conclude that MDCF-2 contains a significantly greater abundance of galactans and mannans than RUSF (*P<*0.05; t-test), while RUSF contains significantly more starch and cellulose (*P*<0.05; t-test) (**Fig. 2a; Supplementary Table 9d,g**). Galactans are represented in MDCF-2 as unbranched β-1,4-linked galactan as well as arabinogalactan I (**Fig. 2b**). Mannans are present as unbranched β-1,4-linked mannan (β-mannan), galactomannan and glucomannan (**Fig. 2c**). Arabinan is abundant in both formulations, although the representation of arabinose and glycosidic linkages containing arabinose is significantly greater in MDCF-2 than in RUSF (see **Extended Data Fig. 5a,b** and **Supplementary Table 9e,f** for results of statistical tests). Arabinan in MDCF-2 is largely derived from its soybean, banana, and chickpea components, while in RUSF this polysaccharide originates from rice and lentil (**Fig. 2a**). Arabinans in both formulations share a predominant 1,5-linked-L-arabinofuranose (Ara*f*) backbone. Soybean arabinans are characterized by diverse side chains composed of 1,2- and 1,3*-*linked-L-Ara*f* connected by 1,2,3-, 1,2,5-, and 1,3,5-L-Ara*f* branch points, while chickpea, lentil, and banana arabinans primarily contain 1,3-linked side chains from 1,3,5-L-Ara*f* branch points (**Extended Data Fig. 5**)^19^.

**Fig. 2.**
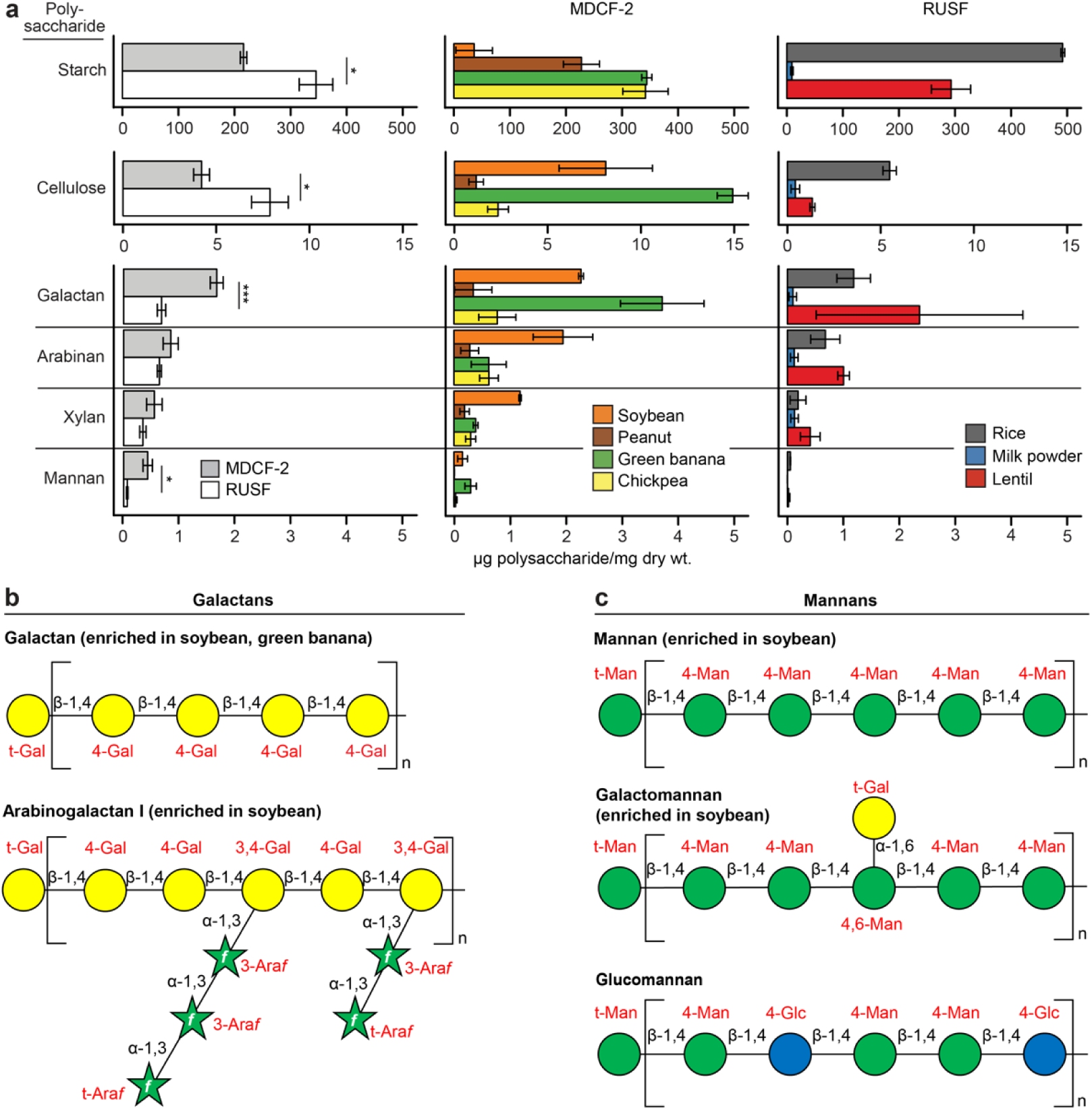
Principal polysaccharides in MDCF-2, RUSF and their component food ingredients. (**a**) Mean values ± SD are plotted. *, *P*<0.05; ***, *P*<0.001 (t-test). (**b**,**c**) Structures of galactans (panel b) and mannans (panel c) in MDCF-2.

### MDCF-2 effects on WLZ-associated MAG gene expression

Microbial RNA-Seq was performed using RNA isolated from fecal samples collected from all study participants just prior to initiation of treatment, and after 1- and 3-months of treatment (n=350 samples). Transcripts were then quantified by mapping reads from each sample to MAGs. The resulting counts tables were filtered based on the abundance and prevalence of MAGs in the full set of all fecal samples. These filtering steps were designed to exclude MAGs with minimal contributions to the meta-transcriptome from subsequent differential expression analysis (exclusion criteria were benchmarked against a simulated meta-transcriptomic dataset using the approach described in *Supplementary Methods*).

We employed principal components analysis (PCA) to determine baseline differences in overall (DNA-based) MAG abundance profiles, or the abundance of MAG-derived RNAs in the expressed meta-transcriptomes, between the treatment groups, and to subsequently identify microbes that were principal drivers of shifts during treatment. **Fig 3a** plots (i) the percent variance explained by the first principal component (PC) in analyses of 837 MAGs in fecal samples collected across all time points from all study participants and (ii) taxa enriched (q<0.05; GSEA) along the first principal component of the MAG abundance and meta-transcriptome datasets (**Fig. 3a**; see **Extended Data Fig. 6** for details of analyses of additional PCs). There were no statistically significant differences in microbiome or meta-transcriptome configuration between groups prior to treatment, or between the MDCF-2 and RUSF groups at each study week (*P*>0.1; PERMANOVA). Analysis of MAG contributions to each PCA highlights the remarkable enrichment of *Prevotella spp.* transcripts, and to a lesser extent, *Bifidobacterium spp.* transcripts, along the principal axis of variation (PC1) of the RNA-based PCA, and to a much lesser degree, the enrichment of these organisms along PC1 of the DNA-based MAG abundance PCA (**Supplementary Table 10**).

**Fig. 3.**
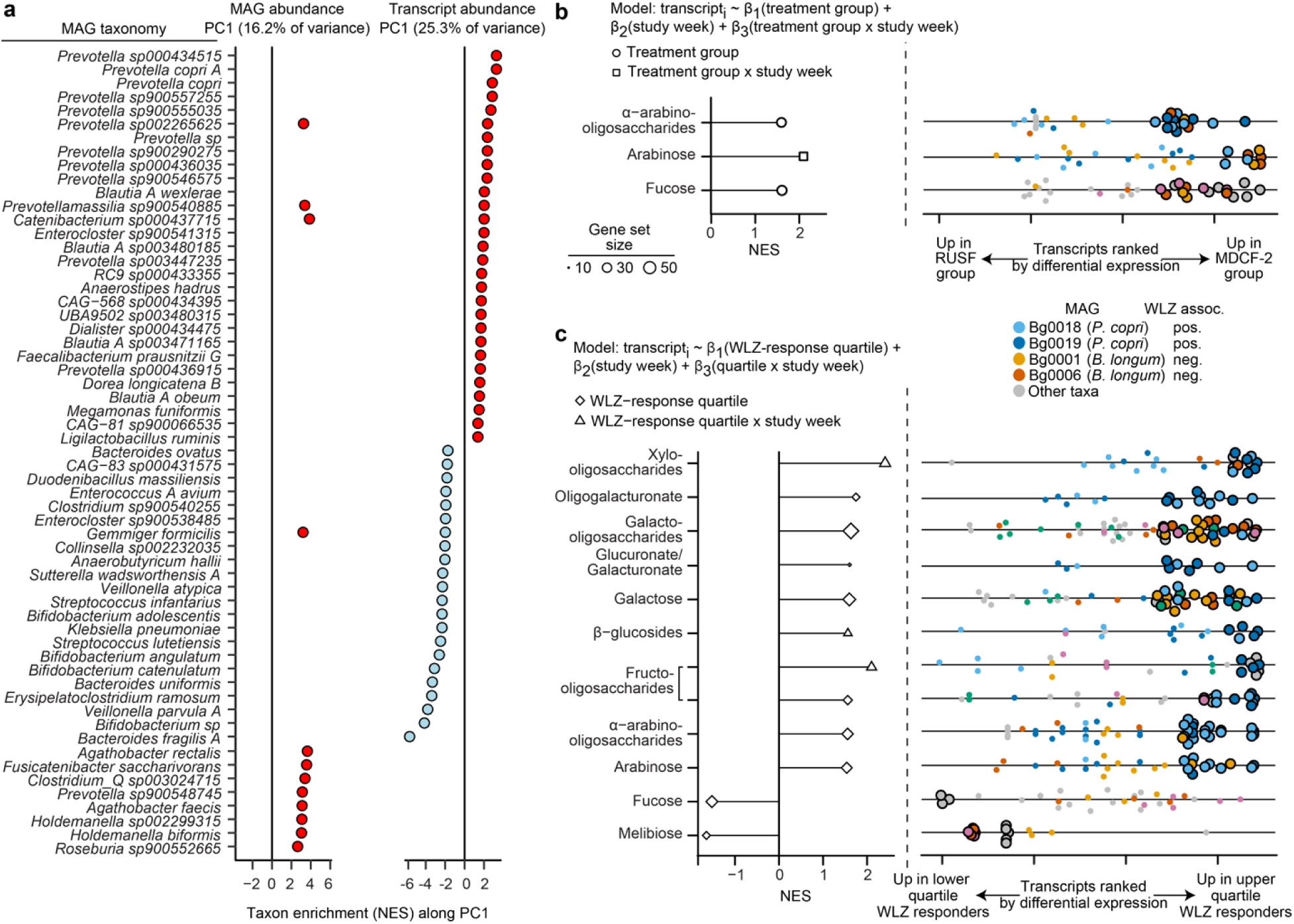
Principal taxonomic features and expressed functions of MDCF-2 and RUSF-treated fecal microbiomes. (**a**) Significant enrichment of taxa (q<0.1; GSEA) along the first principal component (PC1) of MAG abundance or transcript abundance. Abbreviation: NES; Normalized enrichment score. (**b**) Carbohydrate utilization pathways significantly enriched (q<0.1; GSEA) by treatment group (β_1_, circles) or the interaction of treatment group and study week (β_3_, squares). Right subpanel: Each point represents a MAG transcript assigned to each of the indicated functional pathways (rows), ranked by the direction and statistical significance of their differential expression in MDCF-2 versus RUSF treated participants (defined as the direction of the fold-change × -log_10_(*P*-value)). Transcripts are colored by their MAGs of origin. Larger, black outlined circles indicate leading edge transcripts assigned to the pathway described at the left of the panel. (**c**) Carbohydrate utilization pathways significantly enriched (q<0.1; GSEA) in upper vs lower WLZ quartile responders (β_1_, diamonds), or the interaction of WLZ response quartile and study week (β_3_, triangles) (see linear mixed effects model). Right subpanel: Transcripts assigned to each functional pathway. The coloring and outlining of circles have identical meaning as in panel b. The enrichment of glucuronate and galacturonate pathways was driven by the same transcripts; hence, these pathways were considered as a single unit. See **Supplementary Tables 10-14** for supporting information.

We subsequently focused on transcripts expressed by the 222 MAGs whose abundances were significantly associated with WLZ. Transcripts were ranked by their response to MDCF-2 versus RUSF treatment or by their response over time (negative binomial generalized linear model; see equation in **Fig. 3b**). GSEA was then performed to identify metabolic pathways enriched in these ranked transcripts. The analysis revealed an MDCF-2-associated pattern of gene expression characterized by significant enrichment (q<0.1; GSEA) of three metabolic pathways related to carbohydrate utilization [α-arabinooligosaccharide (aAOS), arabinose and fucose; **Fig. 3b**], three pathways related to *de novo* amino acid synthesis (arginine, glutamine, and lysine biosynthesis), and one pathway for *de novo* vitamin synthesis (folate; **Supplementary Tables 11** and **12**). In contrast, none of the 106 metabolic pathways exhibited statistically significant enrichment in their expression in children who received RUSF.

We next investigated which MAGs were responsible for the observed enrichment of expressed pathways. To do so, we turned to ‘leading edge transcripts,’ a term defined by GSEA as those transcripts responsible for enrichment of a given pathway (*Methods*). Among positively WLZ-associated MAGs, two belonging to *P. copri* (MAG Bg0018 and MAG Bg0019) were the source of 11 of the 14 leading-edge transcripts related to aAOS utilization (**Supplementary Table 12**) – a pathway whose expression was significantly elevated in children treated with MDCF-2 compared to RUSF (**Fig. 3b)**. Of the 11 *P*. copri MAGs in our dataset, these two were the only MAGs assigned to this species whose abundances were significantly positively correlated with WLZ. Both MAGs are members of a *P. copri* clade (Clade ‘A’) that is broadly distributed geographically^20,21^ (**Extended Data Fig.7a**; see **Extended Data Fig. 7b** for the predicted carbohydrate utilization pathways represented in all 51 MAGs assigned to the genus *Prevotella* that were identified in our 1,000 MAG dataset).

Although *P. copri* MAGs were the greatest source of leading-edge transcripts related to arabinose and aAOS utilization, other MAGs in the microbiome display expression responses consistent with their participation in metabolizing MDCF-2 glycans (or their breakdown products); these include MAGs that are negatively correlated with WLZ. For example, leading-edge transcripts assigned to aAOS, arabinose and fucose utilization also arose from MAGs assigned to *Bifidobacterium longum* subsp. *longum* (Bg0006), *Bifidobacterium longum* subsp. *suis* (Bg0001), *Bifidobacterium breve* (Bg0010; Bg0014), *Bifidobacterium* sp. (Bg0070), and *Ruminococcus gnavus* (Bg0067) (**Supplementary Table 12**). Features of the metabolism of these glycans in *Bifidobacterium* and *Ruminococcus* MAGs are distinct from those expressed by the *P. copri* MAGs. For example, *B. longum* subsp. *longum* MAG Bg0006 encodes an extracellular exo-α-1,3-arabinofuranosidase (BlArafA) that belongs to glycoside hydrolase (GH) family 43_22; this enzyme cleaves terminal 1,3-linked-L-Araf residues present at the ends of branched arabinans and arabinogalactans, two abundant glycans found in MDCF-2 (**Fig. 2b**; **Extended Data Fig. 5c**)^22,23^. In contrast, *P. copri* possesses an endo-α-1,5-L-arabinanase that cleaves interior α-1,5-L-Ara*f* linkages, generating aAOS. Integrating these predictions suggests a complex set of interactions between primary arabinan degraders like *P. copri* and members of *B. longum*, such as Bg0001 and Bg0006, that are capable of metabolizing products of arabinan degradation (see **Extended Data Fig. 8** for reconstructions of carbohydrate utilization pathways in *Bifidobacterium* MAGs). We cannot discern whether the arabinose available to *Bifidobacterium* is derived from free arabinose or the breakdown products of arabinan polysaccharides. It is important to consider that in these 12- to 18-month-old children with MAM, responses to MDCF-2 are occurring in the context of the underlying co-development of their microbial community and host biology, during the period of transition from exclusive milk feeding to a fully weaned state. A MAG defined as positively associated with WLZ by linear modeling is an organism whose fitness (abundance) increases as WLZ increases. Our studies in healthy 1- to 24-month-old children living in Mirpur have documented how *B. longum* and other members of *Bifidobacterium* decrease in absolute abundance during the period of complementary feeding^24^. For the negatively WLZ-associated *Bifidobacterium* MAGs described above, the levels of consumption of MDCF-2 metabolic products during the period of complementary feeding may not be sufficient to overcome a more dominant effect exerted on their abundance/fitness by the state of community-host co-development. Moreover, the metabolic capacities of *B. longum* including, as well as beyond, those related to processing of MDCF-2 glycans, may influence host growth despite *B. longum* being naturally depleted over developmental time.

Based on these observations, we sought further evidence that the two *P. copri* MAGs are related to the magnitude of ponderal growth responses and to levels of fecal glycan structures generated from MDCF-2 metabolism.

### Bacterial carbohydrate utilization pathways and clinical responses

*PUL conservation in P. copri MAGs* - As noted above, the primary outcome measure of the clinical trial was the rate of change of WLZ over the 3-month intervention. We stratified participants receiving MDCF-2 into WLZ-response quartiles^4^ and focused our analysis on (i) children in the upper and lower WLZ quartiles (n=15/group) and (ii) transcripts expressed by the 222 MAGs whose abundances were significantly associated with WLZ. We tested for enrichment of carbohydrate utilization pathways in transcripts rank-ordered by the strength and direction of their relationship with WLZ response quartile or, in a separate analysis, the interaction between WLZ response quartile and study week; we then performed GSEA to identify enriched pathways (**Supplementary Tables 13** and **14).** Eight carbohydrate utilization pathways were significantly enriched in transcripts differentially expressed in upper compared to lower WLZ quartile responders. One of these pathways (fructooligosaccharides utilization), plus three other pathways that are involved in arabinose, β-glucoside, and xylooligosaccharide utilization, were enriched in transcripts with a positive ‘WLZ quartile x study week’ interaction coefficient (β_3_), suggesting that the extent of the difference in expression of these pathways increases over the course of treatment (**Fig. 3c**, **Supplementary Table 14a**; see **Supplementary Table 14b** for enrichment of expressed vitamin and amino acid biosynthetic pathways related to WLZ-response quartile).

Remarkably, over half of the leading-edge transcripts (67/99; 68%) from the eight, upper WLZ quartile enriched carbohydrate utilization pathways were expressed by *P. copri* MAGs Bg0018 and Bg0019. Moreover, these two MAGs contributed no leading-edge transcripts to lower WLZ-response quartile enriched pathways.

*P. copri* is a member of the phylum Bacteroidota. Members of this phylum contain syntenic sets of genes known as polysaccharide utilization loci (PULs) that mediate detection, import and metabolism of a specific glycan or set of glycans^25^. To further define how expressed genomic features distinguish the capacity of MAGs Bg0018 and Bg0019 to respond to MDCF-2, we identified PULs in these MAGs and compared them to PULs present in the nine other *P. copri* MAGs in this study. These two WLZ-associated *P. copri* MAGs share (i) seven PULs we designated as conserved (*i.e*., pairwise comparisons of ORFs satisfy the requirements that their protein products have >90% amino acid identity and are organized in an identical way within the respective genomes) plus (ii) three PULs designated as present but ‘structurally distinct’ (*i.e*., a given PUL is present in the genomes being compared but component CAZymes or SusC/SusD proteins are missing or fragmented in a way likely to impact their function, or where extra ORFs are present; see *Supplementary Methods*). The representation of these 10 PULs varied among the other nine *P. copri* MAGs which span three of the four principal clades of this organism (**Fig. 4a**; **Supplementary Table 15**). Strikingly, the degree of genomic conservation of these PULs is significantly associated with the strength of WLZ association for each of the 11 *P. copri* MAGs in our MAG dataset across both treatment groups [Pearson r between Euclidean distance from Bg0019 PUL profile and β_1_(MAG) = -0.79 (*P* = 0.0035); **Fig. 4b**, see **Supplementary Table 3** for WLZ associations]. Five of the seven highly conserved PULs are related to utilization of mannan and galactan – glycans that are significantly more abundant in MDCF-2 than RUSF. Expression of three of these seven PULs, as well as two of the conserved but structurally distinct PULs, is also related to the enrichment of transcripts in carbohydrate utilization pathways that distinguish upper from lower WLZ-quartile responders (‘WLZ-response quartile’ or ‘WLZ quartile x study week’ terms in **Fig. 3c**). PULs that generate these leading-edge transcripts are predicted to metabolize β-glucan, glucomannan, β-mannan, xylan, pectin/pectic galactan and arabinogalactan (see **Fig. 4a** for which of these 10 PULs contribute differentially expressed transcripts, plus **Supplementary Table 15**).

**Fig. 4.**
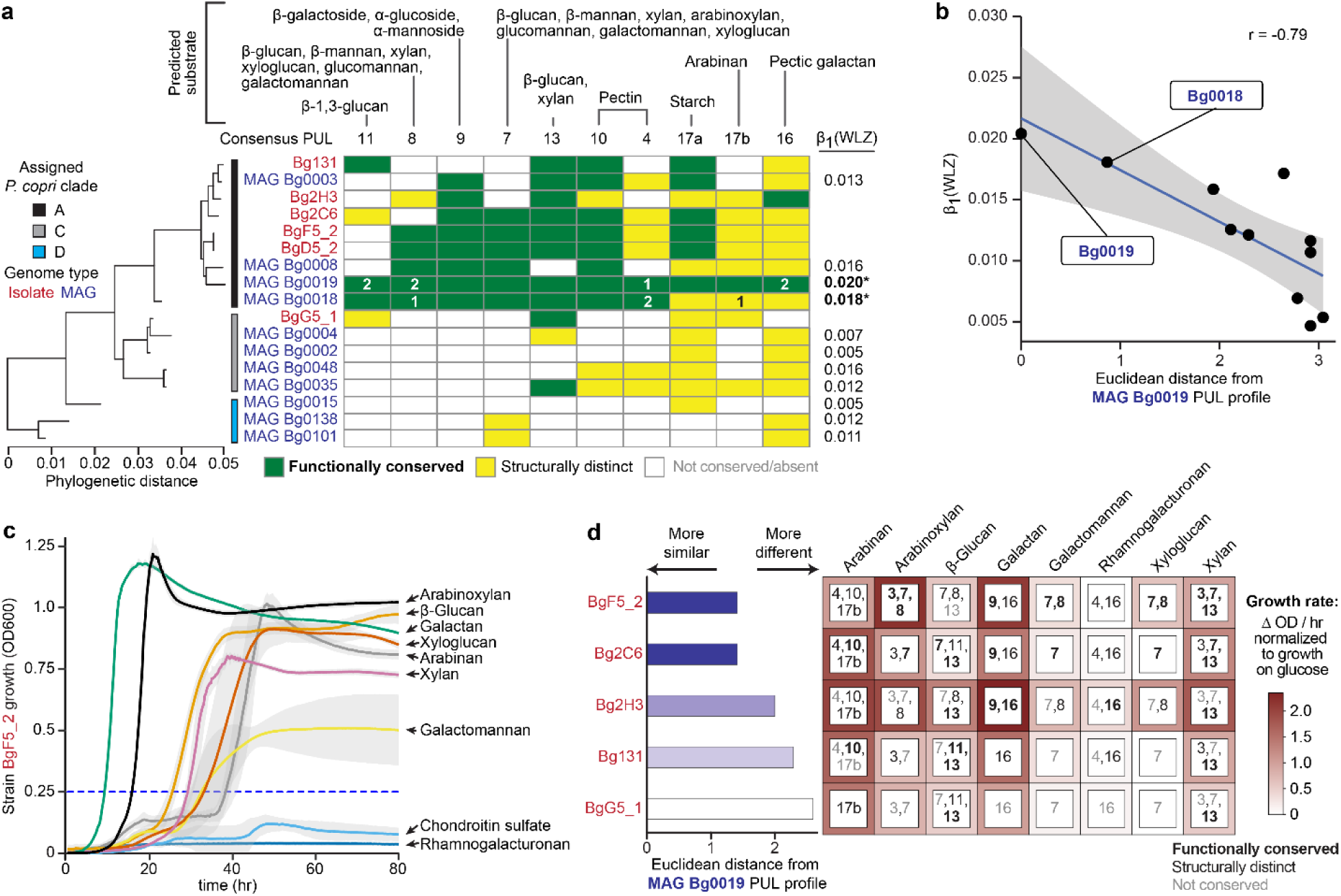
Conservation and expression of Polysaccharide Utilization Loci (PULs) in *P. copri* MAGs and isolates. (**a**) PUL conservation in *P. copri* MAGs identified in study participants (blue font) and in *P. copri* isolates cultured from the fecal microbiota of Bangladeshi children (red font). The phylogenetic tree (left) indicates the relatedness of *P. copri* MAGs and isolates as determined by a marker gene-based phylogenetic analysis (see *Methods* and **Extended Data Fig. 7**). *P. copri* clade designations are indicated by colored vertical bars. The β_1_(WLZ) coefficient for each *P. copri* MAG is indicated on the right of the panel; significant associations (q<0.05) are bolded. The color-coded matrix in the center indicates the extent of conservation of PULs in Bg0019 and Bg0018 versus the other *P. copri* MAGs and cultured isolates. The known or predicted polysaccharide substrates of these PULs are noted. Since MAG Bg0019 displayed a stronger association with WLZ than the other significantly WLZ-associated *P. copri* MAG Bg0018, it was used as a reference for the comparative PUL analysis. The number of leading-edge differentially expressed PUL transcripts in MAG Bg0018 and Bg0019 are shown within the colored cells; they were identified based on analysis of the fecal meta-transcriptomes of MDCF-2 versus RUSF treated participants, and/or from MDCF-2 treated participants in the upper-versus lower-WLZ quartiles (see **Supplementary Table 15** for PUL numbering convention and for annotations of leading-edge transcripts). (**b**) Relationship between PUL conservation in the 11 *P. copri* MAGs identified in study participants and the strength of each MAG’s association with WLZ. (**c,d**) *In vitro* growth assays for five *P. copri* isolates in defined medium supplemented with individual purified glycans representative of those in MDCF-2. Panel c shows growth curves for *P. copri* BgF5_2, the isolate whose PUL profile is most similar to MAGs Bg0019/Bg0018. Data represent mean OD_600_ measurements with standard deviations indicated as grey ribbons (n=3 replicates/condition). Panel d summarizes PUL conservation as well as the growth rates for each of the five *P. copri* strains tested (panel c and **Extended Data Fig. 9a**). Each row represents a given strain. Each colored box represents PULs in that organism predicted to use the carbohydrate tested. PULs are noted as ‘functionally conserved’ (black, bold font), ‘structurally distinct’ but functionally similar (black, not bolded) or ‘not conserved’ (grey) according to the scheme shown in panel a. The color intensity surrounding each box indicates the mean maximum growth rate for each isolate in the presence of each glycan.

A comparative analysis of MAGs Bg0018 and Bg0019 and 22 reference *P. copri* genomes in PULDB^26^ indicated that one of the highly conserved PULs (PUL7) contains a bimodular GH26|GH5_4 β-glycanase with 52% amino acid sequence identity to an enzyme known to cleave β-glucan, β-mannan, xylan, arabinoxylan, glucomannan, and xyloglucan (**Fig. 4a**, **Supplementary Table 15**)^27,28^. The gene encoding this multifunctional enzyme did not satisfy our criteria for statistically significant differential expression between MDCF-2 and RUSF treatment, nor between upper versus lower quartile WLZ-responders. However, it was consistently expressed across these conditions/comparisons (**Supplementary Table 15**) and its enzymatic product is expected to contribute to the utilization of a broad range of plant glycans, including those represented in MDCF-2.

Together, these results highlight both the versatility in carbohydrate metabolic capabilities of these two WLZ-associated *P. copri* MAGs, as well as the specificity of their treatment-inducible metabolic pathways for carbohydrates prominently represented in MDCF-2.

#### Effects of different carbohydrates on in vitro growth of cultured representatives of MAGs Bg0018 and Bg0019

To contextualize our observations regarding conserved polysaccharide degradation features of *P. copri* MAGs, we engaged in an extensive effort to culture and characterize representatives of these MAGs from fecal samples obtained from participants in this clinical trial, plus a previous, shorter duration pilot study of MDCF prototypes^3^. Based on this effort, we selected a set of six *P. copri* isolates that represented diverse repertoires of conserved PULs as well as a range of phylogenetic distances from the WLZ-associated MAGs Bg0018 and Bg0019 (**Fig. 4a; Supplementary Table 1b**). Strains BgD5_2 and BgF5_2 are highly related phylogenetically to each other and to MAGs Bg0018 and Bg0019. Notably, they possess 9 of the 10 conserved PULs in these MAGs (see **Supplementary Table 7c** and **Supplementary Table 15b** for more details of functional conservation between the genomes of these and the other cultured *P. copri* strains and MAGs). Based on the substrate predictions for each conserved PUL, the measured glycan components of MDCF-2, and the variation in conservation of these PULs across our *P. copri* MAGs and isolates, we selected eight candidate glycan substrates for *in vitro* screening: sugar beet arabinan, wheat arabinoxylan, barley β-glucan, potato galactan, carob galactomannan, soybean rhamnogalacturonan, tamarind xyloglucan, and beechwood xylan (**Supplementary Table 16a**). Chondroitin sulfate was included in the panel as a negative control given its resistance to degradation by *P. copri*^29^. Each cultured isolate was grown in a defined medium containing 1% (w/v) of each glycan as the sole carbon source, and growth was determined by tracking optical density over time (**Fig. 4c, Extended Data Fig. 9a**). Strain BgD5_2 displayed poor and inconsistent growth in this medium compared to BgF5_2, even when glucose was used as the sole carbon source; therefore, the BgD5_2 isolate was not included in these *in vitro* experiments. The results underscore the broad glycan utilization capabilities of the *P. copri* isolates but also highlight their distinct preferences for individual glycans. **Fig. 4c,d, Extended Data Fig. 9** and **Supplementary Table 16b-d** demonstrate how their growth phenotypes are aligned with their PUL repertoires, the known and predicted substrate specificities of the carbohydrate-active enzymes (CAZymes) encoded by their PULs, and the results of mass spectrometry-based quantification of their consumption of monosaccharide components of the tested glycans. Isolates whose PUL profiles matched the two WLZ-associated MAGs most closely (BgF5_2, Bg2C6, Bg2H3) displayed the strongest preference for glycan substrates that were enriched in and/or unique to MDCF-2 relative to RUSF, including arabinans (arabinan, arabinoxylan) and galactans/mannans (galactan, galactomannan) (**Fig. 2**; **Extended Data Fig. 5**). Notably, strain BgF5_2 displayed growth preferences for arabinoxylan and galactan; whereas all other strains favored arabinan over arabinoxylan. Together, these results support predictions of the capacities of the two WLZ-associated MAGs to utilize MDCF-2 glycans; they also indicate that BgF5_2 could be considered as a cultured representative of Bg0018 and Bg0019 given its similar glycan utilization preferences/capacities to those predicted for these two MAGs.

#### Fecal glycosidic linkage levels, WLZ responses and PUL expression

The same fecal samplescollected at the 0- and 3-month time points from participants in the upper and lower WLZ quartiles in the MDCF-2 treatment group that had been used for the DNA- and RNA-level analyses were subjected to UHPLC-QqQ-MS-based quantitation of 49 glycosidic linkages. These linkages were measured after their liberation by *in vitro* hydrolysis of fecal glycans (**Supplementary Table 17**). We used linear mixed effects modeling to compare the changes in levels of fecal glycosidic linkages from baseline/pre-intervention to the treatment endpoint (3 months) as a function of WLZ response quartile. The results demonstrated that with treatment, levels of 14 linkages increased significantly more (q<0.05) in members of the upper compared to the lower WLZ response quartile. None of the 49 linkages increased significantly more in children who were in the lower compared to upper WLZ response quartile (**Fig. 5a, Extended Data Fig. 10a, Supplementary Table 18a**). All 14 glycosidic linkages elevated in upper quartile responders are represented in MDCF-2 [e.g., 4,6-mannose, which is predicted to be a product of soybean galactomannan cleavage by endo-1,4-β-mannosidases encoded by PUL7 and PUL8 present in the two WLZ-associated MAGs (**Fig. 5d**, **Supplementary Table 9c**); also see **Fig. 5a** for the likely polysaccharide sources of these 14 linkages in MDCF-2].

**Fig. 5.**
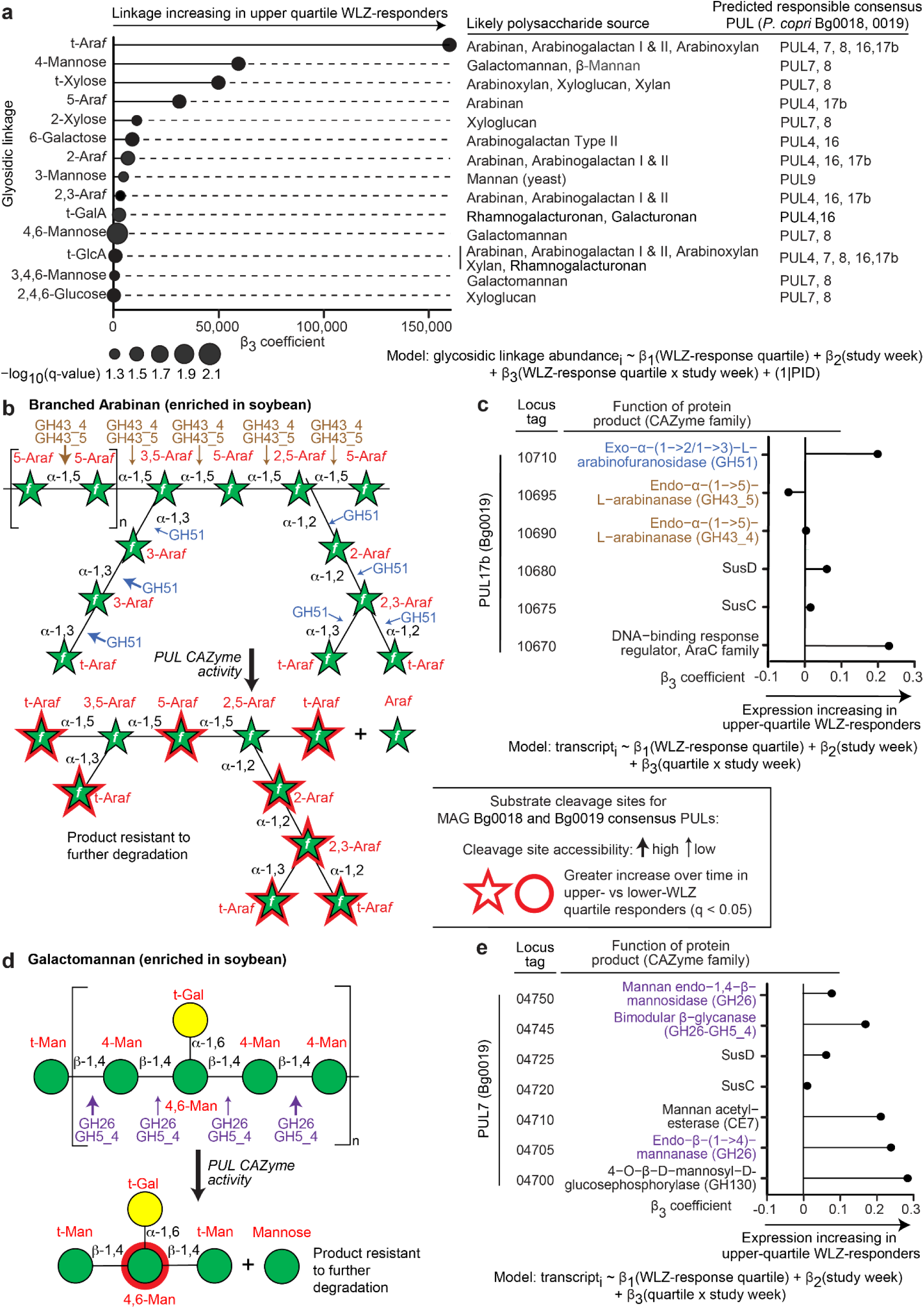
Treatment-responsive glycosidic linkages and corresponding polysaccharide sources and structures, cleavage sites, and predicted products of CAZyme activity. (**a**) Significant changes in fecal glycosidic linkage levels (q<0.05) over time in upper compared to lower WLZ quartile responders. Likely polysaccharide sources for each of the 14 glycosidic linkages are noted in the middle column (see **Extended Data Fig. 5d**). PULs present in *P. copri* MAGs Bg0018 and Bg0019 with known or predicted cleavage activity for the listed polysaccharide sources are noted in the right subpanel. (**b,d**) Structures of the MDCF-2 polysaccharides branched arabinan (panel b) and galactomannan (panel d), plus glycan fragments and their constituent glycosidic linkages predicted to be liberated by conserved *P. copri* MAGs Bg0019 and Bg0018 PULs (see Fig. 4a for results of PUL conservation analysis). Linkages highlighted with arrows are putative sites of cleavage by *P. copri* CAZymes based on their known or predicted enzyme activities; enzymes are labeled by their CAZyme module or modules predicted to perform the cleavage. The size of these arrows (large versus small) denotes the relative likelihood (high versus low, respectively) of glycosidic linkage cleavage by these CAZymes, considering steric hindrance at glycan branch points. The right subpanels illustrate the expression of PUL genes in MDCF-2 treated, upper vs lower WLZ quartile responders (only PUL genes with mcSEED or CAZy annotations are shown). (**c,e**) Predicted activities and expression of *P. copri* PUL CAZymes. Panel c depicts CAZymes assigned to PUL17b, including the GH51 family CAZyme (blue) expected to cleave α-1,2- and α-1,3-linked arabinofuranose (Ara*f*) side chains and the GH43 family CAZymes (brown), including GH43_4 and GH43_5 subfamilies predicted to cleave α-1,5-Ara*f*-linked backbone of branched arabinan, yielding products containing t-Ara*f*, 2-Ara*f*, 2,3 Ara*f* and 5-Ara*f* linkages. Panel e depicts CAZymes assigned to PUL7, including GH26 and GH5_4 CAZymes (magenta) predicted to cleave β-1,4 linked mannose residues of galactomannan, yielding products containing 4,6-mannose, the most significantly differentially abundant linkage in the upper quartile WLZ responders (see panel a).

Levels of glycosidic linkages in feces reflect a complex dynamic that includes, but is not necessarily limited to, the substrate specificities of the CAZymes encoded and expressed by PULs in primary consumers of available polysaccharides, the levels of host consumption of MDCF-2 and components of their ‘background’ diets, and the degree to which the initial products of polysaccharide degradation can be further processed by community members. These points are illustrated by the following observations. *First,* the presence of the 14 glycosidic linkages in feces can be explained in part by the specificity of CAZymes encoded and expressed by PULs conserved between *P. copri* MAGs Bg0018 and Bg0019. **Fig. 5b-e**, **Extended Data Fig. 10b and Extended Data Fig.11** describe which of their PULs are predicted to generate glycan fragments containing these linkages – predictions that are supported by the *in vitro* data generated from the cultured representative of the two MAGs. For example, t-Ara*f,* 5-Ara*f,* 2-Ara*f,* and 2,3-Ara*f* are components of polysaccharides (arabinan, arabinoxylan and arabinogalactan type I/II) present in soybean, chickpea, peanut and banana. CAZymes encoded by *P. copri* Bg0019 PULs 4, 7, 8, 16 and 17b have substrate specificities that allow them to cleave accessible linkages in these polysaccharides (see **Fig. 5b,c; Extended Data Fig. 10** and **Extended Data Fig. 11)**. Some of the products of these cleavage events are likely resistant to further degradation. The exo-α-1,2/1,3-L-arabinofuranosidase and endo-α-1,5-L-arabinanase activities encoded by PUL17b (**Fig. 5b**) are predicted to remove successive residues from the 1,2 and 1,3-linked-L-Ara*f* chains of branched arabinan and hydrolyze the 1,5-linked-L-Ara*f* backbone from this polysaccharide, yielding an enzyme-resistant product containing t-Ara*f*, 5-Ara*f*, 2-Ara*f*, and 2,3-Ara*f* linkages. *Second,* CAZyme transcripts assigned to PULs 4, 7, 8, 16 and 17b were detectable in the fecal meta-transcriptomes of all but one of the 30 participants assigned to the upper or lower WLZ responder quartiles. Levels of expression of the majority of these CAZymes genes were modestly elevated in upper compared to lower WLZ-quartile responders over the course of treatment, although the difference did not reach our threshold cutoff for statistical significance (q<0.05). These transcripts include the GH51 encoded by PUL17b plus the GH26, GH26|GH5_4, GH130 and carbohydrate esterase family 7 (CE7) transcripts from PUL7; see **Extended Data Fig. 10b**. *Third,* while intake of MDCF-2 was not significantly different between the upper and lower WLZ quartile participants (*P*>0.05; linear mixed effects model), data from a food frequency questionnaire (FFQ) administered at the time of each fecal sampling disclosed a positive correlation between consumption of legumes and nuts and the levels of t-Ara*f*, 5-Ara*f*, 2,3-Ara*f*, t-GalA, and 2,4,6-Glucose (**Supplementary Table 18b**). Consumption of these foods was also the most discriminatory response between upper compared to lower WLZ quartile responders (**Supplementary Table 18c**). These observations suggest that children consuming more of the classes of complementary food ingredients present in MDCF-2 may also exhibit enhanced growth responses.

The confounding effects of background diet and the role of *P. copri* in processing MDCF-2 glycans can be directly tested in gnotobiotic mice colonized with a defined community of cultured representatives of WLZ-associated MAGs. One such gnotobiotic model is described in our companion study where mice were colonized with a defined consortium of age- and WLZ-associated Bangladeshi bacterial strains, with or without *P. copri* isolates that captured key features of the carbohydrate metabolic apparatus present in Bg0018 and Bg0019 (such as BgF5_2/BgD5_2). Analyses revealed that these *P. copri* strains were the principal mediators of MDCF-2 glycan degradation *in vivo* and that the combination of the presence of *P. copri* and a MDCF-2 diet was associated with promotion of ponderal growth and had marked effects on multiple aspects of metabolism in intestinal epithelial cell lineages^5^.

## Discussion

The current study illustrates an approach for characterizing the gut microbiome targets and structure-function relationships of a therapeutic food - in this case, MDCF-2. MDCF-2 produced significantly greater weight gain during a 3-month, randomized controlled study of 12- to 18-month-old Bangladeshi children with moderate acute malnutrition compared to a conventional, more calorically dense RUSF. We have focused on metagenome-assembled genomes (MAGs), specifically (i) treatment-induced changes in expression of carbohydrate metabolic pathways in MAGs whose abundances were significantly associated with weight gain (WLZ), and (ii) mass spectrometric analysis of the metabolism of glycans present in the two food formulations. Quantifying monosaccharides, glycosidic linkages and polysaccharides present in MDCF-2, RUSF and their component ingredients disclosed that MDCF-2 contains more galactans and mannans (*e.g*., galactan, arabinogalactan I, galactomannan, β-mannan, glucomannan). Two types of comparisons were performed of the transcriptional responses of MAGs that were significantly associated with WLZ: one involved study participants who had consumed MDCF-2 versus RUSF, and the other focused on MDCF-2 treated children in the upper versus lower quartiles of WLZ response. The results revealed that two *P. copri* MAGs, both positively associated with WLZ, were the principal contributors to MDCF-2-induced expression of metabolic pathways involved in the utilization of its component glycans.

UHPLC-QqQ-MS was able to identify statistically significant changes in glycan composition in feces from children consuming a therapeutic food, even in the face of complex and varied background diets. Intriguingly, although intake of MDCF-2 did not differ between children in the upper and lower quartiles of clinical (WLZ) response, children in the upper quartile trended toward diets containing more legumes and nuts than those in the lower quartile. The “legumes and nuts” food group includes major components of MDCF-2. We postulate that MDCF-2 ‘kick-starts’ a microbiome response that includes changes in the fitness and expressed metabolic functions of key growth-associated bacterial strains, such as *P. copri*. Background diet can further modify this response, as evidenced by the higher levels of microbial metabolic products of legume/nut-associated glycans in the feces of children displaying upper quartile WLZ responses. This observation also suggests that further optimization of the dose of MDCF-2 may be possible; in our study, it was administered as a dietary supplement designed to provide ∼20% of the childrens’ daily energy requirements. More detailed, quantitative assessments of food consumption during future clinical studies of MDCF-2 could serve to not only facilitate design of improved formulations/doses but also to inform future recommendations regarding complementary feeding practices - recommendations that recognize the important role of the gut microbiome in the healthy growth of children.

One definition of ‘microbiome repair’ in malnourished children is a rebalancing of the representation and expressed functions of beneficial organisms so that it assumes a configuration more conducive to healthy microbiome-host co-development. Linking dietary glycans and microbial metabolism in this fashion provides a starting point for culture-based initiatives designed to retrieve isolates of these ‘effector’ taxa for use as potential probiotic agents, or if combined with key nutrients that they covet, synbiotic formulations for repairing perturbed microbiomes that are insufficiently responsive to food-based interventions alone.

Much remains to be discovered about how MDCF-2 treatment is related to weight gain/healthy growth. For example, further work is needed to clarify whether the mediators of *P. copri* effects on the host arise from direct products of its metabolism of MDCF-2 glycans, or whether products of other metabolic pathways in *P. copri*, whose activities are regulated by biotransformation of MDCF-2 glycans, are involved. Additionally, the contributions of metabolites from other community members to these effects are unknown. Furthermore, the observed relationships between strains of *P. copri* and MDCF-2 glycans does not exclude the contribution of other macro- or micronutrients to the superiority of MDCF-2 over RUSF on weight gain in our study. Direct tests of the role played by organisms such as *P. copri* in mediating microbial community and host responses to components of microbiome-targeted therapeutic foods can come from additional clinical studies of probiotic or synbiotic formulations consisting of strains closely related to WLZ-associated MAGs, such as *P. copri* BgF5_2, administered in conjunction with MDCF-2 or with its glycan components. Another approach involves ‘reverse translation’ experiments of the type illustrated in our companion study to this report^5^. This companion study uses (i) a gnotobiotic mouse model colonized with defined collections of cultured, WLZ-associated gut bacterial taxa with or without *P. copri*, (ii) single nucleus RNA-Seq and microbial RNA-Seq and (iii) UHPLC-QqQ-MS to characterize the contributions of *P. copri* to post-weaning weight gain, processing of MDCF-2 glycans, and regulation of intestinal gene expression and metabolism.

## Methods

### Collection and handling of biospecimens obtained from participants in the randomized controlled clinical study of the efficacy of MDCF-2

The human study entitled ‘Community-based Clinical Trial With Microbiota-Directed Complementary Foods (MDCFs) Made of Locally Available Food Ingredients for the Management of Children With Primary Moderate Acute Malnutrition (MAM)’, was approved by the Ethical Review Committee at the icddr,b (Protocol PR-18073; ClinicalTrials.gov identifier: NCT04015999)^4^. Informed consent was obtained for all participants. The objective of the study was to determine whether twice daily, controlled administration of a locally produced, microbiota-directed complementary food (MDCF-2^3,4^) for 3 months to children with MAM provided superior improvements in weight gain, microbiota repair, and improvements in the levels of key plasma biomarkers/mediators of healthy growth, compared to a commonly used rice- and lentil-based ready-to-use supplementary food (RUSF) formulation.

A total of 124 male and female children with MAM (WLZ -2 to -3) between 12- and 18-months-old who satisfied the inclusion criteria were enrolled, with 62 children randomly assigned to each treatment group using the permuted block randomization method. Children in each treatment group were fed their assigned dietary supplement (MDCF-2 or RUSF) twice daily at a study center for the first month, once daily at a study center and once daily at home for the second month, and twice daily at home for the third month. Mothers were otherwise encouraged to practice their customary breast- and complementary-feeding practices. At the end of the intervention period, children returned to their normal feeding routine with continued intensive monitoring for one additional month. Fifty-nine participants in each treatment group completed the 3-month intervention and 1-month post-treatment follow-up^4^.

To minimize the risk of degradation of fecal DNA/RNA, fecal samples were collected within 20 minutes of their production and immediately transferred to liquid nitrogen-charged vapor shippers for transport to a -80 °C freezer at the study center. Specimens were shipped to Washington University on dry ice where they were stored at -80 °C in a dedicated repository with approval from the Washington University Human Research Protection Office.

### Defining the relationship between MAG abundances and WLZ

Procedures used for shotgun sequencing of fecal DNA and preprocessing of the resulting reads, plus MAG assembly and quantitation are described in *Supplementary Methods*.

Linear mixed effects models were used to relate the abundances of MAGs identified in each trial participant to WLZ using the formula:

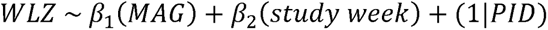

Our data normalization strategies prior to linear modeling did not include a consideration of MAG assembly length. Therefore, we analyzed the TPM (reads per kilobase per million) output of kallisto (v0.43.0) by applying a filter requiring each MAG’s abundance to be >5 TPM in >40% of the 707 fecal samples collected at time points where anthropometry was also measured. This filtering approach yielded 837 MAGs. We then returned to the unfiltered count output from kallisto, performed a variance stabilizing transformation [VST, DESeq2^30^ (v1.34.0)] to control for heteroskedasticity, and filtered the dataset to the same 837 MAGs. We subsequently fit linear mixed effects models to the transformed abundances of each MAG across all 707 fecal samples (lme4^31^, v1.1-27.1; lmerTest^32^, v3.1-3). We used ANOVA to determine the statistical significance of the fixed effects in our model – specifically, the relationship between MAG abundance and WLZ. ‘WLZ-associated MAGs’ were defined as those having *P-*values adjusted for false discovery rate (q-values) <0.05.

### Determining the effects of MDCF-2 supplementation on the abundances of WLZ-associated MAGs

We employed dream^33^ (variancePartition R package, v1.24.0) an empirical Bayesian linear mixed effects modeling framework, to model MAG abundance as a function of treatment group, study week and their interaction, controlling for the repeated measurements taken from each study participant with a random effect term for participant. The equation used to quantify the effects of treatment on MAG abundance took the form:

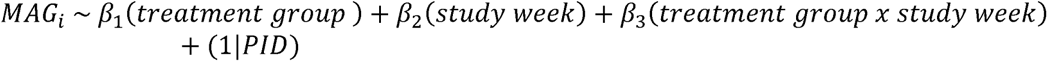

The ‘treatment group’ coefficient β_1_ indicates whether MDCF-2 produced changes in the mean abundance of a given MAG relative to RUSF over the 3-month intervention, while the ‘treatment group x study week’ interaction coefficient β_3_ indicates whether MDCF-2 affected the rate of change of a given MAG more so than RUSF (*i.e*., was a MAG increasing or decreasing more rapidly in the microbiomes of participants in the MDCF-2 versus the RUSF treatment group?). Each coefficient for each MAG abundance analysis is described by an associated t-statistic - a standardized measure, based on standard error, of a given coefficient’s deviation from zero which can be used to calculate a *P*-value and infer the significance of the effect of a given coefficient on the dependent variable. The t-statistics produced by this method can also be used as a ranking factor for input to GSEA. For this analysis, gene sets were defined as groups of MAGs that were either significantly positively (n=75) or significantly negatively (n=147) associated with WLZ. This analysis was conducted for both the ‘treatment group’ (β_1_) coefficient and the ‘treatment group x study week’ interaction (β_3_) coefficient. Statistical significance is reported as q-values after adjustment for false-discovery rate (Benjamini-Hochberg method).

### Microbial RNA-Seq analysis of MAG gene expression

For RNA extraction, approximately 50 mg of a fecal sample, collected from each participant at the baseline, 1-month and 3-month time points, was pulverized under liquid nitrogen with a mortar and pestle and aliquoted into 2 mL cryotubes. A 3.97 mm steel ball and 250 µL of 0.1 mm zirconia/silica beads were subsequently added to each sample tube, together with 500 µL of a mixture of phenol:chloroform:isoamyl alcohol (25:24:1, pH 7.8-8.2), 210 µL of 20% SDS, and 500 µL of 2X Qiagen buffer A (200 mM NaCl, 200 mM Trizma base, 20 mM EDTA). After a 1-minute treatment in a bead beater (Biospec Minibeadbeater-96), samples were centrifuged at 3,220 × g for 4 minutes at 4 °C. One hundred microliters of the resulting aqueous phase were transferred by a liquid handling robot (Tecan) to a deep 96-well plate along with 70 µL of isopropanol and 10 µL of 3M NaOAc, pH 5.5. The solution was mixed by pipetting 10 times. The crude DNA/RNA mixture was incubated at -20 °C for 1 hour and then centrifugated at 3,220 × g at 4 °C for 15 minutes before removing the supernatant to yield nucleic acid-rich pellets. A Biomek FX robot was used to add 300 µL Qiagen Buffer RLT to the pellets and to resuspend the RNA/DNA by pipetting up and down 50 times. A 400 µL aliquot was transferred from each well to an Qiagen AllPrep 96 DNA plate, which was centrifuged at 3,220 × g for 1 minute at room temperature. The RNA flow-through was purified as described in the AllPrep 96 protocol. cDNA libraries were prepared from extracted RNA using an Illumina Total RNA Prep with Ribo-Zero Plus and dual unique indexes. Libraries were balanced, pooled, and sequenced in two runs of an Illumina NovaSeq using S4 flow cells.

As an initial pre-processing step, raw reads were aggregated by sample across the two NovaSeq runs, resulting in a total of 5.0×10^7^±4.7×10^6^ paired-end 150 nt reads per sample (mean±SD). Adapter sequences and low-quality bases were removed from raw reads (Trim Galore^34^, v0.6.4), and pairs of trimmed reads were filtered out if either one of the paired reads was less than 100 nt long. Pre- and post-trimmed sequence quality and adapter contamination were assessed using FastQC^35^ (v0.11.7). Filtered reads were pseudoaligned to the set of 1,000 annotated, dereplicated high quality MAGs to quantify transcripts with kallisto^36^. Reads that pseudoaligned to rRNA genes were excluded, leaving an average of 7.1×10^6^±3.9×10^6^ bacterial mRNA reads (mean±SD) per sample. Counts tables were further filtered to retain only transcripts that pseudoaligned to the 837 MAGs that passed the abundance and prevalence thresholds described above. To minimize inconsistently quantified counts related to low-abundance MAGs, we assigned a transcript count of zero, on a per-sample basis, to any MAG with a DNA abundance <0.5 TPM in that sample.

Differential expression analysis (edgeR^37^, v3.32.1) was conducted using the following steps: (i) transcript filtering for presence/absence and prevalence; (ii) library size normalization using TMM (trimmed mean of M-values); (iii) estimating per-gene count dispersions; and (iv) testing for differentially expressed genes. Transcripts were first filtered using edgeR default parameters, followed by a parameter sweep of transcript abundance and prevalence threshold combinations. Based on this analysis, transcripts with ≥ 5 counts per million mapped reads (CPM) in ≥ 35% of samples were retained for differential expression analysis. The transcripts that passed this filtering were normalized using a TMM-based scaling factor. We next estimated negative binomial dispersions and fit trended per-gene dispersions (using the power method) to negative binomial generalized linear models. These models were used to characterize (i) the effect of treatment group and study week among all participants and (ii) the effect of WLZ quartile and study week among MDCF-2 participants in the upper and lower quartiles of WLZ response using the following model formulae:

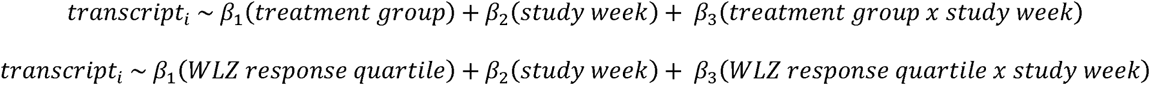

From these models, we identified genes that exhibited significant differential expression using the quasi-likelihood F-test (edgeR, function glmQLFTest) which accounts for the uncertainty in estimating the dispersion for each gene.

For subsequent functional metabolic pathway enrichment analyses, we (i) ordered transcripts assigned to WLZ-associated MAGs based on a ranking metric calculated as the direction of the fold-change × -log_10_(*P*-value) for a given differential expression analysis, (ii) defined gene sets as groups of these ranked transcripts assigned to the same metabolic pathway, and (iii) performed GSEA (fgsea^38^, v3.14). This set of analyses allowed us to identify differentially expressed metabolic pathways comprised of ≥10 genes over time (i) between treatment groups, (ii) between WLZ response quartiles or (iii) as a function of interacting terms in the linear mixed effect models (treatment group x study week; WLZ response quartile x study week). Enrichment results were considered statistically significant if they exhibited q-values<0.1 after controlling for false-discovery rate (Benjamini-Hochberg method).

For targeted transcriptional analyses of the CAZymes encoded by *P. copri* MAGs Bg0018 and Bg0019, we employed dream^33^ in R with no additional filtering, and the formula above relating transcripts to WLZ response quartile, study week, and the interaction of both terms, with the addition of a random effect for participant.

### Principal Components Analysis

Principal Components Analysis (PCA) was performed on VST-transformed DNA or transcript counts for the 837 MAGs passing the filter described in the section entitled ‘Defining the relationship between MAG abundances and WLZ’ above. The PCA performed on transcript abundances encompassed 27,518 genes expressed by these MAGs at thresholds for levels and prevalence that are described in the section entitled ‘Microbial RNA-Seq analysis of MAG gene expression’ above. PCA was performed in R using the ‘prcomp’ function, with each data type centered but not scaled since the dataset was already VST-normalized. The functions ‘get_eigenvalues,’ ‘get_pca_ind,’ and ‘get_pca_var’ from the factoextra^39^ (v1.0.7) package were utilized to extract (i) the variance explained by each principal component, (ii) the coordinates for each sample along principal components, and (iii) the contributions of each variable to principal components 1-3. We used the ‘adonis2’ function within the vegan^40^ library (v2.5-7) to test for the statistical significance of differences in the microbiome (MAGs) or meta-transcriptome between the two treatment groups at baseline or over time.

### LC-MS analyses of carbohydrates present in MDCF-2, RUSF, their component ingredients, fecal specimens and culture medium

#### Sample preparation for glycan structure analysis

Frozen samples of MDCF-2, RUSF, their respective ingredients, and fecal biospecimens were ground with a mortar and pestle while submerged in liquid nitrogen. A 50 mg aliquot of each homogenized sample was lyophilized to dryness. Lyophilized samples were shipped to the Department of Chemistry at the University of California, Davis. On receipt, samples were pulverized to a fine powder using 2 mm stainless steel beads (for foods) or 2 mm glass beads (for feces). A 10 mg/mL stock solution of each sample was prepared in Nanopure water. All stock solutions were again bead homogenized, incubated at 100 °C for 1 h, bead homogenized again, and stored at -20 °C until further analysis.

#### Monosaccharide composition analysis

Methods were adapted from previous publications^41,42^. For analyses of food ingredients and fecal biospecimens, 10 µL aliquots were withdrawn from homogenized stock solutions and transferred to a 96-well plate containing 2 mL wells. For analyses of monocultures of *P. copri* strains grown in the presence of different purified polysaccharides, microplates were withdrawn from anaerobic chamber at the conclusion of the incubation and centrifuged (5,000 x g for 5 minutes). The resulting supernatants were removed and immediately frozen at -80 °C.

Each sample was subjected to acid hydrolysis (4 M trifluoroacetic acid for 1 h at 121 °C) followed by addition of 855 µL of ice-cold Nanopure water. Hydrolyzed samples, plus an external calibration standard comprised of 14 monosaccharides with known concentrations (0.001-100 µg/mL each) were derivatized with 0.2 M 1-phenyl-3-methyl-5-pyrazolone (PMP) in methanol plus 28% NH_4_OH for 30 minutes at 70 °C. The derivatized glycosides were fully dried by vacuum centrifugation, reconstituted in Nanopure water (Thermo Fischer Scientific), and excess PMP was extracted with chloroform. A 1 µL aliquot of the aqueous layer was injected into an Agilent 1290 Infinity II ultrahigh-performance liquid chromatography (UHPLC) system, separated using a 2-minute isocratic elution on a C18 column (Poroshell HPH, 2.1 × 50 mm, 1.9 μm particle size, Agilent Technologies), and analyzed using an Agilent 6495A triple quadrupole mass spectrometer (QqQ-MS) operated in dynamic multiple reaction monitoring (dMRM) mode. Monosaccharides in the food and fecal samples were identified and quantified by comparison to the external calibration curve.

#### Glycosidic linkage analysis

Methods were adapted from a previous publication with modifications^43,44^. Under an argon atmosphere, a 5 µL aliquot from each homogenized stock solution of a sample was permethylated in a 200 µL reaction that contained 5 µL saturated NaOH and 40 µL iodomethane in 150 µL of DMSO. Permethylated glycosides were extracted with dichloromethane, and the extract was dried by vacuum centrifugation. The extracted glycosides were subjected to acid hydrolysis (4 M trifluoroacetic acid for 2 h at 100 □) followed by vacuum centrifugation to dryness. Samples were then derivatized with PMP as described above for monosaccharide analysis, followed by another vacuum centrifugation to complete dryness. Methylated monosaccharides were then reconstituted with 100 µL of 70% methanol in water. A 1 µL aliquot of the aqueous layer was injected into an Agilent 1290 Infinity II UHPLC system, separated using a 16-minute gradient elution on a C18 column (ZORBAX RRHD Eclipse Plus, 2.1 × 150 mm, 1.8 μm particle size, Agilent Technologies), and analyzed using an Agilent 6495A QqQ-MS operated in multiple reaction monitoring (MRM) mode. A standard pool of oligosaccharides and a reference MRM library were used to identify and quantify glycosidic linkages in all samples.

#### Fenton’s initiation toward defined oligosaccharide groups (FITDOG) polysaccharide analysis

Methods were adapted from previous publications^18,45^. To separate endogenous oligosaccharides from the background food matrix, polysaccharides were precipitated with 80% aqueous ethanol. Dried precipitates were reconstituted, homogenized, and 10 mg/mL stock solutions were prepared. The FITDOG reaction was carried out using a 100 μL aliquot of the 10 mg/mL resuspended food pellet and 900 μL of reaction buffer (44 mM sodium acetate, 1.5% H_2_O_2_, 73 µM Fe_2_(SO_4_)_3_(H_2_O)_5_). The reaction mixture was incubated at 100 °C for 45 minutes, quenched with 500 μL 2 M NaOH, and then neutralized with 61 μL of glacial acetic acid. The resulting oligosaccharides were then reduced to their corresponding alditols with sodium borohydride (NaBH_4_) to prevent anomerization during chromatographic separation. For the reduction of oligosaccharides, a 400 μL aliquot of the reaction mixture was incubated with 400 μL 1 M NaBH_4_ at 65 °C for 60 minutes. Oligosaccharide products were then enriched using C18 and porous graphitized carbon (PGC) 96-well solid-phase extraction plates. For the C18 enrichment, cartridges were primed with 2 × 250 μL acetonitrile (ACN) and then 5 × 250 μL water washes prior to loading the reduced sample. Cartridge effluent was collected and subjected to subsequent PGC clean-up. PGC cartridges were primed with 400 μL water, 400 μL 80% ACN/0.1% (v/v) trifluoroacetic acid (TFA), and then 400 μL water prior to loading the C18 effluent. Washing was performed with 8 × 400 μL water, and the oligosaccharides were eluted with 40% ACN/0.05% (v/v) TFA and then dried using a vacuum centrifugal dryer. Oligosaccharides were reconstituted with 100 μL Nanopure water and a 10 μL aliquot was injected into the HPLC-Q-TOF instrument. Separation was carried out using an Agilent 1260 Infinity II HPLC with a PGC column (Hypercarb, 1 × 150 mm, 5 μm particle size, Thermo Scientific) coupled to an Agilent 6530 Accurate-Mass Q-TOF mass spectrometer. Specific HPLC, electrospray source, and MS acquisition parameters are described in greater detail in previous publications^18,45^. Oligosaccharide identification was based on MS/MS fragmentation and retention time (RT) compared to reacted polysaccharide standards (amylose, cellulose, mannan, galactan, linear arabinan, and xylan). Food polysaccharides were quantified using an external calibration curve that included the three most abundant oligosaccharides from each parent polysaccharide as the quantifier species.

#### Statistical analysis of carbohydrate composition

We analyzed the abundances of glycosidic linkages over time and between WLZ-response quartiles using linear mixed effects models (lme4^31^, lmerTest^32^ packages in R) of the following form:

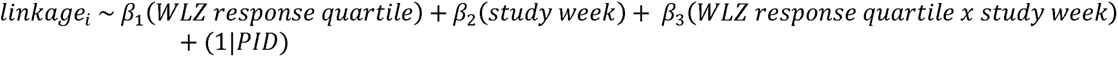

Linkages displaying a significant interaction (q <0.05) between WLZ response quartile and study week (β_3_ coefficient) were identified.

## Data Availability

Shotgun DNA sequencing and microbial RNA-Seq datasets generated from fecal samples, plus annotated P. copri isolate genome sequences are available in the European Nucleotide Archive (accession PRJEB45356). LC-MS datasets of monosaccharide, glycoside linkage and polysaccharide data are deposited in GlycoPOST (accession GPST000244).

https://www.ebi.ac.uk/ena/browser/view/PRJEB45356

## Acknowledgements

We thank Martin Meier, Justin Serugo, Su Deng, Kazi Ahsan, Jessica Hoisington-Lopez, and MariaLynn Crosby for superb technical assistance, members of the Genome Technology Access Core at Washington University School of Medicine for Illumina NovaSeq-based generation of fecal DNA shotgun sequencing and microbial RNA-Seq datasets, plus Eric Martin and Brian Koebbe for high-performance computing support. This work was supported by the Bill & Melinda Gates Foundation, the National Institutes of Health (DK30292) and the Washington University-Centene Personalized Medicine Initiative.

## Author Contributions

M.C.H. and D.M.W. generated short-read and long-read shotgun sequencing datasets of fecal DNAs and, together with H.M.L. assembled MAGs. D.R., A.O., M.K., S.A.L., and A.A. performed metabolic pathway reconstruction and annotation of MAGs, while S.H., V.L., N.T. and B.H. annotated PULs and CAZymes in MAGs. M.C.H. identified WLZ-correlated MAGs and conducted analyses of the enrichment of their encoded functions. D.M.W. generated and together with M.C.H, C.Z., D.R., A.O. and A.A. analyzed microbial RNA-Seq datasets. J.J.C., C.B.L., G.C., N.P.B., Y.C. performed LC-MS analyses of food and fecal glycans. J.L-G. isolated *P. copri* strains with assistance from D.M.W. and M.C.H. M.C.H., D.M.W. and S.H. designed the *in vitro* growth experiments, which were performed by M.C.H., D.M.W., and S.H. with assistance from J.L-G. Y.W., H.W.C., and E.L. integrated the results of gnotobiotic mouse experiments described in the accompanying paper with the MAG-based analysis described in this paper. I.M., S.D., and M.M., under the supervision of T.A., conducted the clinical study of MDCF-2 and RUSF and supplied all biospecimens used for the analyses conducted in this report. M.J.B. oversaw databases of clinical metadata and the biospecimen archive. T.A. and J.I.G. oversaw this research. M.C.H., D.M.W. and J.I.G. wrote the paper with invaluable input from co-authors.

## Competing Interests

A.O. and D.R. are co-founders of Phenobiome Inc., a company pursuing development of computational tools for predictive phenotype profiling of microbial communities. C.B.L. is a co-founder of Infinant Health, interVenn Bio, and BCD Bioscience - companies involved in the characterization of glycans and developing carbohydrate applications for human health.

Supplementary information is available for this paper.

Correspondence and requests for materials should be addressed to Jeffrey Gordon.

Reprints and permissions information is available at www.nature.com/reprints.

## Statistical Information

Statistical analyses were conducted using the approaches described in *Methods* and the figure legends. Samples sizes are indicated along with each statistic test. All relevant statistical tests are two-tailed unless otherwise specified. All measurements were collected from distinct samples. Technical replicates were not collected and analyzed unless otherwise noted.

## Extended Data

**Extended Data Fig. 1.**
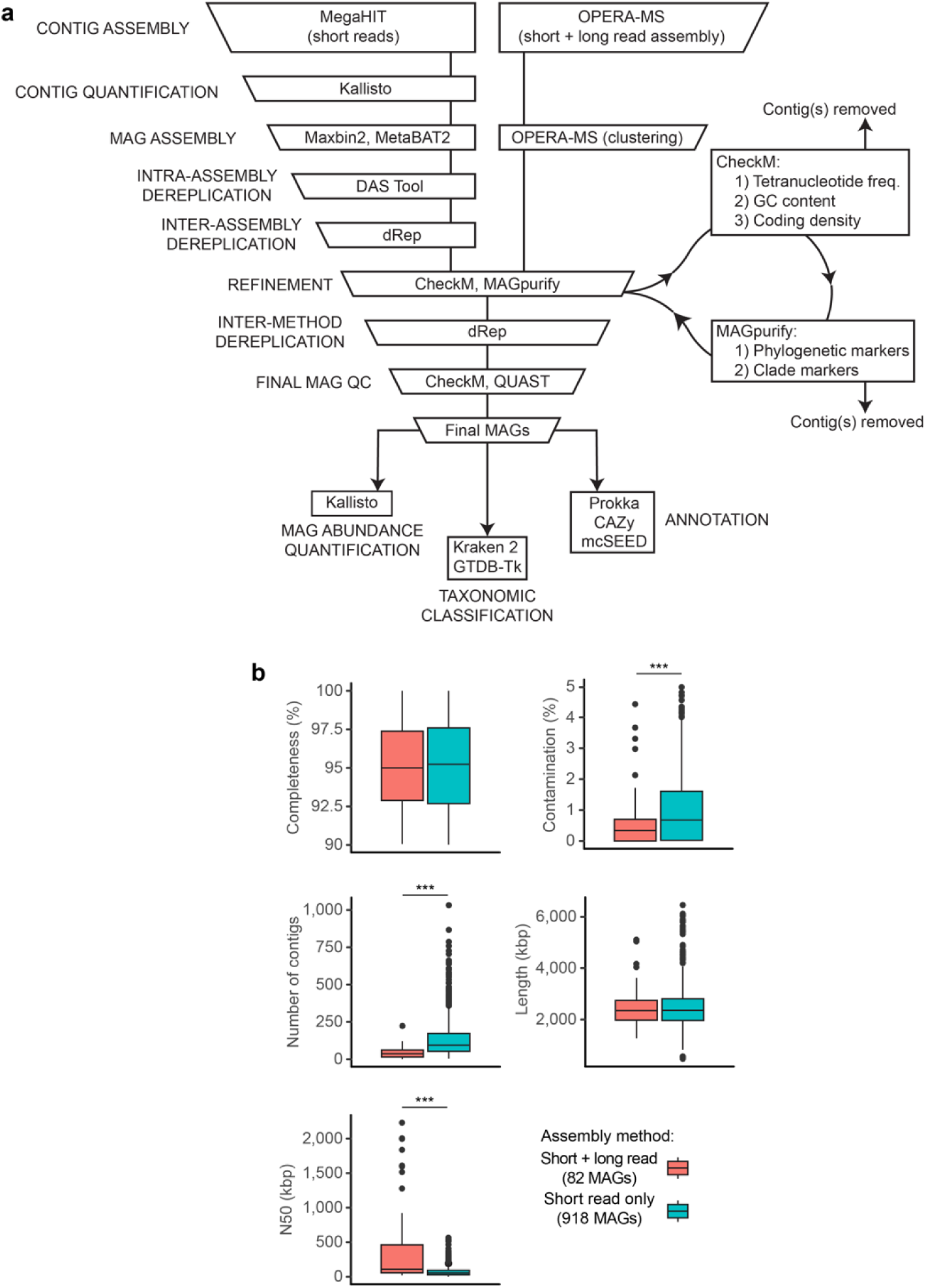
Bioinformatic workflow for MAG assembly, refinement, and quantitation. **(a)** Pipeline for MAG assembly from short-read only or short-read plus long-read shotgun sequencing data. Steps are indicated on the left while the bioinformatic tools employed to accomplish each step are described within each box. **(b)** Comparison of MAG assembly summary statistics derived from CheckM (completeness, contamination) or Quast (number, length and N50 of contigs) for 82 high-quality MAGs obtained from short-plus long-read hybrid assemblies versus 918 high-quality MAGs from short-read only assembly methods. Boxplots show the median, first and third quartiles; whiskers extend to the largest value no further than 1.5 × the interquartile range. ***, *P* < 0.001 (Wilcoxon test).

**Extended Data Fig. 2.**
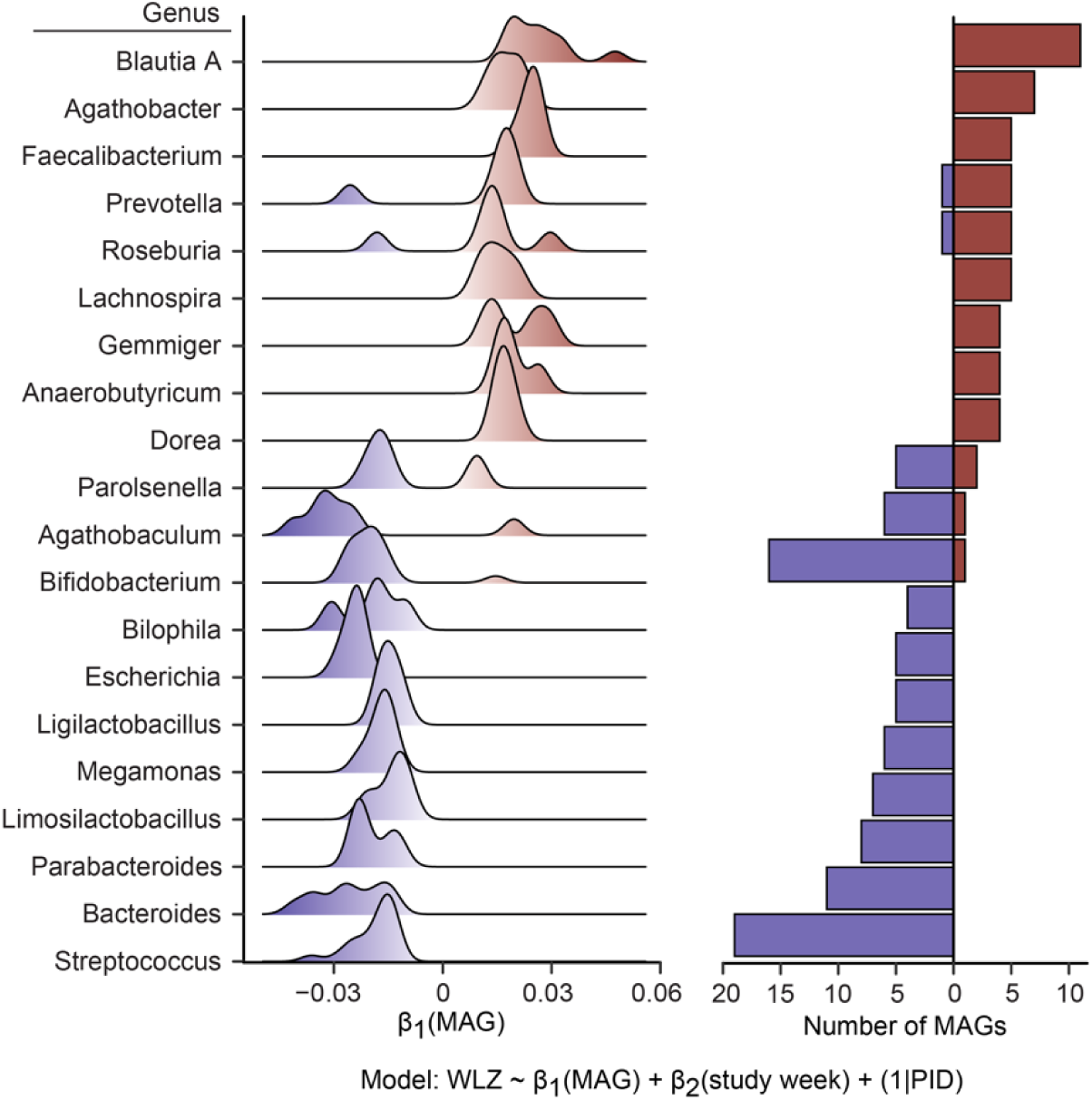
Distribution of WLZ-associated MAGs across taxonomic groups. Left subpanel, density plot showing WLZ-associated MAGs tabulated based on their genus-level classification. β_1_ refers to the coefficient in the linear mixed effects model presented at the bottom of the figure. Genera containing >3 significantly WLZ-associated MAGs are shown. Right subpanel, number of statistically significant WLZ-associated MAGs assigned to each genus depicted in the left subpanel.

**Extended Data Fig. 3.**
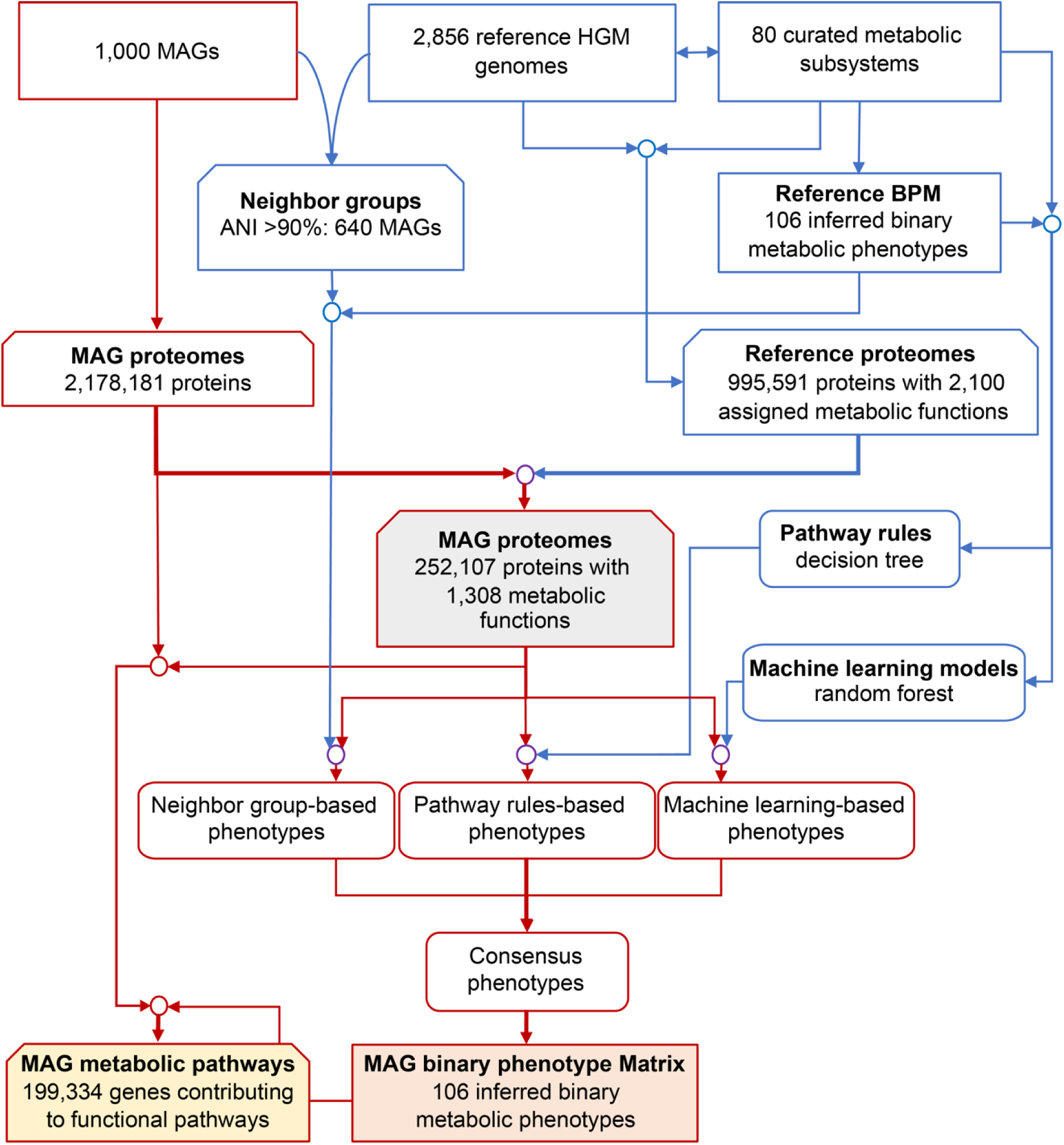
Bioinformatics pipeline for subsystems-based annotation and prediction of functional capabilities (metabolic phenotypes) of MAGs. The flow diagram shows the input data (2,856 reference genomes, 80 curated metabolic subsystems and 1,000 target MAGs) and the main computational steps performed. The pipeline produces two major outputs: (i) a complete set of functionally annotated proteins contributing to 80 reconstructed metabolic subsystems identified in the collection of 1,000 MAGs (annotation results are detailed in **Supplementary Table 5** and **Supplementary Table 6**); and (ii) a Binary Phenotype Matrix (BPM) reflecting the inferred presence or absence of 106 functional metabolic pathways in each of the 1,000 MAGs shown in **Supplementary Table 7a**.

**Extended Data Fig. 4.**
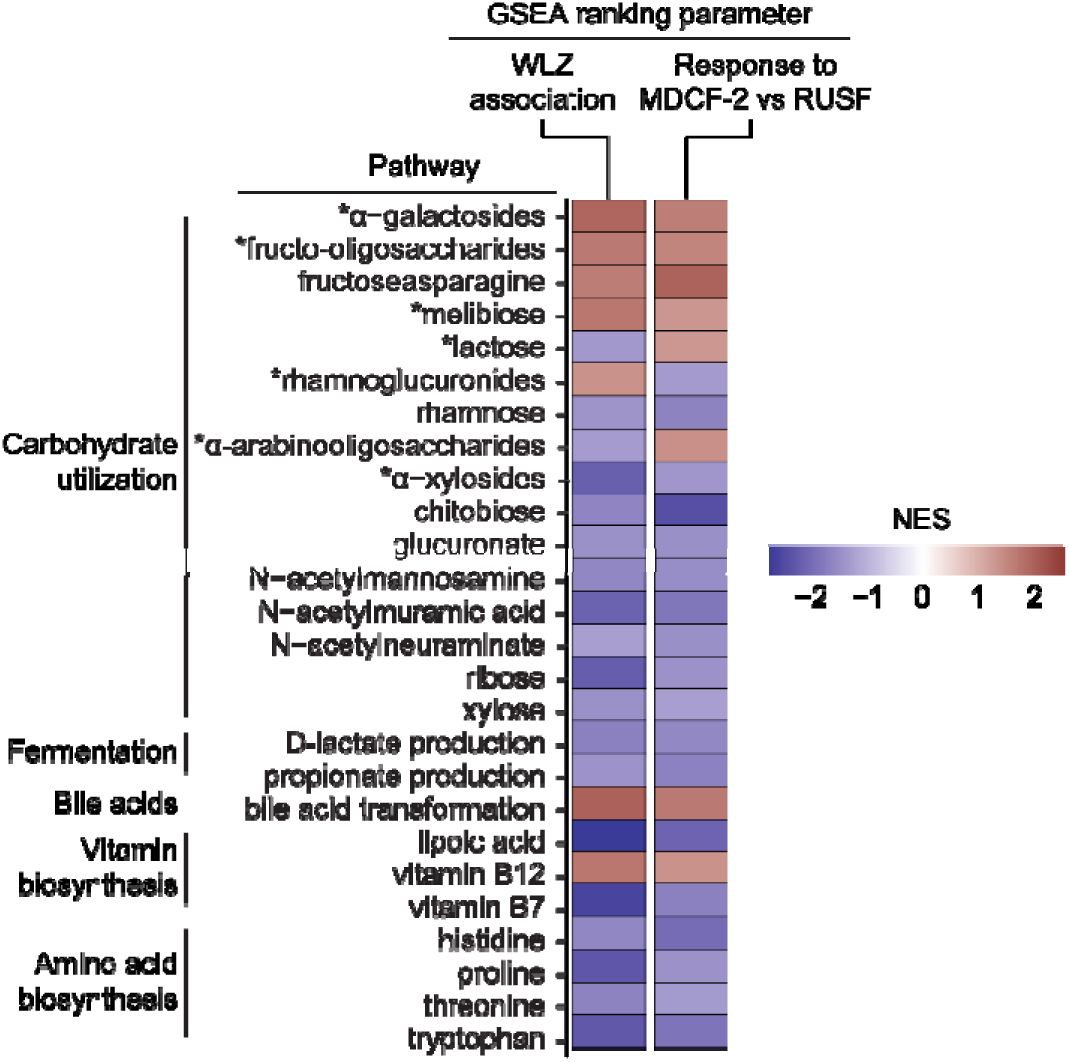
Enrichment of metabolic pathways in WLZ- and treatment-associated MAGs. MAGs were ranked by their WLZ association (negative to positive) or treatment association (RUSF-associated to MDCF-2-associated) and GSEA was employed to determine overrepresentation of pathways in MAGs at the extremes of each ranked list. The results (Normalized Enrichment Score, NES) only include pathways that display a statistically significant enrichment (q<0.05, GSEA) in both the WLZ-associated MAG and treatment-associated MAG analyses. For carbohydrate utilization pathways, disaccharides and oligosaccharides are indicated with an asterisk.

**Extended Data Fig. 5.**
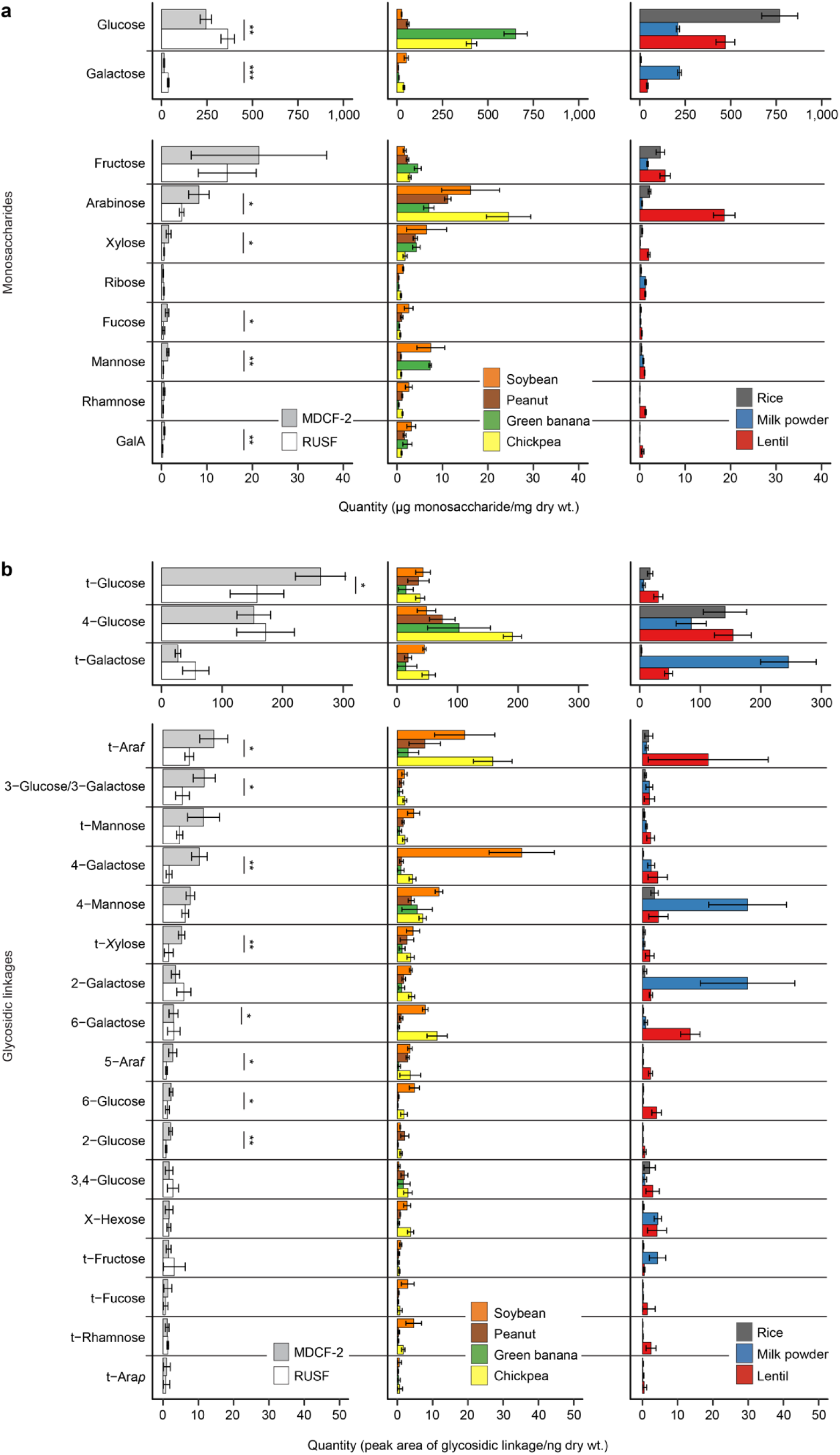
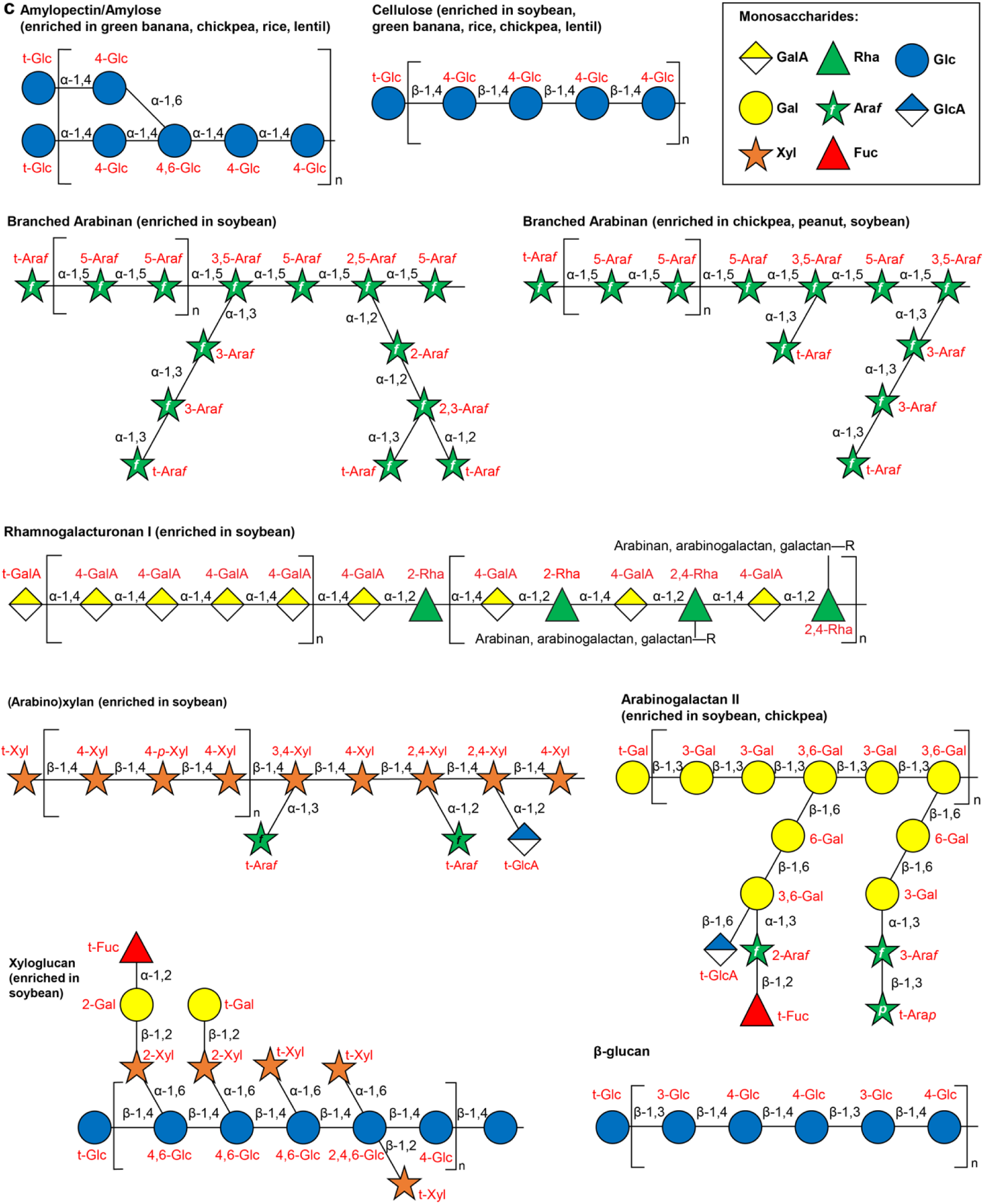
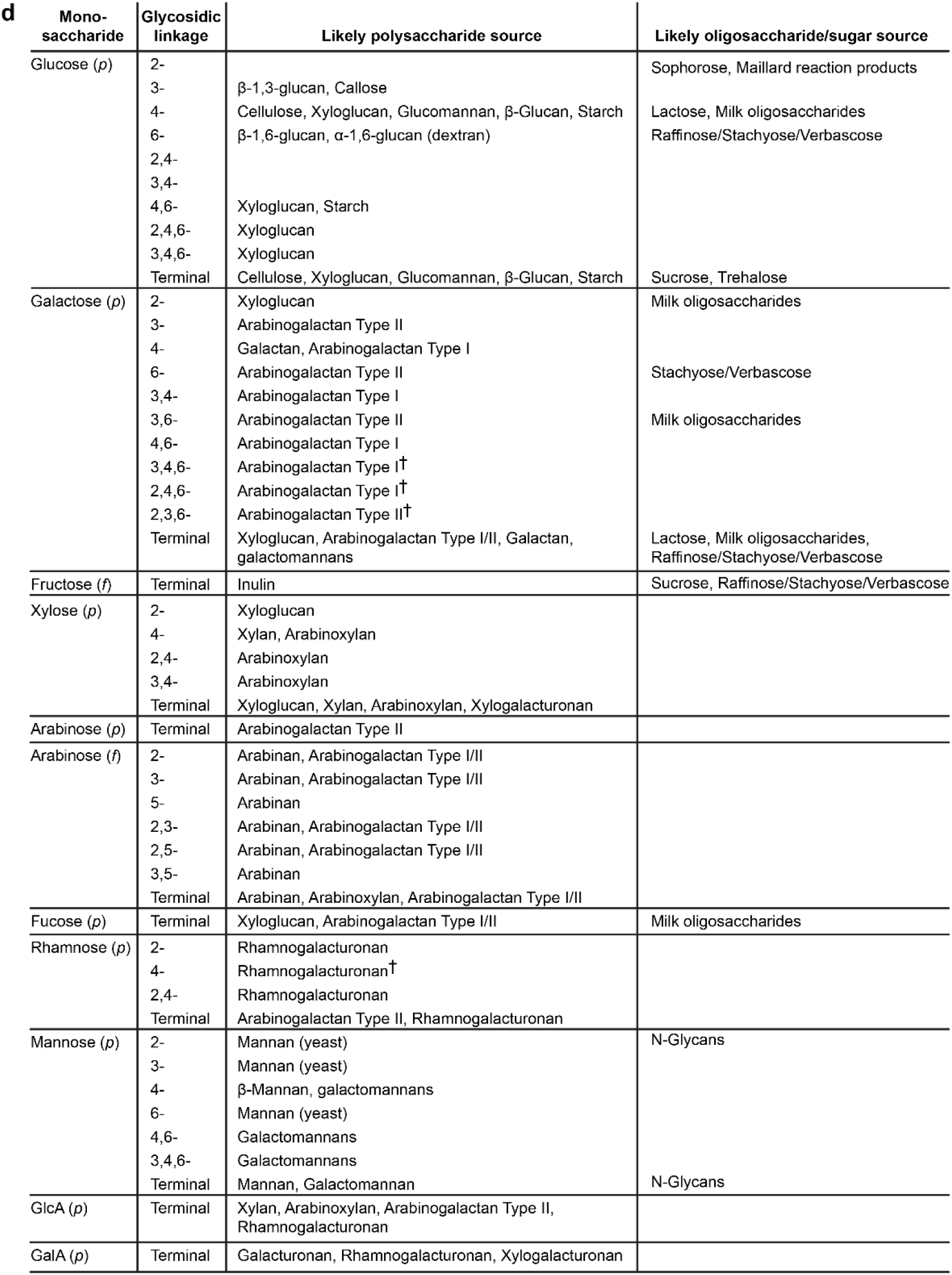
LC-MS analysis of glycans present in MDCF-2, RUSF and their component ingredients. (**a,b**) Analysis of monosaccharides (panel a) and glycosidic linkages (panel b) liberated by hydrolysis of glycans present in MDCF-2 and RUSF, and in the food ingredients used to formulate them. Mean±SD are plotted. *, *P*<0.05, **, *P*<0.01 (t-test). (**c**) Structures of glycans enriched in components of MDCF-2 or RUSF. (**d**) Measured glycosidic linkages in MDCF-2 and RUSF and their likely polysaccharide sources. Sources were inferred based on analysis of literature as well as polysaccharide standards. ‘†’ refers to instances where literature reports were not available and commercial polysaccharide preparations were used for direct experimental validation.

**Extended Data Fig. 6.**
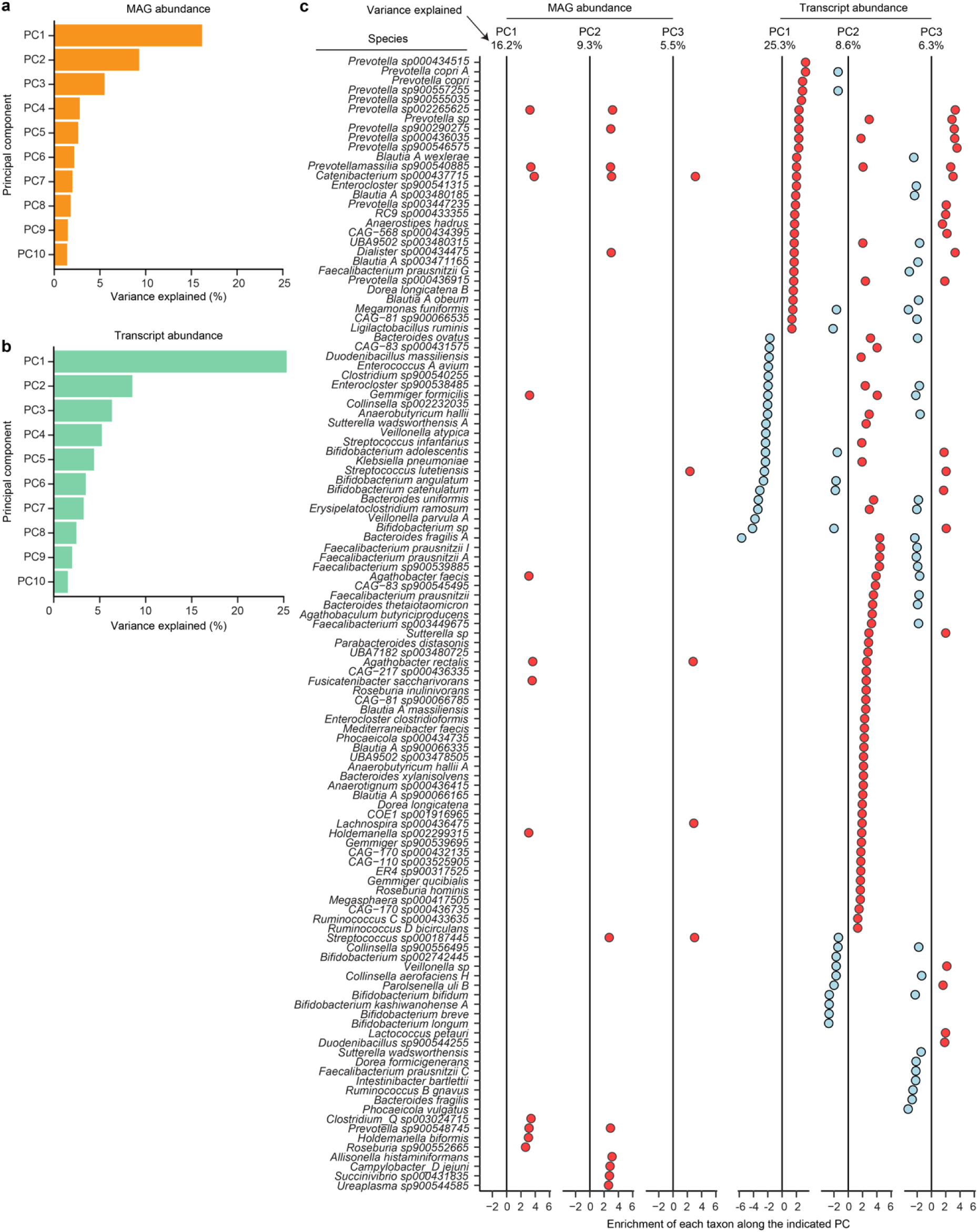
Principal components analysis of transcript and MAG abundances in fecal specimens. (**a,b**) Percent variance explained by the top 10 principal components of a PCA analysis including abundance of MAGs (panel a) at the 0, 2, 4, 8, 12, and 16 week time points in Fig. 1a, or transcripts (panel b) at 0, 4, and 12 weeks. (**c**) Significantly enriched taxa (q<0.05, GSEA) along the first three principal components (PC1-PC3) of the fecal microbiome or meta-transcriptome PCA.

**Extended Data Fig. 7.**
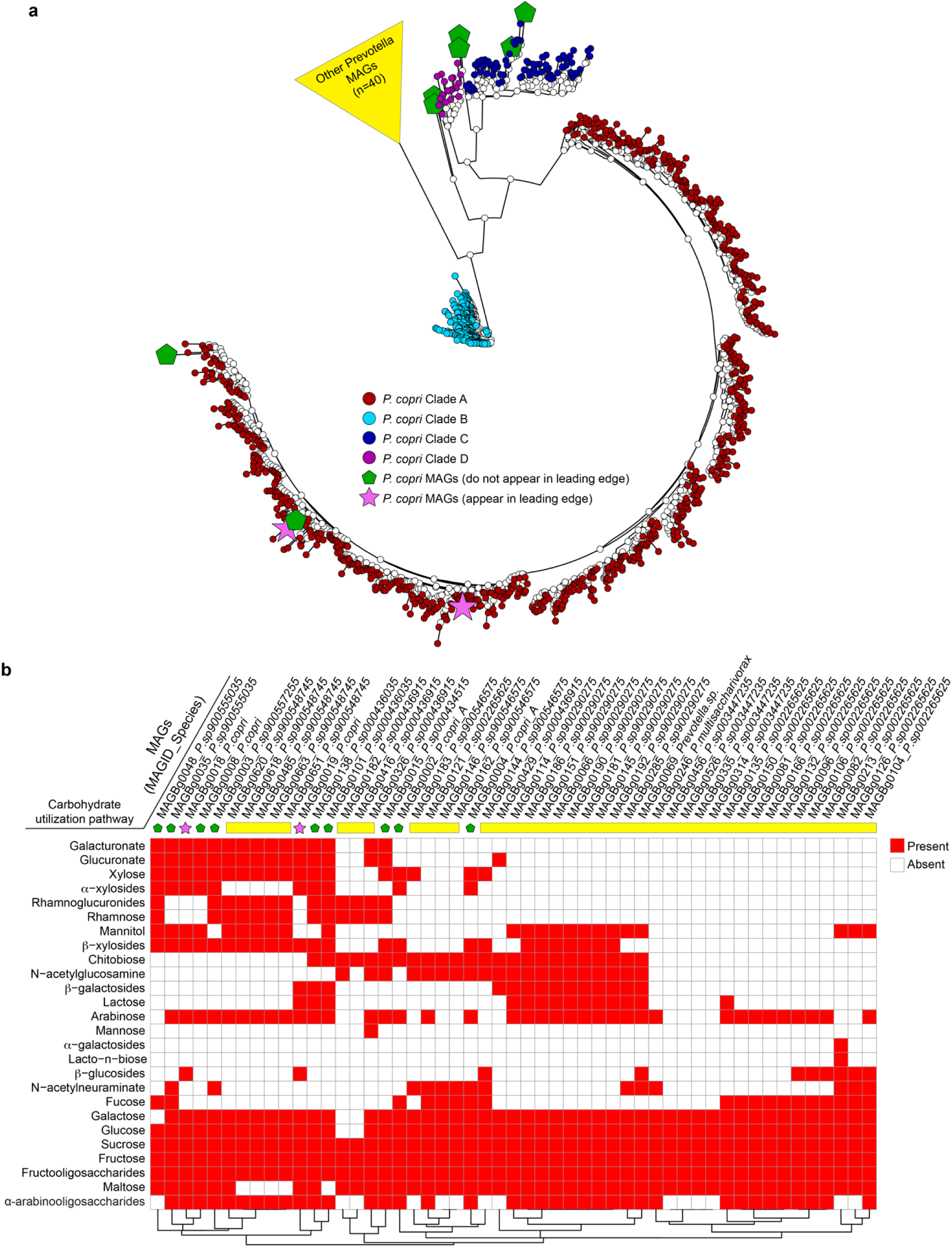
Phylogenetic tree and representation of mcSEED carbohydrate utilization pathways in *Prevotella* MAGs. (**a**) Unrooted, marker gene-based phylogenetic tree of 51 *Prevotella* MAGs from this study, plus 1,049 *P. copri* genomes and MAGs previously assigned to each of four clades^20^. Pink stars denote the two WLZ-associated *P. copri* MAGs. The nine remaining *P. copri* MAGs from this study are highlighted by the green pentagons. The 40 *Prevotella* MAGs not classified as *P. copri*, based on an average branch length >0.5 from all 1,049 reference *P. copri* isolates, are grouped together and depicted as a yellow triangle. (**b**) mcSEED carbohydrate utilization pathways in 51 *Prevotella* MAGs from the current study. MAGs are hierarchically clustered based on the predicted presence (red) or absence (white) of these pathways.

**Extended Data Fig. 8.**
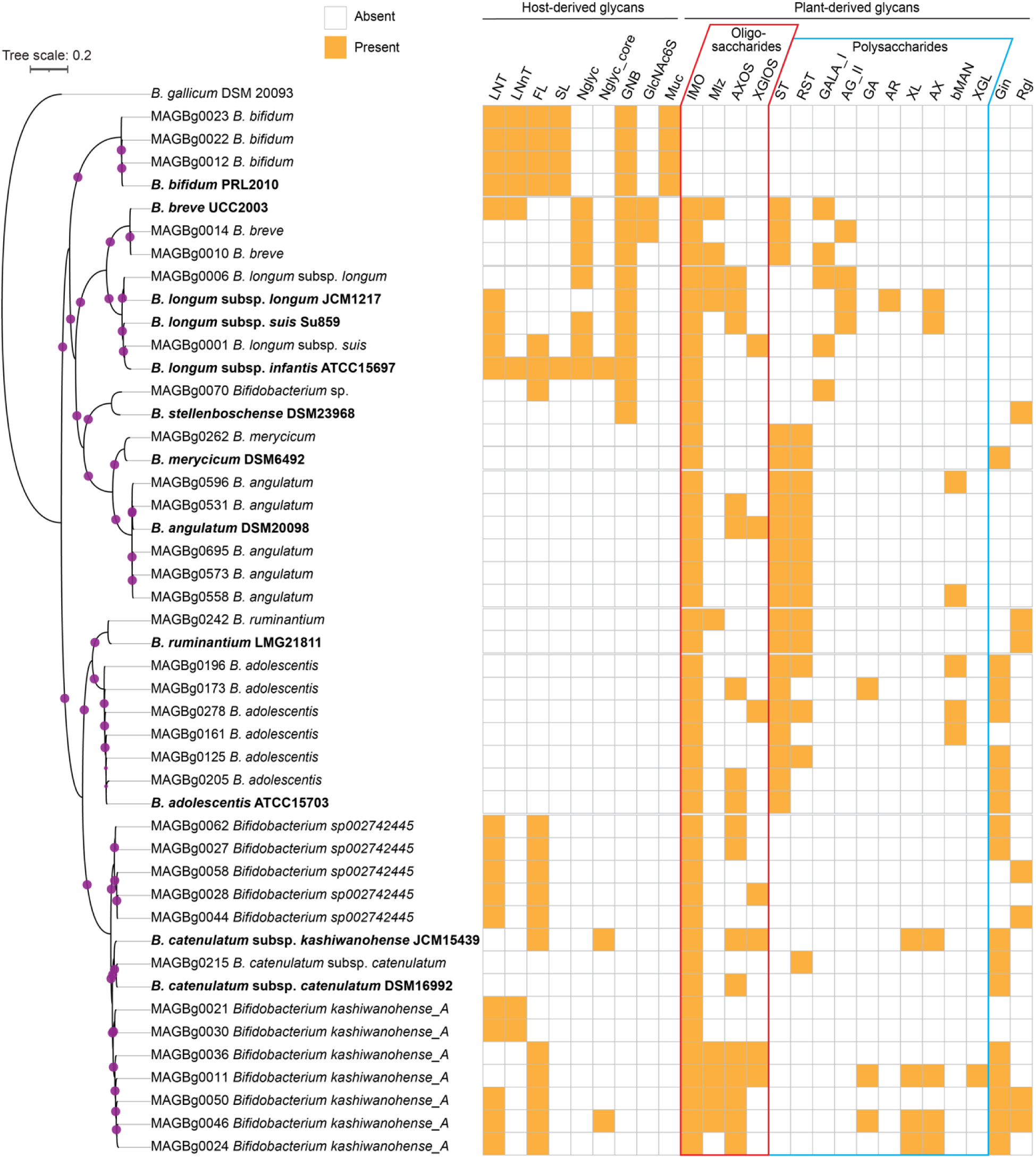
Phylogenetic tree and inferred carbohydrate utilization phenotypes of *Bifidobacterium* MAGs. The phylogenetic tree indicates the relatedness of 34 *Bifidobacterium* MAGs and 14 reference genomes, as determined by sequence similarity among 142 core genes. The size of the pink circles in the dendrogram correspond to bootstrap support for the nodes (out of 100 bootstraps). Type stains used for taxonomic assignments and phenotypic comparisons are bolded. The matrix describes the presence (orange) or absence (white) of 25 predicted carbohydrate utilization phenotypes encompassing host- and plant-derived glycans. LNT, lacto-*N*-tetraose; LNnT, lacto-*N*-neotetraose; FL, 2- and 3-fucosyllactose; SL, 3- and 6-sialyllactose; Nglyc, *N*-glycans; Nglyc_core, *N*-glycan core (Fucα1-6GlcNAcβ1-Asn); GNB, galacto-*N*-biose; GlcNAc6S, *N*-acetylglucosamine-6-sulfate; Muc, mucin *O*-glycans; IMO, isomaltooligosaccharides and panose; Mlz, melezitose; AXOS, arabinoxylooligosaccharides; XGlOS, xyloglucan oligosaccharides; ST, starch and glycogen; RST, resistant starch; GALA_I, type I galactan and arabinogalactan; AGII, type II galactan and arabinogalactan; GA, gum arabic; AR, arabinan; XL, xylan; AX, arabinoxylan; bMAN, β-mannan; XGL, xyloglucan; Gin, ginsenosides; Rgl, rhamnoglycosides.

**Extended Data Fig. 9.**
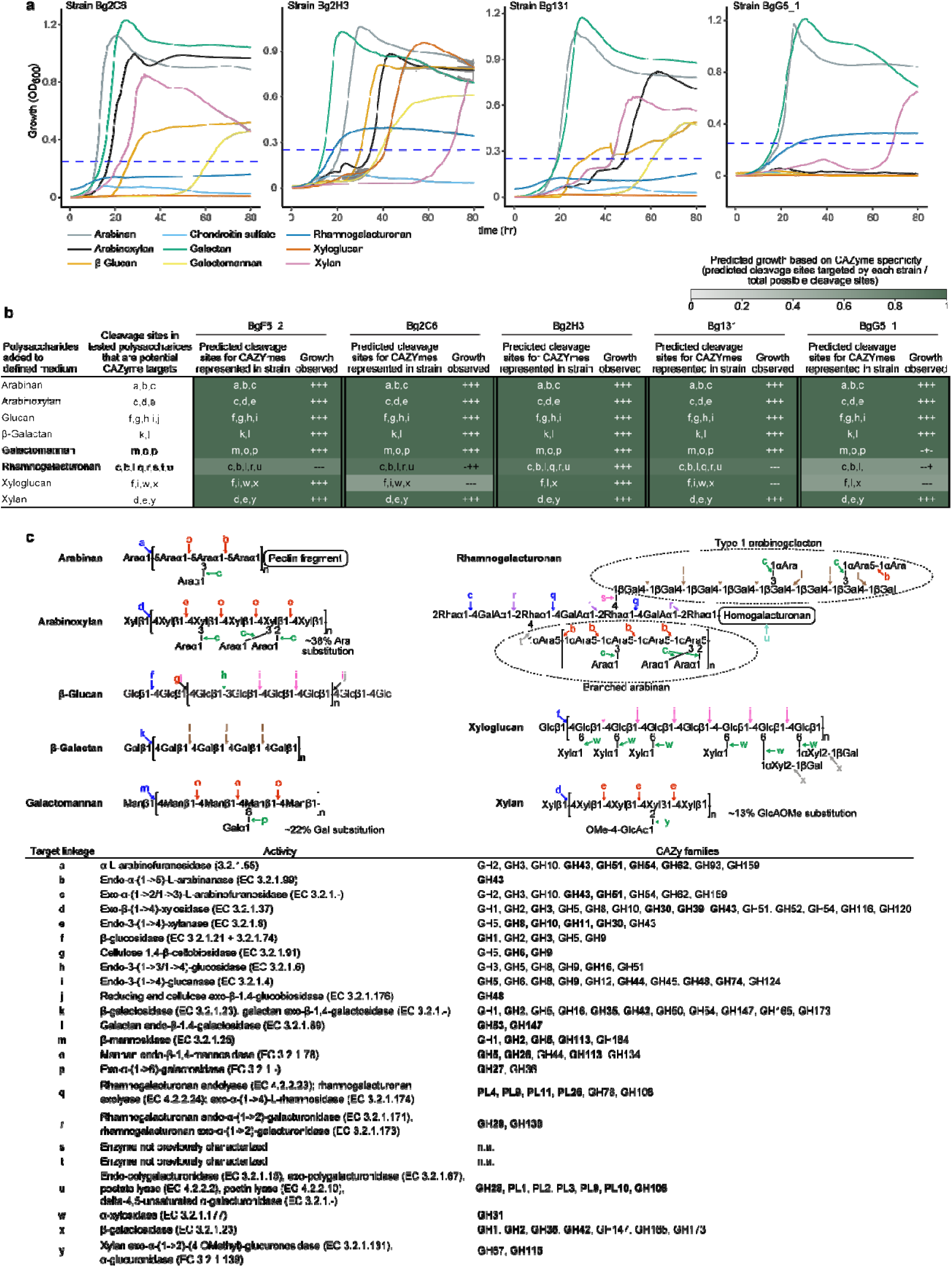
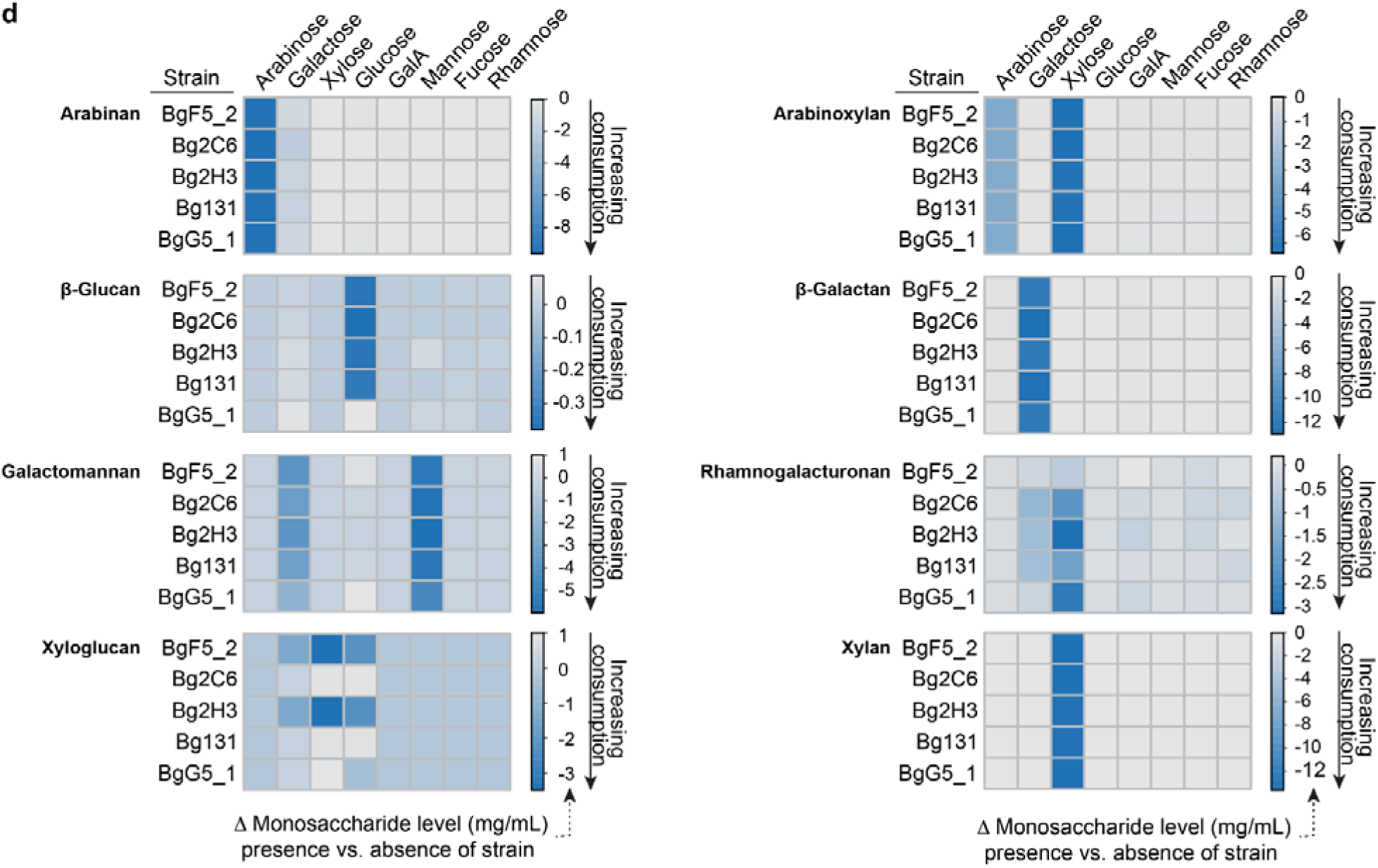
Growth of Bangladeshi *P. copri* strains in defined medium supplemented with purified polysaccharides. (**a**) Growth curves in defined medium containing individual purified polysaccharides similar to those that are abundant in or unique to MDCF-2 compared to RUSF. Curves describe mean values ± sd for OD_600_ measurements (n=3 replicates/growth condition). (**b**) Growth of *P. copri* strains versus a PUL-based prediction of their growth phenotypes. Growth is expressed as ‘+’ or ‘-’ for each of the triplicate cultures according to whether a threshold OD_600_ > 0.25 was attained. The color key expresses ‘prediction’ as the fraction of possible cleavage sites in each polysaccharide that are known or predicted to be targeted by the PUL-associated CAZymes of a given strain. (**c**) CAZymes present in the PULs of each strain and their predicted activities against glycosidic linkages in each purified polysaccharide. Linkages in a polysaccharide that are predicted to be targeted by PUL-associated CAZymes are labeled ‘a,b…y’ in both panels b and c. All CAZyme family assignments for a given enzymatic activity are shown; the family that displays this activity most commonly is noted in bold font. (**d**) Heatmaps representing bacterial consumption of monosaccharides present in the different polysaccharides. UHPLC-QqQ-MS-based monosaccharide analysis was performed using defined medium harvested from monocultures of the *P. copri* stains. Control incubations did not contain added bacteria. Cells in each matrix show the mean difference (at the end of the 189 hour-long incubation) between the concentration of each monosaccharide in three technical replicates of each strain/polysaccharide combination compared to the corresponding uninoculated control.

**Extended Data Fig. 10.**
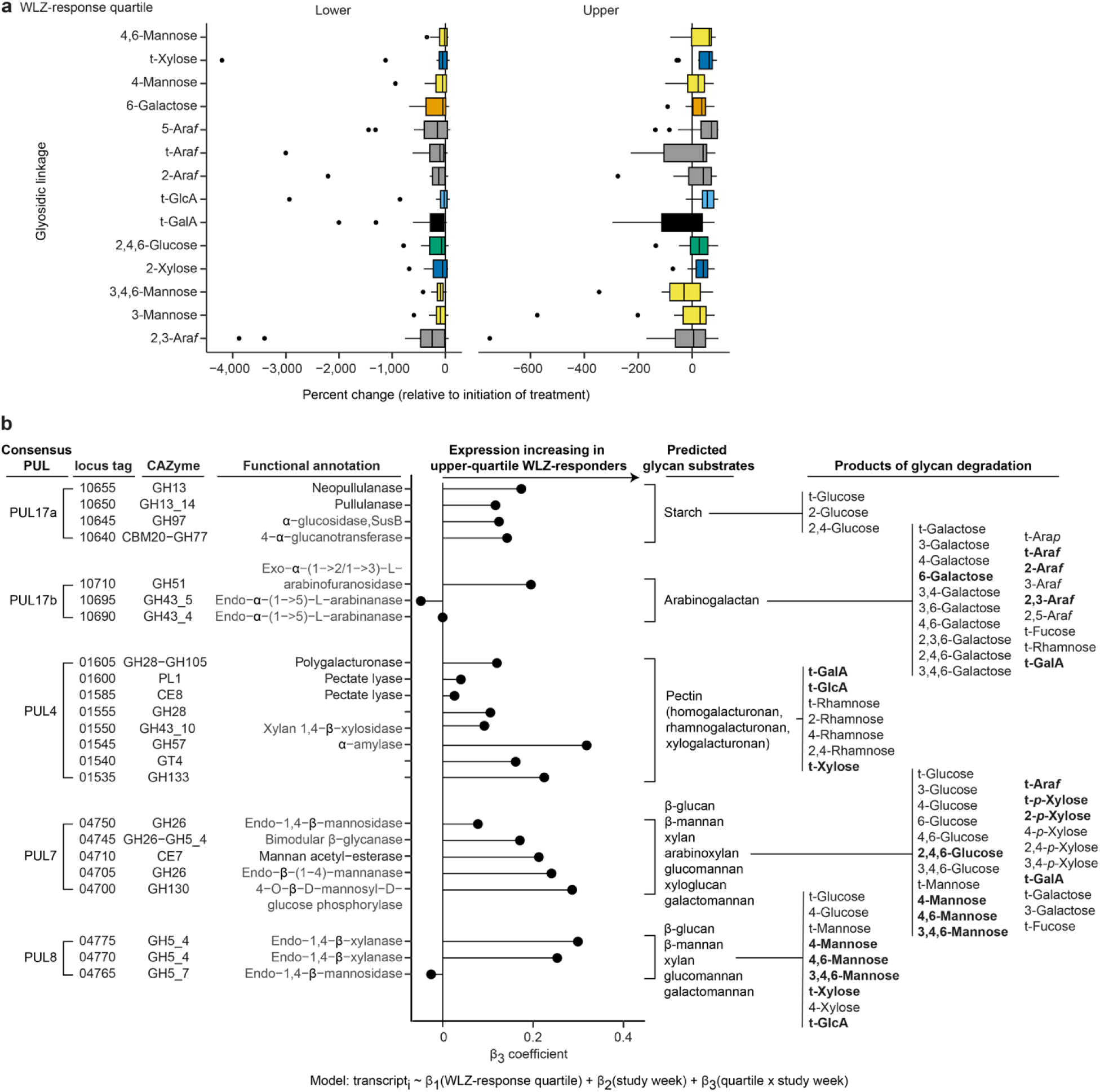
Changes in levels of fecal glycosidic linkages and expression of *P. copri* CAZyme genes after MDCF-2 treatment. (**a**) Boxplot of changes in the levels of fecal glycosidic linkages relative to initiation of treatment among upper and lower WLZ quartile responders. Levels of these 14 linkages increased to a significantly greater extent over time in the upper vs lower WLZ response quartiles (Model: linkage abundance ∼ WLZ-response quartile × study week + (1|PID)). Boxplots indicate the median, first and third quartiles; whiskers extend to the largest value no further than 1.5 × the interquartile range. (**b**) The β_3_ coefficient for the interaction of WLZ-response quartile and study week is shown for CAZymes in consensus PULs in Bg0018 and Bg0019. Predicted PUL substrates and potential glycosidic linkages in each of these substrates are shown on the right. Glycosidic linkages whose abundances were significantly different in fecal samples from the upper versus lower WLZ quartile responders are highlighted in bold font (see Fig. 5a).

**Extended Data Fig. 11.**
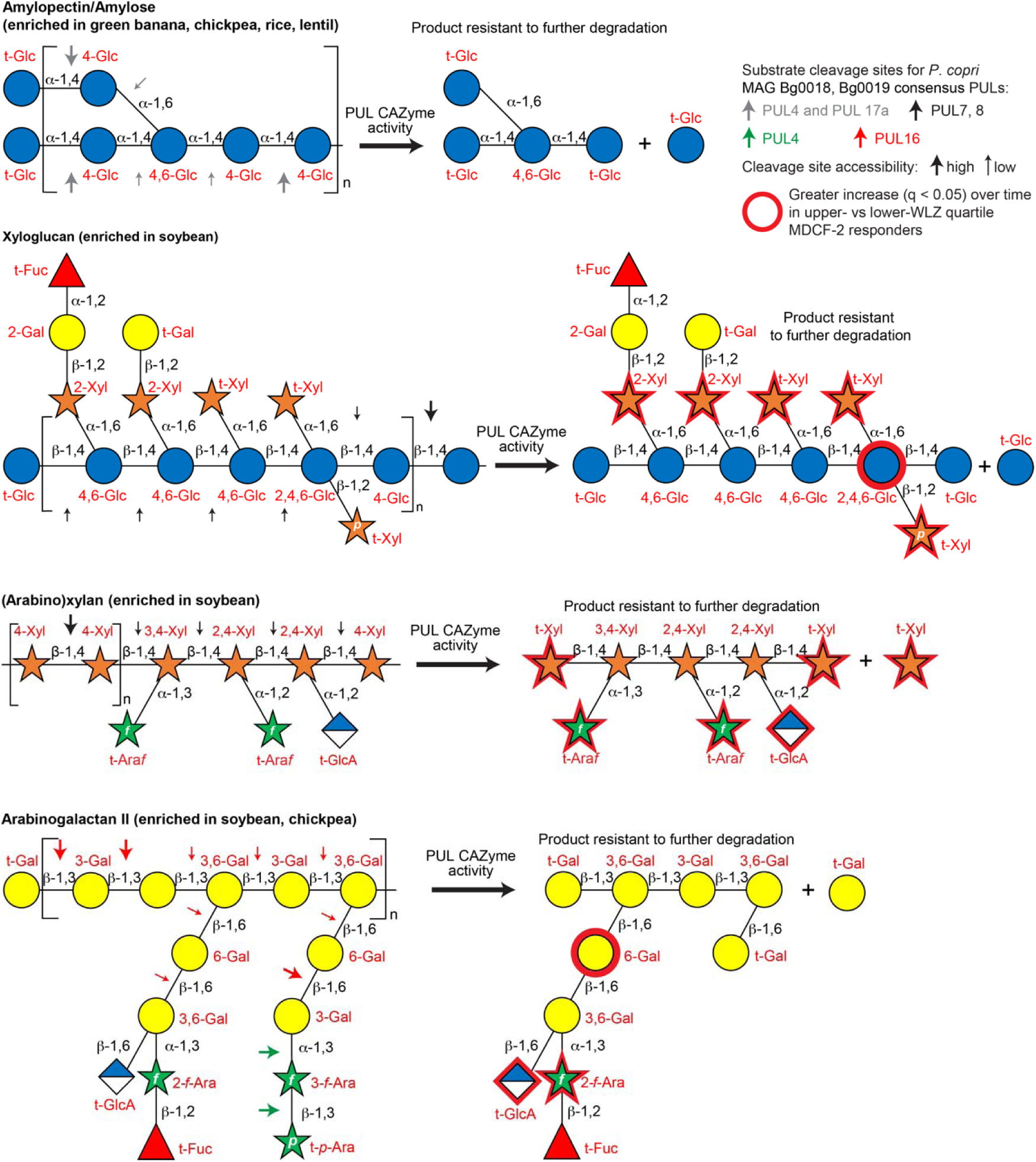
Polysaccharide structures, cleavage sites, and predicted products of CAZyme activity. Glycosidic linkages highlighted with arrows are those predicted as sites of cleavage by CAZymes expressed by the set of PULs, described in Fig. 4a, that are present in *P. copri* MAG Bg0019 and/or Bg0018. Consensus PUL numbers are listed except in the case of Bg0019 PUL3, which is not represented in Bg0018 (see **Supplementary Table 15**). The size of the arrows (large versus small) denotes the relative likelihood (high versus low, respectively) of cleavage of glycosidic linkages by *P. copri* CAZymes when considering steric hindrance at branch points.

**Extended Data Fig. 12.**
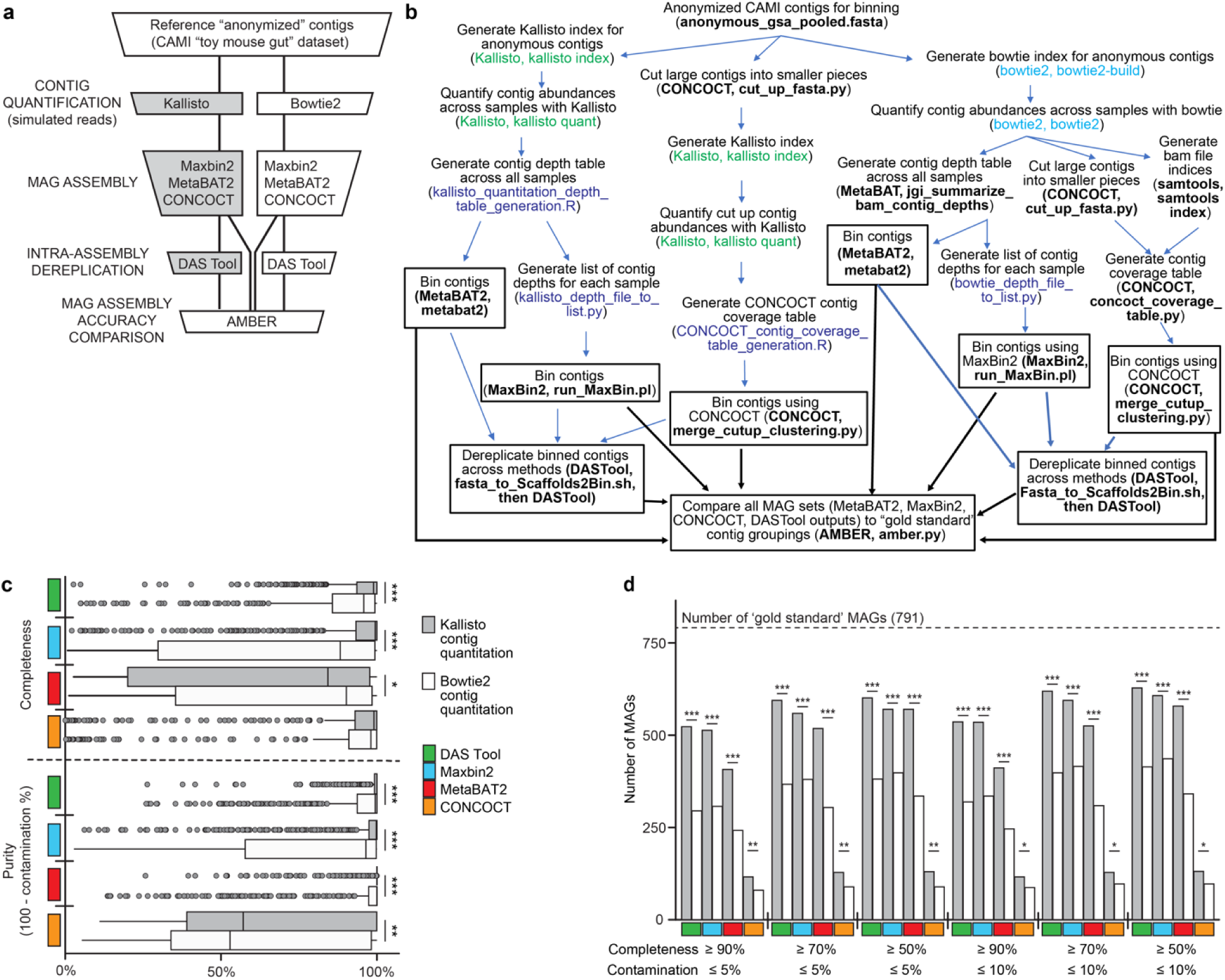
Validation of the MAG assembly pipeline. (**a**) Bioinformatic workflow for comparing the fidelity of MAG assembly from alignment-based (bowtie2) versus pseudoalignment-based (kallisto) contig quantitation. (**b**) A detailed description of the workflow is described in panel a. Each box includes a summary of the computational task plus the name of the program and, where relevant, included the command used to complete the task (in parentheses). Color of the text in parentheses: brown, default code from kallisto; purple, default code from bowtie2; blue, custom script written to achieve tasks described; black, default code used for programs to assemble MAGs and dereplicate contigs. Boxes with a black outline and the thick black arrows emanating from them indicate that binned contigs were used as input to AMBER for MAG assembly comparisons across methods. (**c**) Boxplot describing summary statistics (completeness and purity) for MAG assembly approaches. Boxplots indicate the median, first and third quartiles; whiskers extend to the largest value no further than 1.5 **×** the interquartile range. (**d**) Number of MAGs, obtained from each assembly and quantitation strategy, that are distributed across two quality metrics (completeness, contamination). The number of ‘gold standard’ MAGs (theoretical maximum) in the simulated metagenomic dataset is indicated by a horizontal dashed line. *, *P*<0.05; **, *P*<0.01; ***, *P* < 0.001 (Wilcoxon test for panel c; Fisher’s exact test for panel d).

## SUPPLEMENTARY INFORMATION

### Supplementary Methods

#### Metagenome assembled genomes (MAGs)

##### Short-read shotgun sequencing

DNA was isolated from 942 fecal samples as previously described^4^ and shotgun sequencing libraries were prepared using a reduced-volume Nextera XT (Illumina) protocol^46^. Libraries were quantified, balanced, pooled, and sequenced [Illumina NovaSeq 6000, S4 flow cell; 2.3±1.4×10^7^ 150 nt paired-end reads/sample (mean±SD)]. Reads were demultiplexed (bcl2fastq, Illumina), trimmed to remove low-quality bases, and processed to remove read-through adapter sequences (Trim Galore^34^, v0.6.4). Read pairs where the length of either read was <50 nt after quality and adapter trimming were discarded. The remaining reads were mapped to the human genome (UCSC hg19) using bowtie2^47^ (v2.3.4.1) and were subsequently filtered to remove *H. sapiens* sequences.

Preprocessed, short-read shotgun data were aggregated from each participant’s fecal sample set (n=7-8 samples/participant; 118 participants) prior to MAG assembly. This strategy was adopted to enable the contig abundance calculations required by the MAG assembly algorithms employed below, while at the same time mitigating the risk of chimeric assemblies inherent to co-assembly across individuals. Assemblies were generated for all 118 datasets using MegaHit^48^ (v1.1.4), and the resulting contigs were quantified in each assembly by mapping preprocessed reads to the assembled contigs with kallisto^36^. Contigs were assembled into MAGs using MaxBin2^49^ (v2.2.5) and MetaBAT2^50^ (v2.12.1). The parallel results of both binning strategies were merged and dereplicated using DAS Tool^51^ (v1.1.2) on a per-participant basis.

##### Long-read shotgun sequencing

We applied long-read sequencing to fecal samples obtained at the 0- and 3-month time points from each of the 15 upper quartile WLZ responders in the MDCF-2 treatment group. Aliquots containing 400-1000 ng of DNA from each biospecimen were transferred to a 96-well, 0.8 mL, deep-well plate (Nunc, ThermoScientific) and prepared for long-read sequencing using the SMRTbell Express Template Prep Kit 2.0 (PacBio). All subsequent DNA handling and transfer steps were performed with wide-bore, genomic DNA pipette tips (ART, ThermoScientific). Barcoded adapters were ligated to A-tailed DNA fragments by overnight incubation at 20 °C. Adapter-ligated fragments were then treated with the SMRTbell Enzyme Cleanup Kit to remove damaged or partial SMRTbell templates. A high molecular weight DNA fraction was purified using AMPure beads (ratio of 0.45x well-mixed AMPure bead volume to sample) and eluted in 12 µL of PacBio elution buffer. DNA libraries were sequenced on a Sequel System (Pacific Biosciences) using a Sequel Binding Kit 3.0 and Sequencing Primer v4 with 24 hours of data collection. A total of 3.0×10^9^±9.8×10^8^ bp/sample were collected, with an average subread length of 5,654±871 bp (mean±SD).

Hybrid assembly of short- and long-read data was performed using OPERA-MS^52^ (v0.9.0). OPERA-MS uses assembly graph and coverage-based methods to cluster contigs into MAGs based on optimizing per-cluster Bayesian information criterion (BIC). Prior to hybrid assembly, continuous long reads (CLR) were combined across the two available time points for each participant and reads that mapped to the human genome were removed. Illumina short reads and PacBio long reads (CLR) were provided to OPERA-MS and assembled using the built-in OPERA-MS genome database and default settings (the latter includes polishing of output MAGs with Pilon^53^).

##### MAG dereplication, curation and abundance calculations

After assembling MAGs by both short-read only and short-plus long-read strategies, all MAGs from all assembly strategies were assessed for completeness and contamination (‘lineage_wf’ command in CheckM^54^, v1.1.3) and refined (‘tetra’, ‘outliers’, and ‘modify’ commands in CheckM) to remove contaminating contigs. Additional refinement based on the distribution of phylogenetic markers present in each MAG was performed [‘phylo-markers’, ‘clade-markers’, and ‘clean-bin’ commands in MAGpurify^55^ (v2.1.2)]. A final MAG quality assessment was performed using CheckM, followed by a stringent (≥ 90% complete, ≤ 5% contaminated, ANI ≥ 99%) bulk dereplication across all MAGs from all participants (options ‘-l 50000’, ‘--completeness 90’, ‘--contamination 5’, ‘-pa 0.9’, ‘-sa 0.99’ in dRep^56^ (v2.6.2). The final dataset contained 681±99 (mean±SD) MAGs/participant. MAG assembly summary statistics were collected from CheckM^54^ and quast^57^ analyses (v4.5) and aggregated (**Supplementary Table 1, Extended Data Fig. 12**). Initial MAG annotations were performed using prokka^58^ (v1.14.6). Further details about benchmarking the methods we used for assembly are presented in *Supplementary Discussion*. To quantify the abundance of each MAG in each sample MAGs were processed to create a single kallisto quantification index^59^. Reads from each fecal DNA sample were then mapped to this index.

##### MAG taxonomy

Taxonomic assignments were initially made by employing the Genome Taxonomy Database Toolkit^14^ (GTDB-Tk) and corresponding database (release 95). We complemented these MAG assignments by using Kraken2^60^ (v2.0.8) and Bracken^61^ (v2.5) and a Kraken2-compatible version of the GTDB reference.

*P. copri* has been partitioned into four distinct clades (‘A-D’) based on marker gene phylogeny^20^. To classify *Prevotella* MAGs in this study, we constructed an unrooted, marker gene-based phylogeny using Phylophlan^62^ (v3.0.60). This tree encompassed 17 reference isolate genomes and 1006 MAGs from a previous study^20^ plus any MAGs from our set classified by GTDB-Tk as belonging to the genera *Prevotella* (n=51) or *Prevotellamassilia* (n=13). Putative *Prevotella* MAGs from the present study that clustered within the four previously identified *P. copri* clades were assigned to the corresponding clade based on visualization with Graphlan^63^ (v1.1.4).

Certain *Bifidobacterium* species consist of multiple closely related subspecies (e.g., *B. longum*). Therefore, we calculated a pan-genome for 34 *Bifidobacterium* MAGs in our dataset (**Supplementary Table 1**), plus 14 reference isolate genomes (**Extended Data Fig. 8**), using Roary^64^ (v3.12.0) and a 60% minimum sequence identity threshold for blastp^65^. The reference isolate genomes included 10 *Bifidobacterium* species and three subspecies of *B. longum* (subsp. *longum*, *infantis*, and *suis*). Concatenated nucleotide sequences of 142 identified core genes were aligned using MAFFT^66^ (v7.313). The resulting alignment was trimmed [microseq^67^ R package (v2.1.4)] and then used to construct a maximum likelihood phylogenetic tree [IQ-TREE^68^ (v1.6.12)]. The *B. gallicum* DSM 20093 genome was selected as an outgroup. Putative *Bifidobacterium* MAGs from this study that clustered together with reference genome clades were assigned to the corresponding clade. Using this method, we were able to confirm or update our initial GTDB-Tk-based classifications of all *Bifidobacterium* MAGs and resolve nearly all closely related subspecies (**Supplementary Table 1**, **Extended Data Fig. 8**).

#### Culturing *P. copri* from fecal samples and genome sequencing

Fecal samples, obtained from our previously reported studies of Bangladeshi children living in Mirpur^3,4^, were first screened based on the abundances of *P. copri* V4-16S rDNA ASVs (amplicon sequence variants) and/or *P. copri* MAGs. Five samples from our previous pilot MDCF study^3^, plus an additional 32 samples from the larger randomized controlled clinical trial (prioritized based on participants’ membership in the upper quartile of WLZ response to MDCF-2 treatment) were selected for this culturing effort.

A frozen aliquot (∼0.1 g) of each selected fecal sample was weighed. All subsequent steps were performed under anaerobic conditions (atmosphere; 75% N_2_, 20% CO_2_, 5% H_2_) in a Coy chamber (Coy Laboratory Products, Grass Lake, MI). For the fecal samples that yielded strains BgD5_2, BgF5_2, and BgG5_1, aliquots were resuspended directly in 100 µL of phosphate-buffered saline (PBS) containing 0.5% w/v L-cysteine. All other samples were clarified as described previously^3^. A 100 µL aliquot of each resuspended or clarified sample was serially diluted in PBS containing 0.5% w/v L-cysteine and plated onto Laked sheep blood/Kanamycin/Vancomycin (LKV) agar plates (Hardy Diagnostics, Cat # A60). *Prevotella spp.* produce a pigment known to fluoresce brick red when exposed to ultraviolet light. Therefore, colonies grown from serial dilutions were screened for this phenotype, picked and struck onto Brain Heart Infusion (Difco, Cat # 241830) agar plates containing 10% horse blood. Individual isolated colonies were picked into liquid Wilkins-Chalgren medium (Oxoid, Cat # CM0643), grown overnight at 37 °C and then mixed 1:1 with pre-reduced 30% glycerol (in PBS plus 0.5 % w/v L-cysteine) in 1.8 mL glass vials (E-Z vials, Wheaton). Vials were closed by crimping on a cover containing a PTFE/grey butyl liner (Wheaton). Stocks were preserved at -80 °C for later use. An additional aliquot of each culture was subjected to full-length 16S rDNA sequencing (universal primers 8F and 1391R) to provide an initial taxonomic identification. This effort yielded a total of 108 isolates assigned to *Prevotella spp*, including 86 classified as *P. copri*.

Each *P. copri* isolate was retrieved from storage, cultured in Wilkins-Chalgren medium, and subjected to whole-genome sequencing. Freezer stocks of each bacterial isolate were inoculated into Wilkins-Chalgren medium and grown at 37 °C under anaerobic conditions without shaking until reaching late-log phase. A 6 mL aliquot of each culture was withdrawn from the anaerobic chamber and centrifuged at 5,000 x g for 5 minutes. The resulting cell pellet (10-50 mg) was transferred to a 2 mL cryotube and DNA was extracted by bead beating (Biospec Minibeadbeater-96) for 1 minute with a 3.97 mm steel ball and 250 μL of 0.1 mm zirconia/silica beads in 500 μL of 25:24:1 phenol:chloroform:isoamyl alcohol solution, 210 μL of 20% SDS, and 500 μL of 2X buffer A (200 mM NaCl, 200 mM Trizma base, 20 mM EDTA)^69^. The resulting mixtures were centrifuged at 3,220 x g for 4 minutes at 22 °C. A 420 μL aliquot of the resulting aqueous phase was transferred to a deep 96-well plate and purified using a QIAquick 96-well PCR purification kit (Qiagen). DNA was quantified (Quant-iT dsDNA broad range kit; Invitrogen) and fragment size distribution was measured [TapeStation using a genomic DNA ScreenTape (Agilent)].

Purified DNA was prepared for long-read sequencing using a SMRTbell Express Template Prep Kit 2.0 from Pacific Biosciences (Pacific Biosciences; PacBio); we followed the manufacturer’s instructions for creating HiFi Libraries from low DNA input, with adjustments made to accommodate a 96-well plate configuration^69^. The DNA concentration and fragment size distribution of resulting libraries were evaluated (genomic DNA ScreenTape; TapeStation instrument) and libraries were pooled at equimolar concentrations after normalizing for expected genome size. Pooled libraries were sequenced on a Sequel long-read DNA sequencer (PacBio) using a Sequel Binding Kit 3.0 and Sequencing Primer v4, with 24 hours of data collection. Samples were demultiplexed, and Q20 circular consensus sequencing (CCS) reads were generated using the Cromwell workflow from PacBio. The Flye^70^ assembler (v2.8.1) was used to assemble the genomes, with the HiFi-error set to 0.003, min-overlap set at 2000, and other options kept to default. Genome quality was evaluated using CheckM^54^ (v1.1.3), and genomes were annotated using prokka^58^ (v1.14.6).

#### Subsystems-based annotation and prediction of functional capabilities (‘inferred metabolic phenotypes’) of MAGs and cultured *P. copri* strains

MAG and isolate genes were assigned functions, and metabolic pathways were reconstructed using a combination of (i) public domain tools for sequence alignment and clustering, (ii) custom scripts to process the results of sequence alignments (e.g., for domain annotation in multifunctional proteins), and (iii) a reference collection of 2,856 human gut bacterial genomes for which we have reconstructed and manually-curated metabolic pathways related to 98 distinct metabolites and 106 metabolic phenotypes^17^. These annotations are captured in the mcSEED database, a microbial community-centered adaptation of the SEED genomic platform^15,16^, featuring subsystems-based annotation and pathway reconstruction applied to representative human gut bacterial genomes that were initially automatically annotated by RAST or downloaded from the PATRIC (recently renamed Bacterial and Viral Bioinformatic Resource Center, BV-BRC) database^71^. Each mcSEED subsystem includes a set of functional roles (e.g., enzymes, transporters, transcriptional regulators) that contribute to the prediction of functional metabolic pathways and pathway variants^72^ involved in utilization and catabolism carbohydrates and amino acids, biosynthesis of vitamins/cofactors and amino acids, and generation of fermentation end-products such as short-chain fatty acids. A complete list of MAG genes comprising these pathways, their abbreviations and functions is provided in **Supplementary Table 5** (annotations based on the January 2021 version of the mcSEED database).

Our annotation workflow is illustrated in **Extended Data Fig. 3**. Briefly, we constructed a reference database containing 995,591 functionally annotated proteins comprising the entire set of curated metabolic subsystems from the 2,856 reference genomes, plus an additional 2,988,751 proteins (‘outgroup’ not included in these metabolic subsystems), clustered at 90% amino acid identity (‘cluster’ command, MMSeqs^73^; v1-c7a89). We aligned the predicted protein sequences from the set of 1,000 high-quality MAGs against this reference protein database (DIAMOND^74^, v2.0.0). To account for any influence of MAG fragmentation on metabolic reconstruction, we also identified gene fragments using prodigal^75^ (v2.6.3) and annotated them in parallel. We implemented the following method to account for instances of multidomain structure that require multiple annotations. For each MAG query protein, we used the top 50 hits based on the bitscore, clustered the start and end position coordinates of the corresponding alignments using DBSCAN (Scikit-learn^76^), used the center of each clustered start and end position as potential domain boundary coordinates, and split query proteins into domains with database hits attributed to the corresponding domains. Next, for each domain ≥35 amino acids we used gaussian kernel density modeling (KernelDensity function, neighbors module, Scikit-learn^77^, v0.22.1) of the sequence identity distribution of each set of hits to that domain. A highest local minimum (argrelextrema function, signal module, Scikit-learn) was employed as a threshold to remove low confidence hits. Finally, functional annotations were applied from the reference database to each query protein or domain by majority rule within each set of high-scoring, domain-specific reference hits. High-identity hits to proteins from the outgroup of the reference database were used as criteria to vote “against” applying annotation to each query. This procedure yielded a set of 199,334 annotated MAG proteins, representing 1,308 unique protein products across a set of 80 mcSEED subsystems (**Supplementary Table 5**).

##### Phenotype prediction strategies (Extended Data Fig. 3)

We integrated the results of gene-level functional annotation into *in silico* predictions of the presence or absence (denoted as binary: “1” for presence or “0” for absence) of 106 functional metabolic pathways (**Supplementary Table 7**) using a semi-automated process based on a combination of the following three approaches:

###### Pathway Rules (PR)-based phenotype predictions

This approach uses explicit logic-based “pathway rules” to assign binary phenotypes. These rules combine (i) expert curators’ knowledge regarding the gene composition of various metabolic pathway variants contained in the mcSEED database with (ii) a decision tree method to identify patterns of gene representation in reference genomes corresponding to an intact functional pathway variant (and a respective binary phenotype value denoted as “1”). A total of 106 functional pathway-specific decision trees were generated (Rpart^78^, v4.1.15), where the presence or absence of a particular phenotype was the response variable, and the presence or absence of functional roles (encoded by genes) in each reference pathway were predictor variables. The resulting pathway rules were formally encoded into a custom R script that allowed us to process MAG gene data and assign values (1 or 0) for each of the 106 functional metabolic pathways.

###### Machine Learning (ML)-based phenotype predictions

We compared >30 ML methods (Caret^79^, v6.0.86), using a ‘leave one out’ cross-validation approach in which we removed a single reference genome from the set of 2,856 reference genomes, trained ML models on the remaining genomes, then applied the models to the “test” genome to predict phenotypes. This procedure was then repeated for each genome and each metabolic phenotype. The results of this analysis identified Random Forest as the best-performing method (i.e., it produced the greatest number of correctly predicted phenotypes in our reference training dataset). We then built Random Forest models for each phenotype based on the reference dataset, optimized model parameters using a grid search, and used these models to predict binary (1/0) values for the same set of 106 phenotypes for all MAGs.

###### Neighbor Group (NG)-based phenotype predictions

This approach identifies reference bacteria that are closely related to the MAGs in this study and uses these high-quality reference genomes for phenotype predictions that are robust to variation in MAG quality. Examination of groups of closely related reference organisms suggested that close phylogenetic neighbor genomes tend to either possess or lack an entire pathway variant, whereas more distant neighbors (e.g., other neighbor groups) often carry more divergent pathway variants that specify the same phenotype. We used this observation to develop heuristics that minimize false negative phenotype assignments emerging from the other two prediction strategies. We compiled a set of NGs comprised of MAGs and closely related reference genomes (Mash/MinHash^80^ distance ≤ 0.1, corresponding to ANI ≥90%). At this similarity threshold, we assigned 640 of the 1,000 MAGs from this study to NGs containing as few as four to more than 100 members. Within each NG and for each metabolic pathway, we tentatively assigned a binary phenotype value for a given MAG based on the NG genome with the closest matching gene annotation pattern (based on Hamming distance), even if some of the genes were absent in the query MAG. We limited comparisons to genes required for the function of each respective pathway.

###### Consensus phenotype predictions

We established a procedure to reconcile inconsistent phenotype predictions between the three strategies described above, based on observing discordant gene patterns and/or discordant predicted phenotypes within a given group of neighbor genomes. In the rare case of irreconcilable disagreement between the prediction methods, assignment of a consensus phenotype defaulted to that produced by the ML method. We assigned consensus confidence scores to each prediction based on the degree of concordance between the three techniques and our confidence in the accuracy of each (see *Supplementary Discussion* and **Supplementary Table 7a**). The complete phenotype prediction process was validated using the 2,856 reference genomes in the mcSEED database, their functionally annotated genes and the accompanying patterns of presence/absence of functional metabolic pathways (curator-inferred binary phenotypes). The consensus phenotype predictions were combined into a binary phenotype matrix (BPM) containing 1,000 MAGs and 106 phenotypes (**Supplementary Table 7a**).

###### Gene annotation and phenotype prediction for Bifidobacteria-specific carbohydrate utilization pathways

We adapted the MAG annotation pipeline described above (also see **Extended Data Fig. 3**) to obtain functional annotations of genes comprising 25 additional carbohydrate utilization pathways for a set of 34 *Bifidobacterium* MAGs, followed by inference of their respective binary phenotypes. As input data for this set of *Bifidobacterium*-specific phenotypes, we curated a set of 14 metabolic subsystems in 387 reference human gut-derived *Bifidobacterium* genomes using the mcSEED framework. The reconstructed metabolic pathways and a corresponding BPM for reference *Bifidobacterium* genomes were used to predict carbohydrate utilization phenotypes in the 34 *Bifidobacterium* MAGs. Finally, the automatically generated BPM was further manually curated to account for the variability of certain pathways in this taxonomically restricted set of predictions.

###### Applying enrichment analyses to predicted MAG phenotypes

Not all successfully annotated MAG genes were components of an intact functional pathway. To enable inferred phenotype-based analysis, we filtered gene annotations to those that were part of a complete functional pathway (with a respective binary phenotype value denoted as “1”). This filter resulted in a list of 208,246 genes used for microbiome and metatranscriptome phenotype enrichment analyses.

#### Annotation of CAZymes and PULs in *P. copri* MAGs and cultured *P. copri* strains

CAZymes were annotated following the CAZy classification scheme^81^. Amino acid sequences from MAGs and isolate genomes were analyzed using a bioinformatic workflow that performs homology searches against the CAZy database^82^ and specifically accommodates the modular structure of CAZymes (which often carry a variable number of ancillary modules in addition to their catalytic domain). Details of this workflow and its application are provided in a previous publication^83^.

Polysaccharide Utilization Loci (PUL) were identified/predicted by combining information from marker genes (SusC/SusD pairs), operon structure, and CAZyme annotation information, as previously described^25^. The experimentally validated substrate specificities of CAZyme homologs contained in each PUL were used to infer the carbohydrate substrate(s) of each PUL.

We investigated the conservation of PULs within WLZ-associated *P. copri* MAGs and subsequently, between these MAGs and all other *P. copri* MAGs and cultured *P. copri* isolate genomes from this study. Because no automated method has been described to perform such a pangenome PUL comparison, we proceeded manually via the following steps. First, we predicted the PUL repertoire of Bg0018 and Bg0019 using PULDB^26^. Next, using Bg0019 as a reference, we selected PULs in Bg0018 and in other genomes for additional analysis if they displayed the same gene components in the same genomic order in both genomes. To do so, we conducted pairwise searches (blastp^65^) to identify homologous components (*e.g.*, SusC, SusD, transcriptional regulators, genes encoding CAZymes) of PULs in other *P. copri* genomes. The level of conservation for each PUL across the set of MAGs was categorized as (i) ‘conserved’ (pairwise comparisons of ORFs satisfy the requirements that their protein products have >90% amino acid identity and that the ORFs comprising the PULs being compared are organized in an identical way within the respective genomes), (ii) ‘structurally distinct’ (a given PUL is present in the genomes being compared but one or more CAZymes or one or both SusC/SusD proteins are missing or fragmented in a way likely to impact their function, or where extra PUL ORFs are present), or (iii) ‘not conserved/absent’ (PULs present in the respective genomes but with mutations likely to completely compromise function or where no PUL identified).

To relate the pattern of PUL conservation in *P. copri* MAGs to the WLZ response of study participants, we re-encoded each cell in the matrix shown in **Fig. 4a** as ‘1’ for conserved PULs, ‘0.5’ for structurally distinct PULs and ‘0’ for not conserved/ absent PULs. We imported this matrix into R, calculated a Euclidean distance between the PUL conservation pattern of Bg0019 and each additional MAG, then hierarchically clustered these patterns to generate a tree. Finally, we employed a Pearson correlation to relate the distance between the patterns of PUL conservation in Bg0019 and each additional *P. copri* MAG, to the WLZ association (β_1_ coefficient) of each MAG (**Fig. 4b**).

#### Constructing a marker gene-based phylogeny for *P. copri* MAGs and cultured *P. copri* isolates

We used CheckM^54^ (v1.1.3) to extract and align the amino acid sequences of 43 single copy marker genes in each of the 11 *P. copri* MAGs, each of the 6 cultured *P. copri* strains, plus the type strain of *Bacteroides thetaiotaomicron* VPI-5482 (accession number: 226186.12) as an outgroup. Concatenated, aligned marker gene sequences were analyzed (fasttree^84^; v2.1.10) using the Jones-Taylor-Thornton model and ‘CAT’ evolution rate to create a phylogenetic tree, which was subsequently rescaled using the ‘Gamma20’ optimization. The resulting tree was rooted to the *B. thetaiotaomicron* genome (‘ape’, v5.6-2^85^) in R prior to extracting phylogenetic distances between MAGs and isolates. The tree was visualized using ggtree^86^ (v3.2.1).

#### *In vitro* screening of glycan substrate specificity

We selected a variety of growth substrates representing putative glycan components of MDCF-2, plus glucose (Sigma, Cat # G8270) as a positive control and chondroitin sulfate (Thermo Scientific, Cat # J66156.06) as a negative control. Growth substrates included arabinan, arabinoxylan, β-glucan, galactan, galactomannan, rhamnogalacturonan, xyloglucan, and xylan (all obtained from Neogen Megazyme; **Supplementary Table 16a**). A 2% w/v solution of each polysaccharide was prepared by mixing the polysaccharide with autoclaved, filter-purified water as recommended by the supplier (see **Supplementary Table 16a** for additional details of the preparation of each solution). Each solubilized substrate was allowed to equilibrate in the anaerobic growth chamber for ∼5 days prior to use. Sterility was checked by (i) plating 10 µL aliquots of each polysaccharide preparation onto BHI+10% horse blood agar and (ii) preparing 1:1 mixtures of preparation in *P. copri* Defined Medium (PCDM). This growth assay medium was selected because (i) it was fully defined, allowing supplementation with specific carbon sources and (ii) it successfully supported the growth of *P. copri* strains in preliminary test cultures containing 5% glucose as the sole carbon source. PCDM was prepared based on the published recipe^87^ with slight modification (1 mL of a solution containing 1.9 mM hematin (H3281, Sigma) and 0.2 M L-histidine (Sigma, Cat # H6034) was added to each liter of the medium). Plates and broth were incubated for ≥3 days before checking for growth. The galactan and rhamnogalacturonan solutions displayed contamination. Therefore, a fresh 2% (w/v) solution of each of these polysaccharides was prepared and autoclaved at 121°C for 20 minutes at 15 psi before transferring them to the anaerobic chamber and confirming their sterility as described above. Autoclaved stocks were again checked for sterility before use.

We selected five *P. copri* isolates, representing a diversity of PUL conservation profiles, mcSEED BPMs, and phylogenetic assignments for *in vitro* growth assays. Freezer stocks of each isolate were plated (∼200 µL) onto BHI + 10% horse blood and grown for 48 hours at 37 °C. Isolated colonies were picked into 3 mL Wilkins-Chalgren medium and grown for 15 hours at 37 °C. Each culture was then diluted 1:100 into fresh Wilkins-Chalgren medium and grown for an additional six hours under the same anaerobic conditions at 37 °C in order to bring each culture into a similar phase of growth. Optical density (OD) at 600 nm was measured for each culture (Genesys 10S UV-Vis, Thermo Scientific) and OD_600_ was standardized to 0.02 by dilution into a final volume of 1 mL of PCDM with no added carbon source in a 96 x 1.3 mL deep well plate (Nunc, Thermo Fisher); this procedure allows us to have a uniform inoculum for all test isolates. Nine milliliters of a 1:1 mixture of 2X PCDM containing 2% (w/v) glycan or control substrate was prepared in 50 mL polypropylene tubes (Corning) yielding 11 tubes of medium, each with an individual carbon source. These media plus the OD-standardized cultures were sealed (Alumaseal, Excel Scientific) and transferred to an anaerobic chamber containing a BioTek Precision XS liquid handling robot. The robot was used to aliquot 180 μL of each type of medium into corresponding columns of three 96-well flat-bottom tissue culture plates with a well capacity of 340 µL (TPP, Sigma). A separate procedure on the Precision XS robot was used to mix and aliquot 20 µL of each OD_600_-standardized culture into corresponding rows of each assay plate; one row was reserved as a no-inoculation control. Assay plates were then sealed with optically clear films (Axygen, Cat # UC-500) and placed in the input stack of a BioTek BioStack 4 microplate stacker configured to load a BioTek Eon microplate spectrophotometer. Prior to initiating the experiment, two heated catalyst boxes were used to warm the chamber to 37 °C. The microplate stacker and reader were used to perform one week of continuous monitoring, with OD_600_ readings obtained every 15 minutes. Each assay plate was agitated for five seconds prior to each OD reading. At the conclusion of the experiment, data were exported in tabular form and analyzed in R using custom scripts to determine maximal growth rate and OD_600_ for each replicate of each culture in each glycan. Extracted curve parameters were normalized to growth rates determined in PCDM containing 5% glucose as the sole carbon source.

### Supplementary Tables

**Supplementary Table 1. Metagenome-Assembled Genome (MAG) and isolate genome assembly characteristics and taxonomic assignments.** (**a**) MAG assembly characteristics. (**b**) Genome assembly characteristics for *P. copri* isolates.

**Supplementary Table 2. Abundance and prevalence data for the 1000 MAGs identified in participants of the clinical study.** (**a**) Mapping statistics of paired-end Illumina reads pseudoaligned to the set of 1000 MAGs. (**b**) Abundance and prevalence statistics of MAGs in fecal samples collected from participants in the MDCF-2 and RUSF treatment groups (n = 59 children/group).

**Supplementary Table 3. Relationship between MAG abundance and ponderal growth (WLZ) described in Fig. 1b**. (**a**) MAG associations across all fecal samples with corresponding anthropometry from both treatment groups. (**b**) MAG abundance associations with WLZ across fecal samples with corresponding anthropometry, excluding the 1-month post-treatment time point.

**Supplementary Table 4. MAG abundance responses to MDCF-2 compared to RUSF treatment as described in Fig. 1c,d**. (**a**) MAG differential abundance analysis of the effect of treatment group. (**b**) MAG differential abundance analysis of the effect of the interaction between treatment group and study week.

**Supplementary Table 5. Annotation of MAG genes by mcSEED- and CAZy-based analyses.** (**a**) mcSEED-based metabolic pathway (phenotype) glossary. (**b**) Annotation of genes in the set of 1,000 MAGs based on mcSEED metabolic pathways or CAZymes in the CAZy database. (**c**) Annotation of genes in MAGs comprising an additional set of carbohydrate utilization pathways curated in detail only for Bifidobacterium species. (**d**) Annotation of genes in six *P. copri* isolates based on mcSEED metabolic pathways and CAZymes in the CAZy database.

**Supplementary Table 6. Representation of components of 106 metabolic pathways in 1,000 MAGs based on gene annotations from the mcSEED database.**

**Supplementary Table 7. mcSEED-based binary phenotype matrix (BPM) predictions for 1,000 MAGs and six *P. copri* isolates.** (**a**) BPM for 106 metabolic phenotypes predicted in 1,000 MAGs. (**b**) BPM for 25 carbohydrate-related phenotypes predicted in 34 Bifidobacterium MAGs and reference genomes. (**c**) BPM predictions for six *P. copri* isolates plus the WLZ-associated *P. copri* MAGs Bg0018 and Bg0019.

**Supplementary Table 8. Enrichment of metabolic pathways in MAGs ranked by WLZ-association or by change in abundance in response to MDCF-2 compared to RUSF treatment as described in Extended Data Fig. 4**. (**a**) GSEA for the presence or absence of a functional pathway in MAGs ranked by WLZ association. (**b**) GSEA of pathway enrichment in MAGs ranked by change in abundance in response to MDCF-2 compared to RUSF treatment (’treatment group’ coefficient). **(c)** GSEA of metabolic pathways in MAGs ranked by change in abundance in response to MDCF-2 compared to RUSF treatment (interaction between ‘treatment group’ and ‘study week’ coefficients).

**Supplementary Table 9. Mass spectrometric analysis of glycans in MDCF-2, RUSF, and their component ingredients as described in Fig. 2**, **Fig. 5, Extended Data Fig. 5 and Extended Data Fig. 10.** (**a**) Composition of MDCF-2 and RUSF. (**b**) Total monosaccharide content (µg monosaccharide /mg of dried therapeutic food formulation or its ingredients). (**c**) Glycosidic linkage composition (peak area, arbitrary units /ng of dried diet or ingredient). (**d**) Polysaccharide composition (FITDOG, µg polysaccharide/mg of dried diet or ingredient). (**e**) Difference in monosaccharide composition between MDCF-2 and RUSF (µg of monosaccharide /mg of dried diet). (**f**) Difference in glycosidic linkage content between MDCF-2 and RUSF (peak area, arbitrary units/ng of dried diet). (**g**) Difference in polysaccharide content between MDCF-2 and RUSF (µg polysaccharide/mg of dried diet).

**Supplementary Table 10. Principal Components Analysis (PCA) of samples by MAG abundance or MAG transcript abundance as described in Fig. 3 and Extended Data Fig. 6**. (**a**) Projection of each sample along the three principal components explaining the most variance for the PCA of samples by MAG abundances. (**b**) Contribution and projection of each MAG to the three principal components explaining the most variance for the PCA of samples by MAG abundances. (**c**) Results of enrichment analysis of WLZ-associated MAGs along the three principal components explaining the most variance for the analysis of samples by MAG abundances. (**d**) Projection of each sample along the three principal components explaining the most variance for the PCA of samples by MAG transcript abundances. (**e**) Contribution and projection of each MAG transcript to the three principal components explaining the most variance for the PCA of samples by MAG transcript abundances.

**Supplementary Table 11. MAG transcripts associated with MDCF-2 versus RUSF treatment group and the interaction of treatment group and study week as described in Fig. 3b**. (**a**) Differential gene expression for the treatment group coefficient (edgeR analysis, MDCF-2 compared to RUSF). (**b**) Differential gene expression for the interaction coefficient of treatment group (MDCF-2 compared to RUSF) and study week (edgeR analysis).

**Supplementary Table 12. Enriched expression of functional metabolic pathways and their leading-edge transcripts as determined by ranking of MAG transcripts by response to MDCF-2 compared to RUSF treatment group and the interaction of treatment group and study week as described in Fig. 3b**.

**Supplementary Table 13. MAG transcripts associated with MDCF-2 WLZ-response quartile and the interaction of WLZ-response quartile and study week as described in Fig. 3c**. (**a**) Differential gene expression analysis for upper compared to lower WLZ-response quartile. (**b**) Differential gene expression analysis for the interaction of WLZ-response quartile and study week.

**Supplementary Table 14. Enriched expression of functional metabolic pathways and their leading-edge transcripts as determined by ranking MAG transcripts in MDCF-2 participants by their upper versus lower WLZ quartile response and the interaction of WLZ-response quartile and study week as described in Fig. 3c**. (**a**) Carbohydrate utilization pathway enrichment. (**b**) Vitamin and amino acid synthesis pathways enrichment. (**c**) Summary of leading-edge transcripts assigned to transcriptionally active, WLZ-associated *P. copri* or *B. longum* MAGs.

**Supplementary Table 15. Genomic conservation and expression of polysaccharide utilization loci (PULs) in *P. copri* MAGs and isolates described in Fig. 4**. (**a**) PUL conservation in *P. copri* MAGs. (**b**) PUL conservation in cultured Bangladeshi *P. copri* isolates.

**Supplementary Table 16. *In vitro* growth assays of *P. copri* strains isolated from Bangladeshi children described in Fig 4c,d**. (**a**) Purified polysaccharides selected for *in vitro* screening. (**b**) Growth parameters for each isolate grown in defined medium containing each polysaccharide as the sole carbon source. (**c**) Relationship between PUL-based predicted polysaccharide metabolism for each strain and observed growth for each strain in defined medium containing that polysaccharide. (**d**) Consumption of monosaccharides by strains grown in defined medium containing each purified polysaccharide as the sole carbon source.

**Supplementary Table 17. Mass spectrometric analysis of the fecal glycans in MDCF-2-treated children described in Fig. 5. (a)** Total monosaccharide content in fecal samples collected from MDCF-2 treated participants (µg of monosaccharide /mg of dried fecal sample). (**b**) Glycosidic linkage composition of fecal samples collected from MDCF-2 treated participants (peak area, arbitrary units/ng of dried fecal sample).

**Supplementary Table 18. Relating MDCF-2-responsive transcripts to changes in levels of fecal glycosidic linkages as described in Fig. 5**. (**a**) Changes in fecal glycosidic linkage levels over time in upper compared to lower WLZ quartile responders. (**b**) Spearman correlation between reported dietary intake (food frequency questionnaire) and levels of glycosidic linkages detected in the fecal samples of upper and lower WLZ quartile responders among participants treated with MDCF-2. (**c**) Analysis of food frequency questionnaire responses grouped by WLZ response quartile.

## Supplementary Discussion

### Short-read sequencing only versus hybrid (long- and short-read) MAG assembly

We explored the impact of the addition of long-read sequencing data on various quality characteristics of MAGs assembled from data collected from the 0- and 3-month time points from all upper WLZ quartile responders (n=15) in the MDCF-2 treatment group. In our final set of high-quality, dereplicated MAGs, 918 MAGs represented contigs assembled from short-read only data, while 82 were derived from hybrid short and long-read assemblies (**Supplementary Table 1**). Although the mean quality characteristics of MAGs from each assembly type did not differ in completeness (determined by marker gene analysis) or total length, MAGs derived from hybrid assemblies displayed a significantly lower rate of contamination, fewer contigs, and greater N50 (see **Extended Data Fig. 1b** for statistical analyses).

### Comparing MAG assembly accuracy and quantitation using a pseudoalignment and expectation maximization approach

MAG assembly algorithms that synthesize both contig sequence characteristics and contig abundance to assemble MAGs (*e.g*., MaxBin2^49^, MetaBAT2^50^) require accurate contig quantitation. Alignment-free quantitation approaches (*e.g.*, kallisto) have demonstrated superior speed and accuracy compared to read-mapping-based quantitation in the context of metagenomic analyses where read-mapping ambiguity is common^59^.

We investigated the utility of kallisto-based quantitation for (i) contigs, prior to MAG assembly, and (ii) MAGs themselves after assembly and curation. For this analysis, we employed a ‘mouse gut metagenome toy dataset’ from CAMI II that included 64 ‘mock fecal samples’; these mock samples were produced using sequencing data from 791 publicly available bacterial genomes (representing 549 species) and genomic abundances that mirrored bacterial 16S rRNA gene profiles of 64 actual mouse fecal biospecimens^88^. We utilized three components of this reference dataset for our analyses: (i) simulated sequencing data (1.67×10^7^ Illumina paired-end 150 nt reads) from each of the 64 mock fecal samples, (ii) anonymized reference contigs from the 791 reference genomes, and (iii) reference abundances of contigs/genomes in each fecal sample.

First, we investigated the effect of kallisto quantification of contigs on the fidelity of MAG assembly. To do so, we quantified the reference contigs using either kallisto or bowtie2 and the short-read simulated Illumina data. Next, we assembled MAGs using MaxBin2^49^, MetaBAT2^50^ and CONCOCT^89^ with data from either kallisto or bowtie2 contig quantitation as input. The output of each MAG assembly method for each sample was combined using DAS Tool^51^. Finally, each MAG set was compared against 791 intact reference genomes using AMBER^90^ (**Extended Data Fig. 12a,b**). MAGs generated using kallisto contig quantification and DAS Tool dereplication were more complete (*P*=6.4×10^-14^; Wilcoxon test) and less contaminated (*P*<2.2×10^-16^; Wilcoxon test) than those generated using bowtie2 (**Extended Data Fig. 12c**). Additionally, a significantly greater number of MAGs (*P*<0.05; Fisher’s exact test) were detected using kallisto contig quantitation (**Extended Data Fig. 12d**).

Next, we employed the same simulated dataset to test the accuracy of kallisto-based MAG quantitation. We mapped the short-read data for each of the 64 fecal samples to the set of 791 reference genomes using kallisto and bowtie2. We then correlated the abundance profiles generated by each quantitation method to the ‘true’ abundance profile for each sample. The correlations between true genome abundances and kallisto genome abundances were stronger than those calculated using bowtie (mean Pearson’s *r*^2^= 0.99 for kallisto versus *r*^2^= 0.97 for bowtie; *P*<2.2×10^-16^, Wilcoxon test comparing each distribution of correlation coefficients).

We determined the false positive and false negative rate of MAG detection across all samples. Notably, kallisto quantitation resulted in more false positive abundances across the 64 mock fecal samples [300.2±50.1 versus 69.3±28.4 for bowtie2, respectively (mean±SD); *P*< 2.2×10^-16^, Wilcoxon test] while bowtie2 quantitation resulted in more false negative abundances [0.09±0.42 versus 17.2±26.1 (mean±SD), respectively; *P*<2.2×10^-16^, Wilcoxon test]. Importantly, analysis of the average values of false positive abundance generated using kallisto suggested that a low abundance filter would significantly reduce the false positive rate. For example, applying a filter to this dataset that required >5 TPM for a MAG to be designated as ‘detected’ resulted in a false positive rate significantly lower than that of bowtie2 (*P*=0.02, Wilcoxon test).

The greater number of high quality (less contaminated and more complete) MAGs assembled using kallisto quantitation, plus the increased accuracy of MAG quantitation using this method, led us to employ kallisto for all quantitation tasks in the MAG analysis workflow described in the current study.

### Analysis of consistency in MAG functional metabolic pathway annotation

A global comparison of binary phenotype assignments derived using the Pathway Rules (PR), Machine Learning (ML), and Neighbor Group (NG) approaches described in *Methods* revealed a remarkably low frequency of inconsistencies: in a subset of 640 MAGs where all three methods could be applied, only 4.5% of NG-based phenotype assignments were inconsistent between one or more other methods. These inconsistencies reflect different biases associated with each approach. The NG-based approach exhibits limited performance for small (<5-member) NGs with underrepresented local diversity of gene patterns. Alternatively, PR/ML-based methods appear to be less robust with respect to genome incompleteness in MAGs, resulting in omission (absence) of genes essential for the function of a pathway and, more generally, for pathways with less than three essential genes. Our consensus approach (*Methods*) resolved 70% of observed inconsistencies toward PR/ML-based assignments. In the remaining cases, a consensus phenotype was assigned in favor of the NG-based method. The overall level of inconsistencies between PR- and ML-based phenotype assignments across the entire set of 199,334 assignments in the 1,000 MAGs was much lower (<0.7%). A detailed investigation of selected cases showed that, in general, the ML-based method yielded higher accuracy phenotype assignments. Therefore, in rare cases of irreconcilable disagreement between these two methods in the set of 360 MAGs without NGs, the semi-automated assignment of the consensus phenotype was made in favor of the ML-based approach.

### Non-carbohydrate related differentially expressed transcripts in upper versus lower WLZ quartile responders

Transcripts expressed at greater levels in upper WLZ response quartile participants (β_1_ WLZ quartile term) were also enriched for pathways involved in biosynthesis of vitamin B3 and B9 and the essential amino acids tryptophan, lysine, histidine and leucine (**Supplementary Table 14b**). Leading-edge analysis revealed that *P. copri* Bg0018 and Bg0019 were major contributors to increased expression of transcripts involved in the biosynthesis of vitamins B3 and B9, plus the essential amino acids tryptophan, histidine, leucine and lysine, among the upper quartile responders, but contributed minimally (two transcripts assigned to the arginine biosynthetic pathway) to enrichment of functional pathways among the lower WLZ quartile responders (**Supplementary Table 14b**).

## Data Availability

Shotgun DNA sequencing and microbial RNA-Seq datasets generated from fecal samples, plus annotated *P. copri* isolate genome sequences are available in the European Nucleotide Archive (accession PRJEB45356). LC-MS datasets of monosaccharide, glycoside linkage and polysaccharide data are deposited in GlycoPOST (accession GPST000244).

## Biological Materials

Fecal specimens collected from Bangladeshi children are the property of icddr,b. Material Transfer Agreements exist between icddr,b and Washington University in St. Louis for the use of these samples. Requests for materials should be made to J.I.G.

## Code Availability

Code detailing the steps in the MAG assembly workflow and analyses of microbial RNA-Seq and glycan datasets are available from GitLab (https://gitlab.com/hibberdm/hibberd_webber_et_al_mdcf_poc_mags) and have been accessioned at Zenodo (DOI:10.5281/zenodo.8000098). Code for annotation of bacterial genes and prediction of metabolic phenotypes is available on request.

## References

1. Smith, M. I. et al., Gut microbiomes of Malawian twin pairs discordant for kwashiorkor. Science 339, 548–554 (2013).

2. Blanton, L. V. et al., Gut bacteria that prevent growth impairments transmitted by microbiota from malnourished children. Science 351, aad3311 (2016).

3. Gehrig, J. L., Venkatesh, S., Chang, H.-W. et al., Effects of microbiota-directed foods in gnotobiotic animals and undernourished children. Science 365, eaau4732 (2019).

4. Chen, R. Y., Mostafa, I., Hibberd, M. C. et al., A microbiota-directed food intervention for undernourished children. N. Engl. J. Med. 384, 1517–1528 (2021).

5. Wang, Y., Chang, H.-W. Lee, E. M. et al., Prevotella copri-linked effects of a therapeutic food for malnutrition. bioRxiv doi:10.1101/2023.08.11.553030 (2023).

6. United Nations Children’s Fund. United Nations Children’s Fund (UNICEF), World Health Organization, International Bank for Reconstruction and Development/The World Bank. Levels and trends in child malnutrition: key findings of the 2021 edition of the joint child malnutrition estimates. New York (2021).

7. Victoria, C. G. et al, Revisiting maternal and child undernutrition in low-income and middle-income countries: variable progress towards an unfinished agenda. Lancet 397, 1388–1399 (2021).

8. Heidkamp, R. A. et al, Mobilising evidence, data, and resources to achieve global maternal and child undernutrition targets and the sustainable development goals: an agenda for action. Lancet 397, 1400– 1418 (2021).

9. Roberton, T. et al., Early estimated of the indirect effects of COVID-19 pandemic on maternal and child mortality in low-income and middle-income countries; a modelling study. Lancet Global Health 8, e901–e908 (2020).

10. Subramanian, S. et al., Persistent gut microbiota immaturity in malnourished Bangladeshi children. Nature 510, 417–421 (2014).

11. Raman, A. S. et al., A sparse covarying unit that describes healthy and impaired human gut microbiota development. Science 365, eaau4735 (2019).

12. Giallourou, N. et al., Metabolic maturation in the first 2 years of life in resource-constrained settings and its association with postnatal growths. Sci. Adv. 6, eaay5969 (2020).

13. Kau, A. L. et al, Functional characterization of IgA-targeted bacteria taxa from undernourished Malawian children that produce diet-dependent enteropathy. Science Transl. Med. 7, 276ra24 (2015).

14. P. A. Chaumeil, A. J. Mussig, P. Hugenholtz, D. H. Parks, GTDB-Tk: a toolkit to classify genomes with the Genome Taxonomy Database. Bioinformatics 36, 1925–1927 (2020).

15. Overbeek, R. et al., The subsystems approach to genome annotation and its use in the project to annotate 1000 genomes. Nucleic Acids Res. 33, 5691–702 (2005).

16. Aziz R. K. et al., SEED servers: high-performance access to the SEED genomes, annotations, and metabolic models. PLOS One 7, e48053 (2012).

17. Rodionov, D. A. et al., Micronutrient requirements and sharing capabilities of the human gut microbiome. Frontiers Microbiol. 10, 1316 (2019).

18. Amicucci, M. J. et al., A nonenzymatic method for cleaving polysaccharides to yield oligosaccharides for structural analysis. Nat. Commun. 11, 3963 (2020).

19. Aspinall, G. O., Cottrell, I. W., Polysaccharides of soybeans. VI. Neutral polysaccharides from cotyledon meal. Can. J. Chem. 49, 1019–1022 (1971).

20. Tett, A. et al., The Prevotella copri complex comprises four distinct clades underrepresented in Westernized populations Cell Host and Microbe 26, 666–679 (2019).

21. Vatanen, et al., A distinct clade of Bifidobacterium longum in the gut of Bangladeshi children thrives during weaning, Cell 185, 4280–4297 (2022).

22. Fujita, K. et al., Degradative enzymes for type II arabinogalactan side chains in Bifidobacterium longum subsp. longum. Appl. Microbiol. Biotechnol. 103, 1299–1310 (2019).

23. Komeno, M., Hayamizu, H., Fujita, K., and Ashida, H., Two Novel α-l-Arabinofuranosidases from Bifidobacterium longum subsp. longum belonging to glycoside hydrolase family 43 cooperatively degrade arabinan. Appl. Environ. Microbiol. 85, e02582 (2019).

24. Barratt, M. J. et al. *Bifidobacterium longum* subsp. *infantis* strains for treating severe acute malnutrition in Bangladeshi infants. Science Trans. Med., 14, 640 (2022).

25. Terrapon, N., Lombard, V., Gilbert, H. J., and B. Henrissat, Automatic prediction of polysaccharide utilization loci in Bacteroidetes species. Bioinformatics 31, 647–655 (2015).

26. Terrapon, N. et al., PULDB: the expanded database of Polysaccharide Utilization Loci. Nucleic Acids Res. 46, D677–D683. (2018).

27. Palackal, N. et al., A multifunctional hybrid glycosyl hydrolase discovered in an uncultured microbial consortium from ruminant gut. Appl. Microbiol. Biot. 74, 113–124 (2007).

28. McGregor, N. et al., Structure-function analysis of a mixed-linkage β-Glucanase/Xyloglucanase from the key ruminal Bacteroidetes Prevotella bryantii B14. J. Biol. Chem. 291, 1175–1197 (2016).

## Supplementary References

29. Fehlner-Peach, H. et al., Distinct polysaccharide utilization profiles of human intestinal *Prevotella copri* isolates. Cell Host and Microbe. 26, 680–690 (2019).

30. Love, M. I., Huber, W., and Anders, S., Moderated estimation of fold change and dispersion for RNA-seq data with DESeq2. Genome Biol. 15, 550 (2014).

31. Bates, D., Mächler, M., Bolker, B., and Walker, S., Fitting linear mixed-effects models using lme4. J. Stat Softw 67, 1–48 (2015).

32. Kuznetsova, A., Brockhoff, P. B., and Christensen, R. H. B., lmerTest Package: Tests in linear mixed effects models. J. Stat. Softw. 82 (13), 1–26 (2017).

33. Hoffman, G. E. and Roussos, P., Dream: powerful differential expression analysis for repeated measures designs. Bioinformatics. 37, 192–201 (2020).

34. Krueger, F., James, F., Ewels, P., Afyounian, E., and Schuster-Boeckler B., TrimGalore (2021), doi:10.5281/zenodo.5127899.

35. Andrews, S., FastQC: A quality control tool for high throughput sequence data. https://www.bioinformatics.babraham.ac.uk/projects/fastqc/ (2010)

36. Bray, N. L., Pimentel, H., Melsted, P., and Pachter, L., Near-optimal probabilistic RNA-seq quantification. Nature Biotech. 34, 525–527 (2016).

37. McCarthy, D. J., Chen, Y., and Smyth, G. K., Differential expression analysis of multifactor RNA-Seq experiments with respect to biological variation. Nucleic Acids Res. 40, 4288–4297 (2012).

38. Korotkevich, G. et al., Fast gene set enrichment analysis. bioRxiv 060012 (2021).

39. Kassambara, A. and Mundt, F. (2020). factoextra: Extract and Visualize the Results of Multivariate Data Analyses. R package version 1.0.7. https://CRAN.R-project.org/package=factoextra

40. Oksanen, J. et al. (2020). vegan: Community Ecology Package. R package version 2.5–7. https://CRAN.R-project.org/package=vegan

41. Xu, G., Amicucci, M. J., Cheng, Z., Galermo, A. G., and Lebrilla, C. B., Revisiting monosaccharide analysis–quantitation of a comprehensive set of monosaccharides using dynamic multiple reaction monitoring. Analyst 143, 200–207 (2018).

42. Amicucci, M. J. et al., A rapid-throughput adaptable method for determining the monosaccharide composition of polysaccharides. *Intern*. J. Mass Spectrom. 438, 22–28 (2019).

43. Galermo, A. G. et al., Liquid chromatography–tandem mass spectrometry approach for determining glycosidic linkages. Anal. Chem. 90, 13073–13080 (2018).

44. Galermo, A. G., Nandita, E., Castillo, J. J., Amicucci, M. J., and Lebrilla, C. B., Development of an extensive linkage library for characterization of carbohydrates. Anal. Chem. 91, 13022–13031 (2019).

45. Nandita, E. et al., Polysaccharide identification through oligosaccharide fingerprinting. Carbohydr. Polym. 257, 117570 (2021).

46. Baym, M. et al., Inexpensive multiplexed library preparation for megabase-sized genomes. PLOS One 10, e0128036 (2015).

47. Langmead, B. and Salzberg S. L., Fast gapped-read alignment with Bowtie 2. Nature Methods 9, 357–359 (2012).

48. Li, D., Liu, C. M., Luo, R., Sadakane, K., and Lam, T. W., MEGAHIT: an ultra-fast single-node solution for large and complex metagenomics assembly via succinct de Bruijn graph. Bioinformatics 31, 1674–1676 (2015).

49. Wu, Y. W., Simmons, B. A., and Singer, S. W., MaxBin 2.0: an automated binning algorithm to recover genomes from multiple metagenomic datasets. Bioinformatics 32, 605–607 (2016).

50. Kang, D. D. et al., MetaBAT 2: an adaptive binning algorithm for robust and efficient genome reconstruction from metagenome assemblies. PeerJ. 7, e7359 (2019).

51. Sieber, C. M. K. et al., Recovery of genomes from metagenomes via a dereplication, aggregation and scoring strategy. Nature Microbiol. 3, 836–843 (2018).

52. Bertrand, D. et al., Hybrid metagenomic assembly enables high-resolution analysis of resistance determinants and mobile elements in human microbiomes. Nature Biotech. 37, 937–944 (2019).

53. Walker, B. J. et al., Pilon: An integrated tool for comprehensive microbial variant detection and genome assembly improvement. PLoS ONE 9, e112963 (2014).

54. Parks, D. H., Imelfort, M., Skennerton, C. T., Hugenholtz, P., and Tyson, G. W., CheckM: assessing the quality of microbial genomes recovered from isolates, single cells, and metagenomes. Genome Res. 25, 1043–1055 (2015).

55. Nayfach, S., Shi, Z. J., Seshadri, R., Pollard, K. S., and Kyrpides, N. C., New insights from uncultivated genomes of the global human gut microbiome. Nature 568, 505–510 (2019).

56. Olm, M. R., Brown, C. T., Brooks, B., and Banfield, J. F., dRep: a tool for fast and accurate genomic comparisons that enables improved genome recovery from metagenomes through de-replication. ISME J. 11, 2864–2868 (2017).

57. Gurevich, A., Saveliev, V., Vyahhi, N., and G. Tesler, QUAST: quality assessment tool for genome assemblies. Bioinformatics 29, 1072–1075 (2013).

58. Seemann, T., Prokka: rapid prokaryotic genome annotation. Bioinformatics. 30, 2068–2069 (2014).

59. Schaeffer, L., Pimentel, H., Bray, N., Melsted, P., and Pachter, L., Pseudoalignment for metagenomic read assignment. Bioinformatics 33, 2082–2088 (2017).

60. Wood, D. E., Lu, J. and Langmead, B., Improved metagenomic analysis with Kraken 2. Genome Biol. 20, 257 (2019).

61. Lu, J., Breitwieser, F. P., Thielen, P., and Salzberg, S., Bracken: estimating species abundance in metagenomics data. PeerJ. Computer Science 3, e104 (2017).

62. Asnicar, F. et al., Precise phylogenetic analysis of microbial isolates and genomes from metagenomes using PhyloPhlAn 3.0. Nature Commun. 11, 2500 (2020).

63. Asnicar, F., Weingart, G., Tickle, T. L., Huttenhower, C., and Segata, N., Compact graphical representation of phylogenetic data and metadata with GraPhlAn. PeerJ 3, e1029 (2015).

64. Page, A. J. et al., Roary: rapid large-scale prokaryote pan genome analysis. Bioinformatics. 31, 3691– 3693 (2015).

65. Altschul, S. F., Gish, W., Miller, W., Myers, W. W., and Lipman, D. J., Basic local alignment search tool. J. Mol. Biol. 215, 403–410 (1990).

66. Katoh, K. and Standley, D. M., MAFFT multiple sequence alignment software version 7: improvements in performance and usability. Mol. Biol. Evol. 30, 772–780 (2013).

67. Snipen, L. and Hovde Liland, K. (2021). microseq: Basic Biological Sequence Handling. R package version 2.1.5. https://CRAN.R-project.org/package=microseq

68. Nguyen, L. T., Schmidt, H. A., von Haeseler, A., and Minh, B. Q., IQ-TREE: A fast and effective stochastic algorithm for estimating maximum-likelihood phylogenies. Mol. Biol. Evol. 32, 268–274 (2015).

69. Han, N. D. et al., Microbial liberation of *N*-methylserotonin from orange fiber in gnotobiotic mice and humans. Cell 185, 2495–2509 (2022).

70. Kolmogorov, M. et al., Assembly of long, error-prone reads using repeat graphs. Nature Biotechnology 37, 540–546 (2019).

71. Davis, J. J. et al., The PATRIC Bioinformatics Resource Center: expanding data and analysis capabilities. Nucleic Acids Res. 48, D606–D612 (2020).

72. Ye, Y., Osterman, A., Overbeek, R., and Godzik, A., Automatic detection of subsystem/pathway variants in genome analysis. Bioinformatics. 21, i478–86 (2005).

73. Steinegger, M. and Söding, J., MMseqs2 enables sensitive protein sequence searching for the analysis of massive data sets. Nat. Biotechnol. 35,1026–1028 (2017).

74. Buchfink, B., Reuter, K., and Drost, H. G., Sensitive protein alignments at tree-of-lifescale using DIAMOND. Nat. Methods 18, 366–368 (2021).

75. Hyatt, D. et al., Prodigal: prokaryotic gene recognition and translation initiation site identification. BMC Bioinformatics 11, 119–119 (2010).

76. Abraham, A. et al., Machine learning for neuroimaging with scikit-learn. Front. Neuroinform. 8, 14 (2014).

77. Pedregosa, F. et al., Scikit-learn: Machine learning in python. J. Machine Learning Res. 12, 2825– 2830 (2011).

78. Therneau, T., Atkinson, B., and Ripley, B., Rpart: Recursive Partitioning (2013). https://github.com/bethatkinson/rpart/

79. Kuhn, M., Building Predictive models in R using the caret package. J. Stat.Softw. 28, 1–26 (2008).

80. Ondov, B. D. et al., Mash: fast genome and metagenome distance estimation using MinHash. Genome Biol. 17,132 (2016).

81. Drula, E. et al., The carbohydrate-active enzyme database: functions and literature. Nucleic Acids Res. (2021).

82. Lombard, V., Ramulu, H. G., Drula, E., Coutinho, P. M., and Henrissat, B., The carbohydrate-active enzymes database (CAZy) in 2013. Nucleic Acids Res.42, D490–D495 (2014).

83. Delannoy-Bruno, O. et al., Evaluating microbiome-directed fibre snacks in gnotobiotic mice and humans. Nature 595, 91–95 (2021).

84. Price, M. N. et al. FastTree 2 – Approximately maximum-likelihood trees for large alignments. PLoS One 5(**3**), e9490 (2010).

85. Paradis, E. and Schliep, K. ape 5.0: an environment for modern phylogenetics and evolutionary analyses in R. Bioinformatics 35, 526–528. (2018).

86. Yu, G. Using ggtree to visualize data on tree-like structures. Curr. Protoc. Bioinformatics 69, e96. (2020).

87. Li, J. et al., A versatile genetic toolbox for Prevotella copri enables studying polysaccharide utilization systems. EMBO J, 40, e108287 (2021).

88. Meyer, F. et al., Tutorial: Assessing metagenomics software with the CAMI benchmarking toolkit. Nature Protocols, 16, 1785–1801 (2021).

89. Alneberg, J. et al., Binning metagenomic contigs by coverage and composition. Nature Methods 11, 1144–1146 (2014).

90. Meyer, F. et al., AMBER: Assessment of metagenome BinnERs. GigaScience 7, giy069 (2018).

